# Genetic mechanisms of mitotic error in 840,853 blood genomes

**DOI:** 10.64898/2026.09.08.26362505

**Authors:** Yichen Si, Nolan Kamitaki, David Tang, Po-Ru Loh

## Abstract

Mosaic aneuploidies in leukocytes are commonly detectable in aging individuals. Here, we studied inherited genetic influences on susceptibility to such aneuploidies by precisely quantifying chromosomal ploidy from whole-genome sequencing read-depth in 840,853 individuals. This approach revealed autosomal gains in ∼70,000 individuals (7-fold more than previous data sets) and ubiquitous loss of the X chromosome in females and loss of Y in males, each of which exhibited *>*50% heritability. Genome-wide association analyses of 12 mosaic aneuploidies identified *>*800 common inherited influences on the generation and proliferation of aneuploid cells. Variants in mitotic spindle assembly checkpoint genes consistently affected aneuploidy of most chromosomes, whereas a missense variant in *PMF1* (which encodes a subunit of the outer kinetochore) associated with increased segregation fidelity of some chromosomes at the expense of others. Structural variation within multiple chromosomes associated with allele-specific effects on missegregation rates. An inherited hybrid centromere formed by an ancient recombination of 22q with 13p—generating the largest common structural variant in the human genome—associated with 4-fold greater risk of chromosome 22 gain. Abundant copy number and sequence variation in two X-linked CTCF-binding repeat arrays, *ICCE* and *DXZ4*, associated with a 4-fold range in the likelihoods of different X chromosomes to be lost. These results show how genetic risk of chromosomal gains and losses varies greatly across chromosomes and across people, providing instruments for assessing the effects of mosaic aneuploidies on health.

## Main

Age-related somatic aneuploidies were first observed in cytogenetic studies of cultured lymphocytes^1^ and inferred to primarily involve loss of the X chromosome (LOX) in females and loss of Y (LOY) in males^2^. High-throughput SNP-array studies subsequently observed a wide variety of hematopoietic mosaic chromosomal alterations (mCAs) in elderly individuals^3, 4^, with LOY and LOX (also termed mLOY and mLOX) the most common^5, 6^. These and other large-scale studies identified strong associations of hematopoietic autosomal mCAs with incident blood cancer risk^3, 4^ and LOY with many clinical outcomes^5, 7–10^ with varying support for causality^10^.

Susceptibility to sex chromosome loss in blood has a strong genetic component: genome-wide association studies (GWAS) have identified hundreds of genetic risk loci influencing LOY^11, 12^ and 42 loci for LOX^13^, implicating genes involved in chromosome segregation, cell-cycle regulation, and DNA damage response^11–13^. Autosomal copy-neutral loss of heterozygosity (CN-LOH) mutations are also known to act on rare inherited variation in *cis*, acquiring selective advantage by changing the allelic dosage of such variants^14–17^. However, the factors that influence susceptibility to autosomal aneuploidies are largely unknown: GWAS have found just one robustly replicated association of a *MAD1L1* haplotype with chromosome 15 gain^16^ and five risk loci for autosomal mCAs in aggregate^15, 18^.

Autosomal aneuploidies have been difficult to study in GWAS cohorts because despite commonly being present within blood cell populations^19^, they rarely affect sufficiently many cells to be detectable using existing tools for analyzing bulk DNA data, even from whole-genome sequencing (WGS)^17, 20^. Consequently, whereas GWAS have analyzed over 100,000 cases of LOX^13^ and LOY^12^, the most common autosomal aneuploidies have been found in only ∼1,000 participants^16, 17^. This has precluded comparing genetic effects on aneuploidy across different human chromosomes—which are known to exhibit chromosome-specific missegregation biases in cell line experiments^21–23^, and which harbor abundant centromeric sequence variation^24–26^ with unknown functional consequences. Additionally, even for LOX and LOY, key questions regarding the source of allelic bias in LOX^13, 14^ and the causality of LOX and LOY-associated health outcomes^10, 13^ have remained unanswered.

### Age-acquired mosaic aneuploidy in blood DNA from 840,853 individuals

To investigate these questions, we developed a new computational method for sensitively detecting and quantifying mosaic aneuploidies from blood-derived WGS data available for 490,089 UK Biobank (UKB)^27^ and 350,764 All of Us (AoU)^28^ participants. Mosaic aneuploidies increase or decrease representation of the affected chromosome relative to other chromosomes, such that they can be detected from deviations in chromosome-wide sequencing coverage. This intuitive approach has been difficult to apply to population cohorts because technical variation in sequencing coverage usually overwhelms the slight deviations in coverage generated by subtle levels of aneuploidy in healthy blood. Here, we overcame this challenge using a new approach for denoising WGS read-depth profiles^17^, which enabled precise calibration of chromosome-wide coverage (relative error *<*0.001; Methods and Supplementary Tables 1–2).

We first used this approach to precisely quantify fractions of leukocytes with sex chromosome losses across population cross-sections spanning the human adult lifespan (18 to 90+ years of age), yielding surprising insights into the ubiquity and clonal trajectories of X and Y loss. Median LOX cell fractions increased slowly but steadily with age (to ∼2% in female 80-year-olds), and population variation in LOX was modest (typically within 2-fold of the median; Fig. 1a and Supplementary Figs. 1–3). In contrast, LOY cell fractions increased exponentially with age, with much greater variance: most 80-year-old males had LOY in ∼1–10% of leukocytes, but a few percent had *>*50% LOY or no detectable LOY (Fig. 1b and Supplementary Figs. 1–3). Moreover, whereas LOY accelerated with increasing age (Fig. 1b)—suggesting exponential growth driven by clonal selection—LOX did not (Fig. 1a), suggesting little or no clonal proliferation, and hinting that the near-linear trajectory of LOX might derive from recurrent mitotic missegregation. Reflecting these contrasting dynamics, median LOX cell fractions in younger females exceeded median LOY in age-matched males until age ∼60 (Supplementary Fig. 4). These results from denoised WGS read-depth analyses were consistent between UKB and AoU (Supplementary Fig. 3) and passed several validation checks for robustness (Supplementary Figs. 5–9 and Supplementary Note 1).

**Figure 1.**
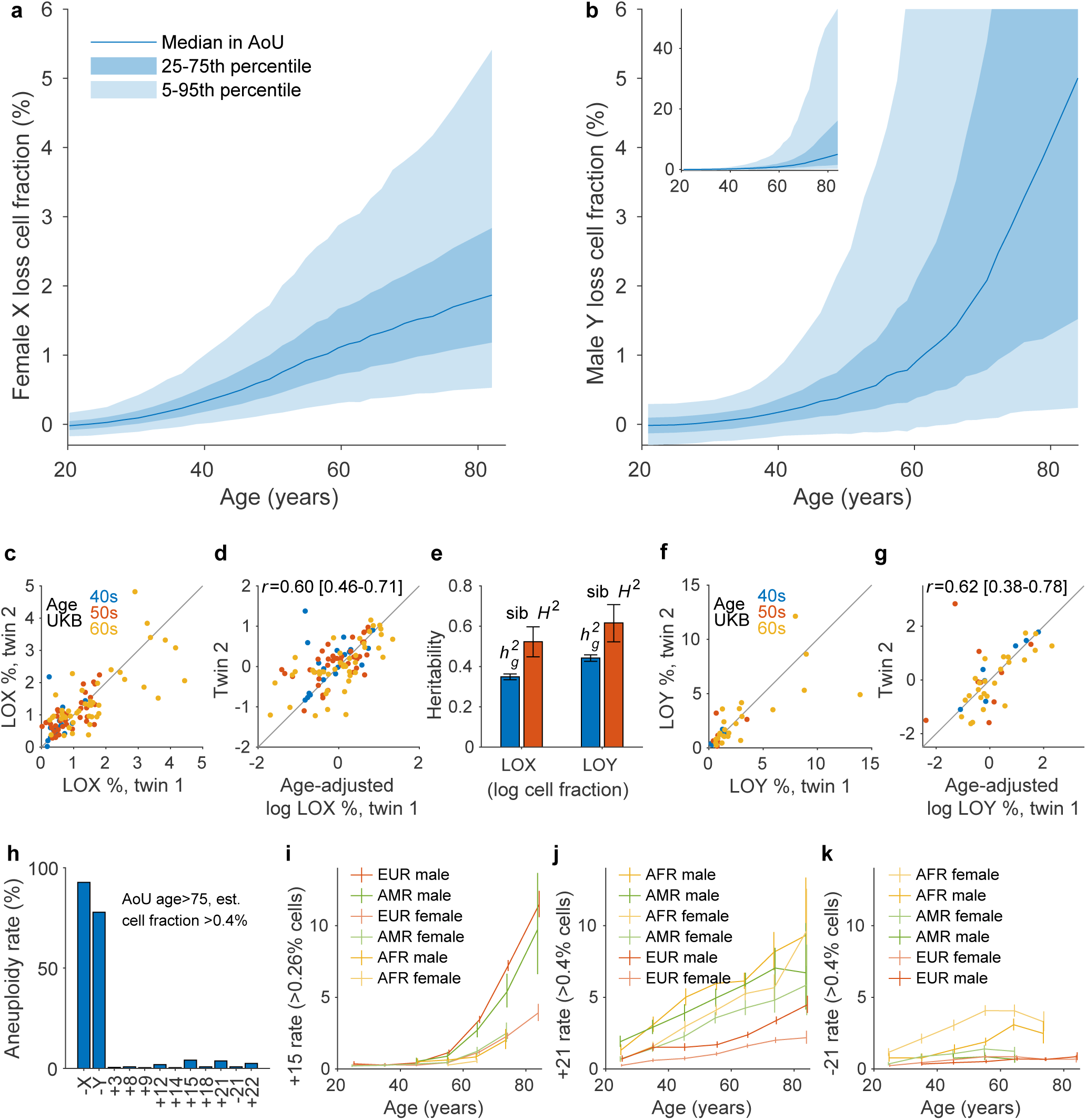
Aneuploidies in blood commonly accrue with age and are strongly influenced by inherited genetic variation. **a**, **b**, Estimated fractions of X loss in blood DNA from female AoU participants (**a**; n=206,847) and Y loss in males (**b**; n=138,703). Participants were stratified into age tranches; curves indicate medians (light and dark blue shaded regions, 5^th^–95^th^ and 25^th^–75^th^ percentiles) within each tranche. **c**, **d**, Twin concordance of X loss levels in UKB (adjusted for age in **d**). **e**, Heritability of age-adjusted X and Y loss levels estimated using SNP-array genotypes (*h*_g_^2^) and sibling pairs (sib *H*^2^). **f**, **g**, Twin concordance of Y loss levels in UKB (adjusted for age in **g**). In panel **f**, one twin pair with a Y loss percentage exceeding the plot range is not visible. **h**, Rates of 12 common aneuploidies in AoU participants over age 75 (n=27,828). To facilitate comparison across different aneuploidies, positivity for each aneuploidy was defined here as an estimated leukocyte fraction of *>*0.4% (based on WGS read-depth) even though aneuploidies of longer chromosomes are detectable at lower cell fractions. **i**, Rates of detecting gain of chromosome 15 in *>*0.26% of leukocytes in AoU participants stratified by ancestry, sex, and age (n=348,956). **j**, **k**, Analogous plots for gain or loss of chromosome 21 detected in *>*0.4% of leukocytes. Error bars, 95% CIs.

Beyond age, the main covariate associated with levels of both LOX and LOY was genetic ancestry: European-ancestry individuals exhibited higher LOY^29^ but lower LOX (Supplementary Fig. 10a,b). Other covariate associations were largely specific to either LOX or LOY: LOX cell fraction increased with lymphocyte percentage^13^ (Supplementary Fig. 11), while LOY (but not LOX) levels were higher in current smokers^30, 31^ (Supplementary Fig. 10c,d) and individuals with lower body mass index^12, 32^ (Supplementary Fig. 12).

Multiple other lines of evidence supported a primary role of genetics in determining LOX and LOY levels. Monozygotic twin pairs in UKB exhibited strikingly high concordance in LOX levels between female twins and LOY between male twins (*R*=0.60 [95% CI, 0.46–0.71] and 0.62 [0.38–0.78] adjusting for age; Fig. 1c,d,f,g). Correlations among siblings corroborated these results (suggesting 52% [45%–60%] heritability of age-adjusted LOX levels and 62% [52%–71%] heritability of LOY), and SNP-heritability estimates were modestly lower as expected^33^ (Fig. 1e). As such, female and male propensities for leukocytic X and Y loss are among the most heritable of human complex traits studied to date^34^. The extent of this heritability was not previously discernible owing to less precise ascertainment of LOX and LOY (as binary phenotypes for detectable LOX/LOY)^11, 13, 14^.

Autosomal aneuploidies were also commonly detectable from denoised WGS read-depth, at much higher rates than previously observed^15–17, 20^. Focusing on 10 autosomal copy-number alterations (9 gains and 1 loss) for which most mCAs are whole-chromosome events^17, 20^, these aneuploidies were detectable at a combined frequency of nearly 10% (∼70,000 events across UKB and AoU; Fig. 1h and Supplementary Table 2) and exhibited known sex biases^15^ (Supplementary Fig. 13). Gains of chromosomes 15 and 21 were the most common autosomal aneuploidies, and in African-ancestry participants, loss of chromosome 21 was also common (Fig. 1i–k). Similar to LOX and LOY, autosomal aneuploidies showed varying age trajectories and enrichments in different genetic ancestries (Supplementary Fig. 14), again pointing to strong genetic influences.

### Distinct germline drivers of different hematopoietic aneuploidies

To explore the genetic basis of hematopoietic aneuploidies, we optimized a set of WGS-based aneuploidy phenotypes for association analysis in UKB and AoU (Supplementary Table 3), enabling much more powerful GWAS than previously possible. For LOX and LOY (detectable in most individuals), we computed log-transformed cell fractions (Supplementary Fig. 15); in UKB, these phenotypes generated several-fold stronger genetic associations than previous SNP-array allelic imbalance-based binary phenotypes (∼20x stronger for LOX, ∼3x for LOY; Supplementary Fig. 16). For autosomes, we computed probabilistic aneuploidy calls (Supplementary Fig. 17), capturing several-fold more aneuploidies than allelic imbalance-based analyses^15, 17^ (Supplementary Fig. 18a,b). Most were whole-chromosome aneuploidies, with the remainder primarily arm-level events (consistent with centromeric fission potentially caused by split kinetochores^35^; Supplementary Figs. 18c and 19).

GWAS of the 12 aneuploidy phenotypes in UKB identified 828 lead associations (*P*=5×10^-8^–7×10^-1240^) of inherited common or low-frequency variants (minor allele frequency *>*0.1%) with hematopoietic aneuploidies (Fig. 2a, Supplementary Figs. 20–30, Supplementary Tables 4–6). These included 173 lead associations with autosomal aneuploidies (172 novel) and 245 lead associations with LOX (∼6-fold the number previously identified^13^). The associations replicated robustly in AoU, with 97% exhibiting consistent effect directions (Supplementary Fig. 31 and Supplementary Tables 4–6). GWAS in AoU produced broadly concordant results while identifying several additional associations involving variants or phenotypes (e.g., chromosome 21 loss) enriched in non-European ancestries (Supplementary Figs. 21–30, Supplementary Tables 7–9, and Supplementary Note 2).

**Figure 2.**
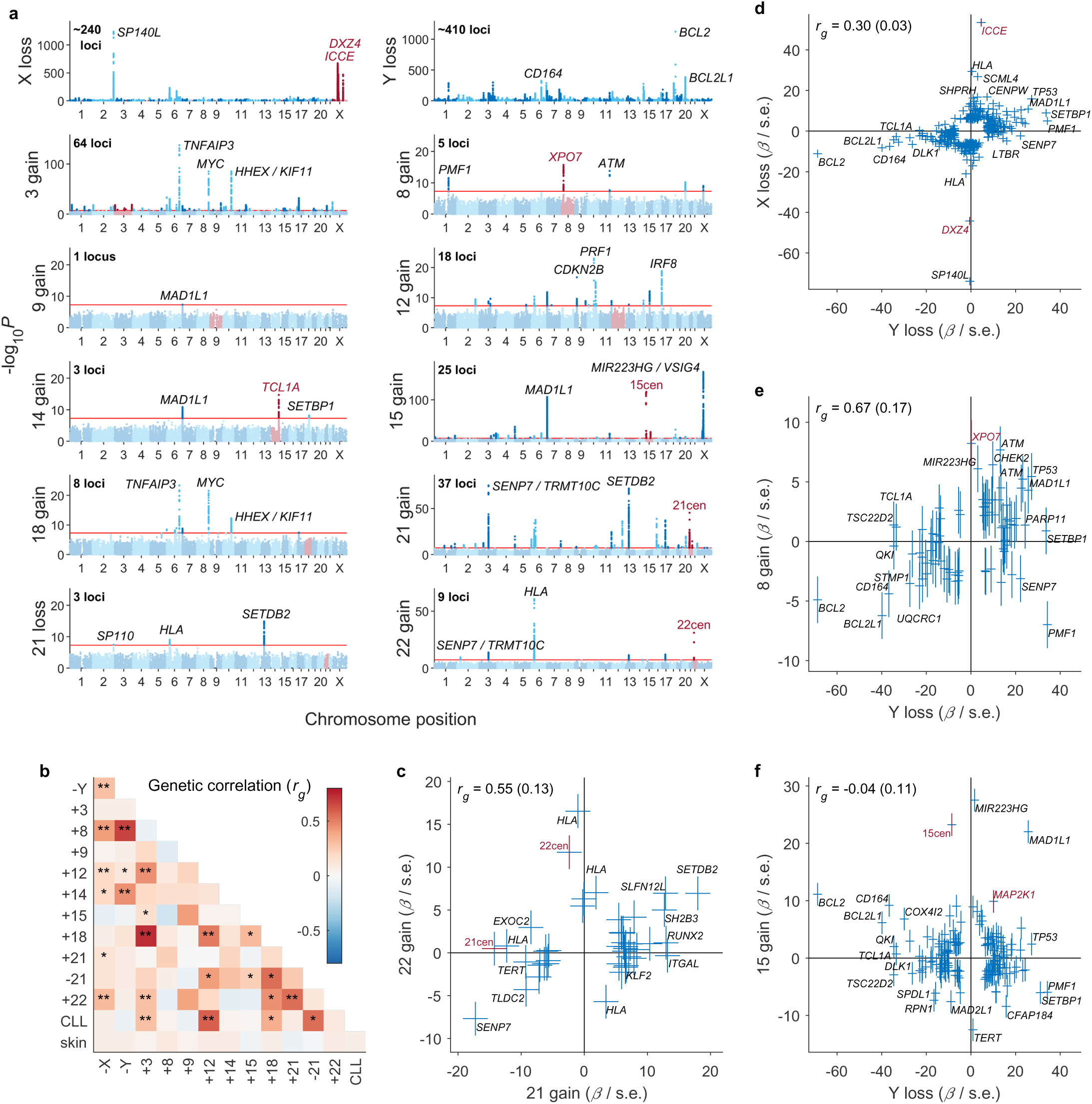
Shared and distinct genetic influences on different mosaic aneuploidies at hundreds of human genomic loci. **a**, Associations of common inherited variants with 12 aneuploidies in blood WGS from the UKB EUR cohort. Bolder colors indicate genome-wide significant associations (*P<*5×10^-8^; red line). Inherited variants on the chromosome lost or gained are colored red. For each aneuploidy, the total number of significant loci is indicated at the top left, and the top three loci are labeled. **b**, Genetic correlations among mosaic aneuploidies, chronic lymphocytic leukemia (CLL), and skin cancer (a negative control). *, *P<*0.05; **, *P<*0.001. **c**–**f**, Comparisons of variant associations (*z* = *β*/s.e. in UKB) with pairs of aneuploidies with varying levels of genetic correlation. Error bars, 95% CIs.

Many of the associations implicated genes involved in mitotic chromosomal segregation, cell-cycle regulation, DNA damage response, apoptosis, and immune cell function, similar to previous GWAS of LOX^13^, LOY^11, 12^, and other forms of clonal hematopoiesis^36–40^. For example, the variants that associated most strongly with autosomal aneuploidies included an eQTL for *MIR223HG* (which hosts microRNA-223, a regulator of hematopoietic differentiation^41^; *P*=4.1×10^-167^ for chromosome 15 gain); a missense variant in the mitotic checkpoint gene *MAD1L1* (*P*=1.9×10^-107^ for chromosome 15 gain^16^); and a haplotype at *TNFAIP3* (which encodes a tumor suppressor for hematopoietic malignancies^42^; *P*=4.2×10^-138^ for chromosome 3 gain; Fig. 2a). Each of these variants also associated with multiple other aneuploidies. For 6 of the 12 aneuploidies, the top three associations also included one or more loci on the affected chromosome (Fig. 2a), suggesting strong allele-specific effects on missegregation (generated by variation at centromeres, *ICCE*, and *DXZ4*; discussed in detail below) or clonal selection on cells with increased dosage of a proliferation-increasing allele (at *XPO7* and *TCL1A*^31, 43^; Supplementary Table 10).

Despite the high-level similarity in the functions of genes implicated by GWAS loci for different hematopoietic aneuploidies, different aneuploidies nonetheless exhibited distinct genetic architectures. Pairwise genetic correlations (*r*_g_) among the 12 aneuploidies tended to be only slightly positive (median *r*_g_=0.15, none significantly negative; Fig. 2b). The few pairs with somewhat larger genetic correlations included gains of chromosomes 21 and 22 (*r*_g_=0.55 (s.e., 0.13); Fig. 2c), LOX and LOY (*r*_g_=0.30 (0.03); Fig. 2d), and LOY and gain of chromosome 8 (*r*_g_=0.67 (0.17); Fig. 2e). Gains of chromosomes 3, 12, and 18 also had high genetic correlations (Fig. 2b), probably owing to the tendency of these aneuploidies to co-occur^14^. In contrast, LOY and gain of chromosome 15 had near-zero genetic correlation, with several variants associating strongly with opposite effects on these two aneuploidies (Fig. 2f). This heterogeneity in effect directions was common among variants in genes likely to affect cell survival or proliferation (Supplementary Fig. 32), suggesting that these variants may confer either selective advantage or disadvantage to different aneuploid genomes.

Gene-based burden tests identified 28 genes for which rare variants predicted to impact protein function associated with LOX, 49 genes for LOY (2 previously reported^44^), and 11 genes for autosomal aneuploidies (FDR∼0.01; Supplementary Fig. 20 and Supplementary Tables 11–13), corroborating the themes from GWAS and confirming target genes at many loci. For LOX, 6 out of 12 LOX-increasing associations involved genes encoding proteins with roles in centromere function: inner kinetochore components CENPO, CENPC, and CENPQ; and the components of the MIS18 complex (MIS18*α*, MIS18*β*, and MIS18BP1), which facilitates centromeric loading of CENP-A^45^. Notable LOX-decreasing associations included protein-altering burden in *SP100* (*P*=4.0×10^-424^), *SP140L*, and *PML*, indicating a key role of PML nuclear bodies in LOX. Rare variants predicted to cause loss of function of two genes in the PAR1 pseudoautosomal region, *GTPBP6* and *DHRSX*, associated with strongly reduced levels of both LOX and LOY (roughly halved cell fractions; Supplementary Tables 11–12), suggesting that leukocytes cannot tolerate biallelic knockout of these genes (produced when LOX or LOY eliminates the functional copy).

### Chromosome-specific effects of kinetochore and centromere variation on missegregation

Some of the most interesting and interpretable genetic associations involved variants for which comparing effect sizes across different aneuploidies allowed us to deduce chromosome-specific vulnerabilities to different failure modes of mitotic segregation. Several such variants affected genes with functions at the kinetochore, the macromolecular protein structure that mechanically links mitotic centromeres and spindle microtubules^46–50^ (Fig. 3a,b). The strongest of these effects were generated by coding and splice-altering variation in *MAD1L1*, *MAD2L1*, *SPDL1*, *ZWILCH*, and *KNTC1* (Fig. 3b and Supplementary Note 3), which facilitate signaling from unattached kinetochores to the spindle assembly checkpoint (SAC) to delay anaphase^51^ (Fig. 3a). The effect sizes of these variants varied across aneuploidies, with minimal effects on LOX and chromosome 21 gain and no detectable effects on gain of chromosome 22 (Fig. 3b). One possible explanation is that chromosome 22 might rarely be the last chromosome to be ready for anaphase; if so, it would be largely unaffected by reduced SAC fidelity.

**Figure 3.**
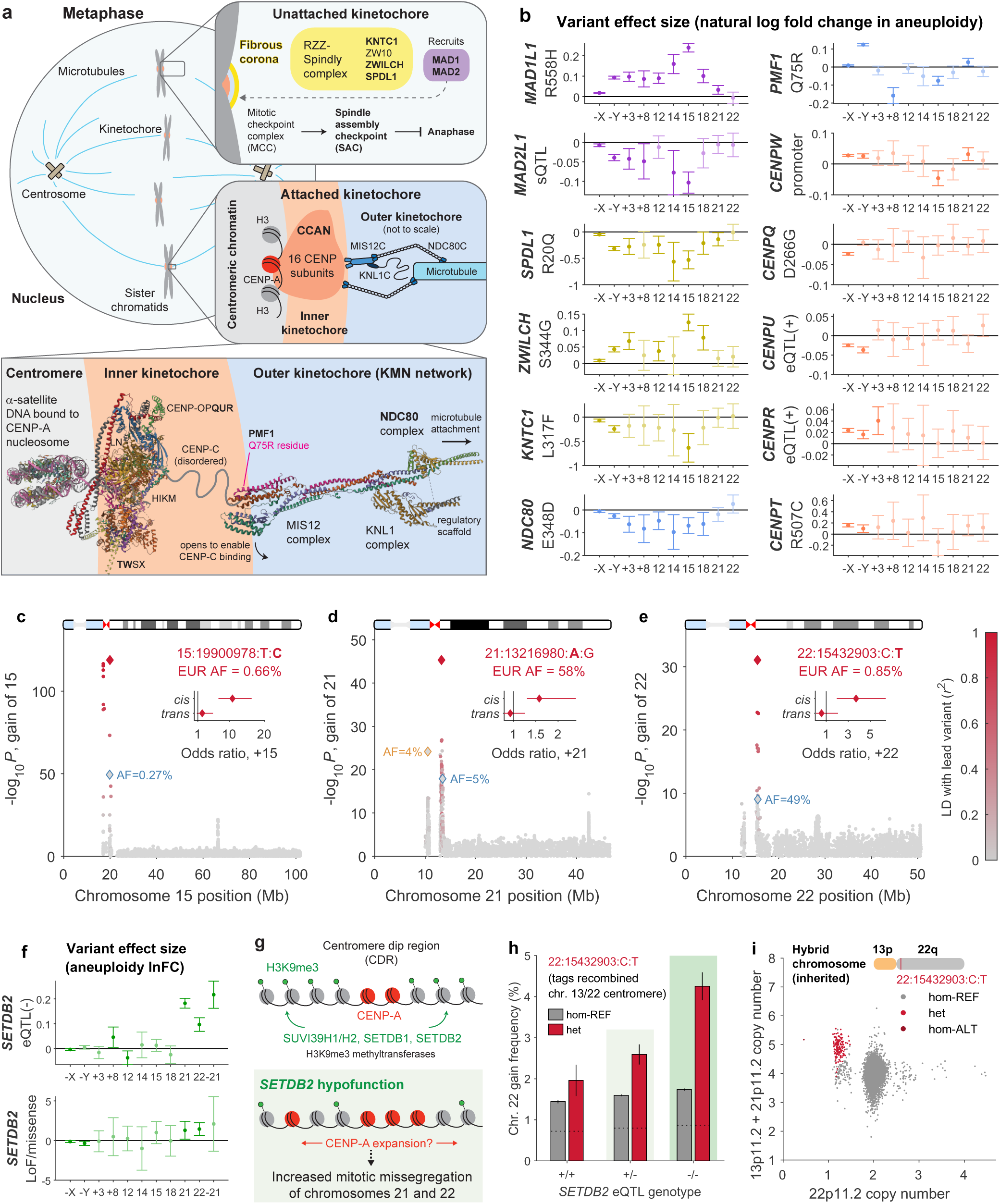
Genetic associations with mosaic aneuploidies identify numerous effects on kinetochore and centromere function. **a**, Schematic of the human kinetochore. The background depicts a cell in metaphase; sister chromatids are held together at centromeres, at which kinetochores are in the process of forming end-on microtubule attachments (shown in detail in zoomed-in boxes). The inner kinetochore (orange shading) is composed of 16-subunit constitutive centromere associated network (CCAN) complexes (PDB 75RS^46^) that bind centromeric *α*-satellite DNA at CENP-A nucleosomes (red). The KMN network, forming the outer kinetochore (blue shading; PDB 8PPR^48^), physically links the CCAN to microtubules (right and bottom boxes). At unattached kinetochores (top-right box), a meshwork of RZZ-Spindly complexes within the fibrous corona^49^ (yellow) recruits MAD1-MAD2 complexes (purple) to signal to the spindle assembly checkpoint (SAC) and delay anaphase. Protein subunits implicated by aneuploidy-associated genetic variants shown in **b** are indicated in bold. **b**, Effect sizes (in the UKB EUR cohort) of aneuploidy-associated inherited variants in kinetochore/SAC genes. Variant effect sizes (y-axis) are in units of natural log fold change (of cell fraction for X and Y loss and of prevalence for autosomal aneuploidies; Methods). Variant labels (y-axis labels) indicate missense mutations or putative functional consequences (eQTL/sQTL, expression/splicing quantitative trait locus; Supplementary Note 3). Variant effects are plotted using the color scheme for kinetochore components in **a**; darker colors indicate effects with 95% CIs that do not overlap 0 (*P<*0.05). Data are presented for 10 of 12 common aneuploidies, excluding gain of chromosome 9 and loss of chromosome 21 owing to insufficient statistical power. **c**–**e**, Associations of inherited variants on chromosomes 15, 21, and 22 with mosaic gains of these chromosomes in the UKB EUR cohort. In each plot, the red diamond indicates the top association; other associations are colored according to linkage disequilibrium (LD) with this variant. Blue and gold outlined diamonds indicate independently associated centromere haplotypes, labeled with their allele frequencies (AF). Inset, odds ratio for association of the lead variant with gain of the homolog containing the variant (*cis*) or the opposite homolog (*trans*) in heterozygous carriers with detectable allelic imbalance^17^. The effect allele is indicated in bold. **f**, Effect sizes of the *SETDB2* expression-reducing eQTL haplotype (13:49518324:CTA:C) (top) and burden of *SETDB2* rare coding variants (bottom) in association analyses of aneuploidies in UKB EUR (units, natural log fold change). **g**, Model for effects of *SETDB2* variation on missegregation of chromosomes 21 and 22: *SETDB2* hypofunction reduces H3K9me3 methyltransferase activity, producing aberrant expansion of the CENP-A regions at which kinetochores form and reducing kinetochore fidelity. **h**, Frequency of mosaic gain of chromosome 22 (mean probabilistic call; Methods) in UKB EUR participants stratified by genotypes of the *SETDB2* eQTL and 22:15432903:C:T; the latter segregates with an inherited ectopic recombination of 22q with 13p. Dashed horizontal lines indicate half of the frequency of mosaic gain of 22 in noncarriers of the tag SNP; in carriers (dark red), the portion of the bar above the dashed line is the inferred frequency of mosaic gain of the hybrid chromosome 22. **i**, Copy-number profiling of UKB participants (downsampled 50-fold) showing near-perfect segregation of 22:15432903:C:T with read-depth deviation consistent with simultaneous gain of 13p (represented by 21p sequence in GRCh38) and loss of 22p. Error bars, 95% CIs. GWAS sample sizes are provided in Supplementary Table 3.

Other variants affected genes encoding components of the kinetochore, again with distinct effects on different aneuploidies. A missense variant in *NDC80* associated with a pattern of effect sizes similar to those of the SAC-modulating variants, whereas a missense variant in *PMF1* (producing a Q75R substitution within the CENP-C binding head of the outer kinetochore^47, 48^) associated with increased loss and gain of Y (Supplementary Fig. 33) but decreased risk of gains of chromosomes 8 and 15 (Fig. 3b). Variants in inner kinetochore genes exhibited similarly heterogenous effects: the *CENPW* promoter variant rs9388486 (PromoterAI^52^ score −0.32) associated with bidirectional effects on different aneuploidies, while a missense variant in *CENPQ* (D266G) appeared to specifically affect LOX (Fig. 3b). These effects suggest subtle differences in how different human centromeres function, perhaps generated by differences in alpha-satellite sequence or methylation^53–56^.

Recent work has also demonstrated extensive allelic variation within centromeres^25, 26^ that influences kinetochore positioning^25, 26, 57^, suggesting the possibility of allelic effects on centromere function^25, 26^. Here, centromeric haplotypes on chromosomes 15, 21, and 22 associated strongly with aneuploidies of these chromosomes (*P*=8.4×10^-32^–1.3×10^-119^ in UKB; Fig. 3c–e). These associations were allele-specific (i.e., phased genotypes associated with gain of the homolog in *cis* but not the homolog in *trans*), suggesting that certain common centromere alleles are indeed more prone to missegregation—sometimes by several-fold (OR=10.8 [6.9–16.3] and 3.7 [2.0–6.4] for *cis* effects on gains of chromosomes 15 and 22; Fig. 3c–e). Moreover, for each of the three chromosomes, multiple centromeric haplotypes associated independently with aneuploidy (Supplementary Table 14). The lead associations passed several robustness checks (Supplementary Note 4), including replication in AoU (Supplementary Table 6) and consistent effects of the chromosome 21 haplotype on losses and gains of chromosome 21 (Supplementary Fig. 34).

Intriguingly, the effect at the chromosome 22 centromere appeared to be driven by an ectopic centromeric recombination of chromosomes 13 and 22 recently observed in 3 out of 415 individuals with centromere assemblies^26^. Analyses of WGS read-depth and chimeric sequences in UKB and 1000 Genomes^58^ demonstrated near-perfect linkage disequilibrium of the risk allele with the ectopic recombination, showing that it is a remarkably large, common inherited structural polymorphism (Fig. 3i, Supplementary Fig. 35, Supplementary Table 15, and Supplementary Note 5). This unique structural variant, which replaced the p-arm of chromosome 22 with that of chromosome 13, probably arose in Southern Europe, where it seems to be most common (1 carrier per ∼30 individuals in this ancestry group; Supplementary Fig. 35a).

Further insight into the mechanism underlying the fourfold increased missegregation rate of the hybrid chromosome 13/22 came from an epistatic interaction with another genetic effect, at *SETDB2* (Fig. 3f–h). A common expression-decreasing *SETDB2* eQTL (the major allele in most populations) associated with increased aneuploidy of chromosomes 21 and 22—generating the strongest and second-strongest genome-wide associations with loss and gain of chromosome 21 (Fig. 2a)—and rare coding variants in *SETDB2* corroborated this effect (Fig. 3f). SETDB2 is an H3K9me3 methyltransferase, and H3K9me3 methylation defines the boundaries of the centromeric CENP-A domains at which kinetochores form^59–61^. This suggests a model in which SETDB2 hypofunction leads to hypomethylation of histones in alpha-satellite DNA and aberrant expansion of CENP-A domains on chromosomes 21 and 22, increasing missegregation (Fig. 3g). This effect was exacerbated in carriers of the recombined chromosome 13/22 (*P*=3.0×10^-10^ for genetic interaction; Fig. 3h), suggesting that the hybrid 13/22 centromere is particularly sensitive to centromeric hypomethylation.

### X-linked *ICCE* and *DXZ4* repeat arrays generate five-fold variation in X loss propensity

Loss of the X chromosome in females appeared to be driven by a different mechanism involving non-centromeric structural variation on chromosome X. Two loci on chromosome X, at *ICCE* and *DXZ4*, harbored outstandingly strong associations with LOX levels that replicated across UKB (Fig. 2a) and AoU (Fig. 4a). *ICCE* and *DXZ4* contain highly polymorphic macrosatellite repeats^62, 63^ (Fig. 4b,c and Supplementary Fig. 36) that form long-range CTCF-mediated intrachromosomal interactions^63^; the *DXZ4* repeat is an essential component of the 3D structure of the inactive X chromosome, bipartitioning it into two superdomains^64–66^. Copy-number variation of these repeat arrays associated much more strongly with LOX levels than nearby SNPs and indels (*P*=2.4×10^-1044^ and 2.3×10^-871^ for *ICCE* and *DXZ4* wild-type copy-number in AoU; Fig. 4a, Supplementary Fig. 37, and Supplementary Note 6).

**Figure 4.**
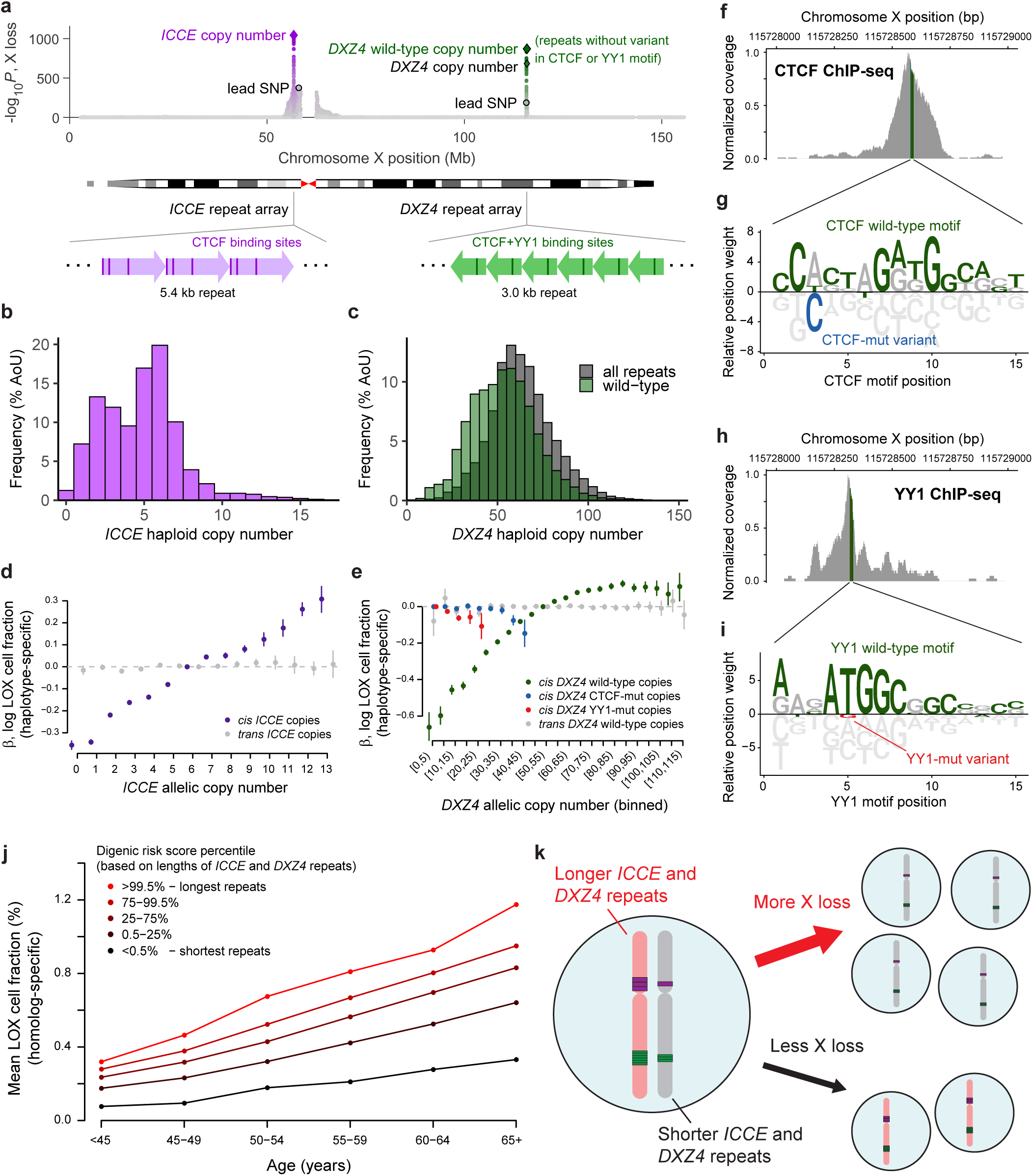
Repeat length and sequence variation in the *ICCE* and *DXZ4* repeat arrays associate with several-fold variation in allele-specific X loss levels. a, Associations of inherited X chromosome variants with X loss levels in AoU (n=193,296 unrelated female participants; Methods). *P*-values are plotted for SNPs, indels, *ICCE* and *DXZ4* repeat copy numbers, and copy numbers of intra-repeat SNP and indel variants. For *DXZ4*, the copy number of repeats with wild-type CTCF and YY1 motifs (i.e., the number of repeat units containing the REF allele in both motifs) is also included. **b**, **c**, Distribution of *ICCE* and *DXZ4* allele lengths (number of repeats) in AoU (n=137,798 haploid males). **d**, **e**, Effect sizes of *ICCE* and *DXZ4* allelic copy numbers on loss of the X chromosome in *cis* (i.e, fraction of cells in which the homolog carrying the allele is lost) and *trans* (i.e, fraction of cells in which the opposite homolog is lost) in UKB EUR-ancestry females. For *DXZ4*, effect sizes are from a joint model including *DXZ4* wild-type repeat copy number as well as the numbers of repeat units containing the common CTCF motif variant and YY1 motif variant, respectively (Supplementary Note 6). **f**, CTCF ChIP-seq alignment coverage in K562 cells at *DXZ4*. ChIP-seq reads were aligned to GRCh38; the plot range shows a 1kb region within a *DXZ4* monomer assembled on the plus strand of GRCh38 (Supplementary Fig. 44), corresponding to positions 247–1,254 of the original monomer^62^. **g**, Visualization of CTCF motif position weight matrix. At each position, bases above the *y*=0 line are enriched in CTCF motifs; bases below are depleted. Base heights are absolute values of log_2_ probabilities relative to geometric means across the four bases. Green letters indicate the reference *DXZ4* sequence; the common PSV within the CTCF motif is indicated in blue (chrX:115728581:T:C, which replaces a T with a strongly disfavored C). **h**, YY1 ChIP-seq coverage in K562 cells in the same 1kb region of *DXZ4*. **i**, YY1 position weight matrix, with the reference *DXZ4* sequence in green and the common PSV within the CTCF motif in red (chrX:115728322:T:G, which replaces a conserved T with a G). **j**, Mean homolog-specific X loss cell fractions among X chromosomes stratified by X loss digenic risk score (based on *ICCE* and *DXZ4* wild-type repeat lengths; Methods) and participant age in UKB (EUR-ancestry females). **k**, Inferred model: X chromosomes with longer *ICCE* and *DXZ4* repeats have increased risk of missegregation during mitosis, generating increasing levels of X loss with age that skew toward loss of the homolog with longer repeats. Error bars, 95% CIs.

At *ICCE*, common alleles containing 0–13 copies of a core 5.4-kb repeat unit formed an allelic series of increasing effect sizes on haplotype-resolved LOX cell fractions (Fig. 4d). These associations were specific to loss of the X chromosome in *cis*, consistent with a direct effect of longer *ICCE* repeats on increased missegregation; *ICCE* allele length did not associate with loss of the X chromosome in *trans* (Fig. 4d). *ICCE* copy number variation almost fully accounted for associations of SNPs and indels at the locus: residual associations centered at the X chromosome centromere, suggesting an independent effect of centromeric variation on LOX (Supplementary Fig. 38a,b).

At the much longer *DXZ4* array (10–120 copies of a 3.0-kb monomer), a more complex association pattern provided clues about mechanism. Longer *DXZ4* repeats also associated with increased loss of the X chromosome in *cis*, but conditional association analyses identified two paralogous sequence variants (PSVs) within the *DXZ4* array that appeared to disrupt this effect (Fig. 4e and Supplementary Fig. 38c–f). These two PSVs modified bases within CTCF and YY1 binding sites (∼260bp apart within each *DXZ4* repeat unit; Fig. 4f,h) and were predicted to reduce binding of CTCF and YY1 (Fig. 4g,i); analyses of CTCF ChIP-seq data corroborated this prediction (Supplementary Fig. 38g). Partitioning *DXZ4* allelic copy numbers into numbers of repeats carrying the CTCF motif-disrupting PSV, the YY1 motif-disrupting PSV, or neither PSV (wild-type repeats) showed that only the copy number of *DXZ4* wild-type repeats associated with increased X loss; copy numbers of repeats containing the CTCF or YY1 motif-disrupting PSV associated with near-zero, slightly negative effects (Fig. 4e). This suggests that increased CTCF and YY1 binding at *DXZ4* reduces segregation fidelity of the inactive X chromosome.

Combining these two genetic effects into a digenic risk score for LOX (accounting for allelic series nonlinearity; Fig. 4e) showed that common combinations of *ICCE* and *DXZ4* alleles associated with a 4-fold range in the propensities of X chromosomes to be lost (comparing haploid chromosomes in the top and bottom 0.5% of digenic scores; Fig. 4j). The effects at *ICCE* and *DXZ4* contributed additively in a model of LOX levels (log-transformed cell fraction of the homolog in *cis*), and each locus explained ∼5% of the heritability of age-adjusted LOX levels (the GWAS phenotype for LOX, combined across each individual’s two homologs). Taken together, these results support a model in which increased lengths of the CTCF-binding *ICCE* and *DXZ4* repeats that create the superstructure of the inactive X chromosome also generate vulnerabilities to missegregation (Fig. 4k), perhaps by influencing the nuclear positioning of the inactive X chromosome, its replication speed or timing, or cohesion between sister chromatids.

### Allele-specific effects of X chromosome variation on recurrent X loss and clonal selection

Previous studies of genetic effects on X loss have observed numerous variants across the X chromosome at which LOX exhibits an “allelic shift”: that is, in females heterozygous for a variant, the homolog carrying one allele is lost more often than the other homolog is lost^13, 14^. Puzzlingly, the same variants exhibited weaker or no association with overall risk of X loss (in GWAS of LOX case status)^13, 14^. Haplotype-resolved analyses of LOX provided insights into these mysteries. First, most female UKB participants exhibited losses of each of their two homologous X chromosomes (presumably in distinct cell populations; Fig. 5a–c). This observation provides further evidence that LOX is driven by recurrent missegregation in many cells, and it explains the reduced power of GWAS of allelic imbalance-based LOX phenotypes (which measure the difference rather than the sum of the fractions of cells with loss of each homolog). However, allelic shift analyses of WGS read-depth-based LOX phenotypes still identified far more X chromosomal associations than GWAS of LOX levels (Supplementary Fig. 39).

**Figure 5.**
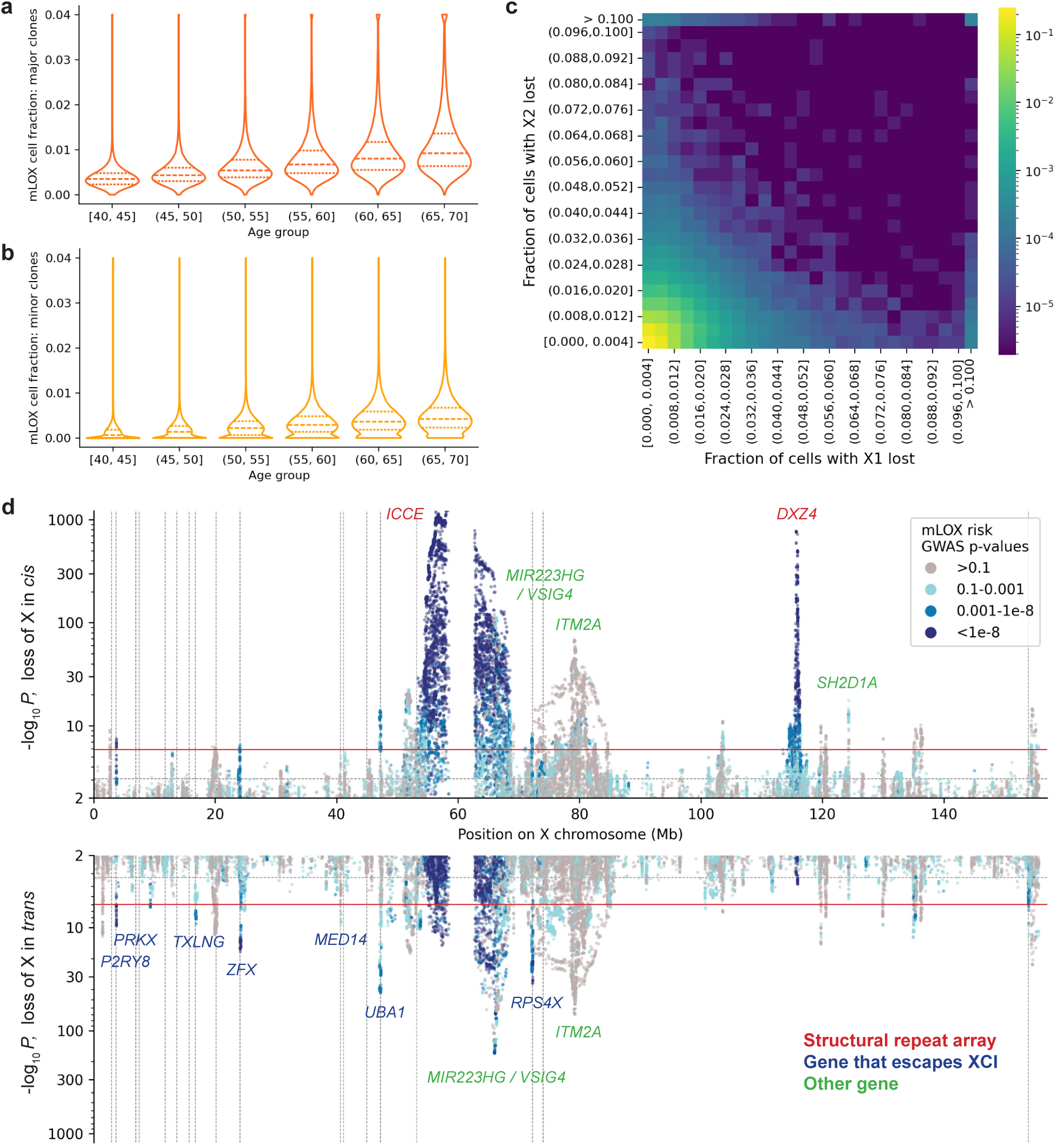
Female loss of X is polyclonal and associates with allele-specific effects of inherited X chromosome variation. **a**, Distribution of the total cell fraction of the major clones (i.e., fraction of cells having lost the homolog lost in more of an individual’s leukocytes) in each 5-year age group in UKB. **b**, Analogous distribution for minor clones. **c**, Joint distribution of the total cell fractions of the two types of clones (loss of X1 and X2, ordered arbitrarily). **d**, Associations of inherited X chromosome variants with loss of the X chromosome in *cis* (top) and *trans* (bottom), analyzing log-transformed, clipped cell fractions. Variants are colored by the extent to which they associate with overall X loss: some variants associate strongly with opposing effects on cell fractions of X loss involving the homologs in *cis* versus *trans*, such that they do not associate with overall X loss (gray points).

To further explore this phenomenon, we tested X chromosome variants for allele-specific associations with loss of the homolog in *cis* or *trans*. Variants at *ICCE* and *DXZ4* associated hundreds of times more strongly with LOX in *cis* than in *trans*, as expected, but at most other loci, variants associated with opposite-direction effects (of similar magnitudes) on fractions of cells with LOX in *cis* and *trans* (Fig. 5d and Supplementary Table 16). Such associations could in theory arise from alleles that bias X chromosome inactivation (XCI), as previously proposed^13^; however, we did not observe allelic bias in XCI in female OneK1K^67^ participants (although statistical power was limited; Supplementary Fig. 40 and Supplementary Note 7). These single-cell analyses did suggest that LOX always affects the inactive X chromosome^6^ (Supplementary Fig. 41).

Interestingly, variants at several X chromosome loci deviated somewhat from having reciprocal associations with LOX in *cis* and *trans*, such that they also associated with overall LOX (GWAS *P<*0.001), and most of these loci were centered at or near genes that escape X inactivation (*P2RY8*, *PRKX*, *TXLNG*, *ZFX*, *UBA1*, and *RPS4X*; Fig. 5d). Variants at these loci tended to associate more strongly with LOX in *trans* than in *cis*. We speculate that this could reflect incomplete escape from X inactivation^68^, such that alleles have a larger effect on relative clonal fitness when they are on the active, retained X chromosome (i.e., LOX occurs in *trans*) than when they are partially expressed on the inactive homolog (affected by LOX in *cis*).

Collectively, these results suggest that the allelic skews observed in X loss originate from multiple genetic effects: a primary effect of *ICCE* and *DXZ4* copy number variation on missegregation in *cis*, a secondary role of escape-gene variants that affect cell fitness, and other still-unexplained effects that require further study.

### Limited effects of hematopoietic sex chromosome aneuploidies on health

The large numbers of genetic risk loci for LOX and LOY provided an opportunity to evaluate the causality of epidemiological associations between hematopoietic sex chromosome aneuploidies and clinical outcomes^5, 7–10, 13^ using Mendelian randomization (MR)^69^. Bidirectional MR analyses testing for relationships of LOX and LOY with 9–10 health outcomes (38 directional relationships) did not identify strong evidence of any causal relationships: no relationship consistently reached significance (*P<*0.05/38) across four MR methods (Fig. 6a, Supplementary Fig. 42 and Supplementary Tables 17 and 18). Analyses of polygenic risk scores (PRS) for LOY levels in UKB (using 5-fold cross-validation to obtain out-of-sample predictions) likewise indicated limited causal effects on male health. The out-of-sample PRS predicted 23.4% of variance in age-adjusted LOY levels (Fig. 6b), but it showed no association with most health outcomes (Fig. 6c) and was not predictive of longevity phenotypes (Fig. 6d,e). Only prostate cancer exhibited a significant association with LOY PRS (*P*=3.9×10^-4^; Fig. 6d), with some support from MR (*P*=0.002 to 2×10^-5^ across methods; Fig. 6a); however, the effect size was modest (8% higher prostate cancer prevalence among male UKB participants in the top versus bottom LOY PRS quartile).

We also wondered whether the large aneuploidy-associated germline structural variants we identified—at *ICCE*, *DXZ4*, and the hybrid chromosome 13/22—might affect health outcomes. Analyses of 3,447 clinical phenotypes in AoU identified just one significant association, of *DXZ4* repeat length with reduced risk of hemorrhoids (*P*=1.8×10^-9^; Supplementary Fig. 43a and Supplementary Note 8). This association was female-specific (*P*=1.1×10^-9^ in females, *P*=0.22 in males) and was the strongest genetic association with hemorrhoids in the entire genome in females (Supplementary Fig. 43b), replicating in female AoU participants not initially genotyped (*P*=0.014). Interestingly, the association appeared to be driven by total *DXZ4* copy number (rather than wild-type copy number; *P*=1.1×10^-9^ versus *P*=1.3×10^-8^), suggesting a distinct molecular effect of *DXZ4* length variation (e.g., on expression of a nearby gene) unrelated to CTCF binding and LOX. Consistent with an independent mechanism, LOX levels did not associate with hemorrhoids (*P*=0.30), nor did *ICCE* copy number (*P*=0.98). This finding motivates further study of *DXZ4* length variation and its effects on X chromosome segregation and on female health.

**Figure 6.**
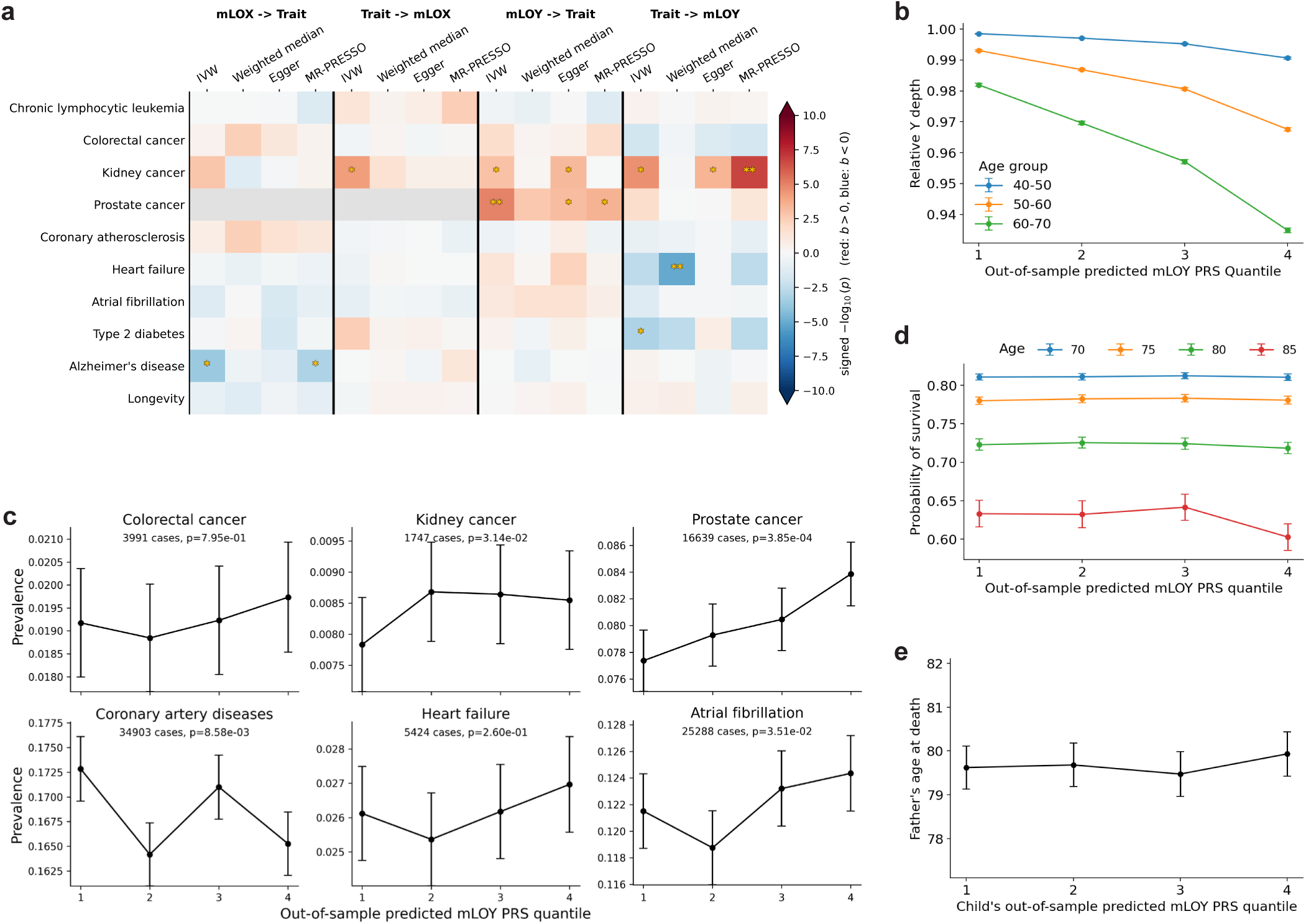
Limited effects of hematopoietic sex chromosome aneuploidies on health. **a**, *P*-values from Mendelian randomization analyses evaluating causality of associations between X loss and health outcomes (left two columns) and between Y loss and health outcomes (right two columns). Four MR models were run: inverse-variance weighted (IVW), weighted median, MR Egger, and MR-PRESSO. Data are presented for health outcomes highlighted in previous studies of hematopoietic X and Y loss. Darker colors indicate more significant *P*-values; *, *P<*0.05/38; **, *P<*0.001/38. **b**, Normalized Y chromosome WGS coverage in EUR-ancestry male UKB participants stratified by age and Y loss polygenic risk score (PRS) quartile (using PRS values computed out-of-sample; Methods). Individuals of higher age and polygenic risk tend to have lower Y depth, indicating loss of Y. Centers, means; error bars, 95% CIs. **c**, Disease prevalences in each out-of-sample LOY PRS quartile. **d**, Fractions of participants living to age 70, 75, 80, and 85 years in each out-of-sample LOY PRS quartile (restricted to participants old enough at recruitment to assess survival to each age; Methods). **e**, Father’s age at death in each out-of-sample LOY PRS quartile (treating a UKB participant’s LOY PRS as an approximation of the PRS of the participant’s father). Centers, means; error bars, 95% CIs.

## Discussion

This work shows that many inherited genetic variants strongly influence the likelihood of blood genomes to acquire aneuploidies during aging. These insights were enabled by new computational methods for detecting low-level aneuploidy from blood DNA sequencing data—turning population biobank cohorts^27, 28^ into a hematopoietic aneuploidy data set of unprecedented scale (hundreds of thousands of sex chromosome aneuploidies; ∼70,000 autosomal aneuploidies). This resource provided an opportunity to observe how germline variation shapes naturally occurring aneuploidy within healthy blood cell populations.

Several genetic effects appeared to impact centromere and kinetochore function in ways that generate vulnerabilities of specific human chromosomes to mitotic missegregation. These results build on recent breakthroughs establishing kinetochore structure (using microscopy methods)^46–49^ and characterizing centromeric sequence variation^24–26^ and CENP-A epigenetic regulation^54, 55, 59–61^ (using long-read sequencing technologies). Here, GWAS of aneuploidies detected from abundant short-read WGS data identified several centromeric haplotypes that confer risk of missegregation, including a recently discovered ectopic centromeric recombination^26^. Identifying the causal variants and molecular mechanisms underlying the other effects (which did not appear to involve hybrid centromeres; Supplementary Note 5) will require future work, perhaps analyzing larger reference panels of centromere assemblies or measuring centromere variation in short-read data sets^70^ using pangenome approaches^71^. Such analyses may help explain why these effects appear to be particularly strong for acrocentric chromosomes (15, 21, and 22).

Other genetic effects centered at genes with roles in cellular survival and proliferation, as observed for LOX and LOY^11–13^. Many questions about these effects also remain unresolved, including how the same polymorphisms can confer opposite fitness effects to different aneuploid genomes, and whether these fitness effects are specific to certain blood cell types. More fundamentally, the extent to which aneuploidies themselves confer a proliferative advantage to leukocytes^72^ is still unclear; here, age trajectories across population cross-sections suggested exponential growth of some aneuploidies (including gain of 15 and LOY) but not others (loss of 21 and LOX). The striking difference between the trajectories of LOX in females compared to LOY in males, both of which result in X0 karyotype, is an intriguing mystery.

Lastly, key questions remain about the health consequences of hematopoietic aneuploidies, and more generally, the genetic effects that promote them. Our Mendelian randomization analyses suggested a limited role of hematopoietic LOX and LOY in disease, but a more definitive determination will require other methods or yet-larger GWAS cohorts. Beyond blood, genetic effects on chromosome segregation may affect cell division in other tissues, potentially influencing risk of cancer or meiotic error (though meiosis involves largely distinct mechanisms and genetics^73, 74^). These sets of questions, prompted here by genetic insights into aneuploidies in biobank cohorts, may be fruitful areas for future study.

## Methods

### Ethics

This research complies with all relevant ethical regulations. The study protocol (NHSR-4981) was determined to be not human subject research by the Broad Institute Office of Research Subject Protection as all data analyzed were previously collected and de-identified.

### UKB data set

UKB is a prospective cohort of volunteer participants aged 40–70 years at recruitment between 2006 and 2010^75^. Participants attended assessment centers at which they provided biosamples, answered touchscreen questionnaires, and participated in nurse interviews. Following initial assessment, health outcome data has accrued from linkage with UK national health registries; additionally, a subset of participants returned for one or more repeat assessments in subsequent years.

Short-read WGS data were generated for 490,414 UKB participants as previously described^27^; briefly, blood-derived DNA was sequenced using Illumina NovaSeq 6000 machines to an average coverage of 32.5×, and DRAGEN 3.7.8 was used to align 151-bp paired-end reads to GRCh38 and perform SNP and indel calling. Blood samples used for WGS had been acquired at initial assessment for 99.6% of sequenced individuals, so in analyses of age in UKB, we used age at initial assessment; averaged across the cohort, this underestimated age at blood draw by ∼1 week.

Because UKB participants could withdraw from the study at any time, analyses reported in this manuscript have small discrepancies in participant counts, reflecting the numbers of available (non-withdrawn) participants at the time of each analysis.

### AoU data set

AoU is a longitudinal cohort of volunteer participants aged 18–90+ years at recruitment, beginning in 2018^76^. Most participants provided biosamples and data from surveys and electronic health records (EHR); data on subsequent health outcomes were obtained from EHR linkage.

Short-read WGS data were generated from blood- or saliva-derived DNA for 414,817 individuals in the AoU v8 release (365,918 from blood). Sequencing was performed as previously described^28^; briefly, DNA was sequenced using Illumina NovaSeq 6000 machines, and reads were aligned to GRCh38 with Illumina DRAGEN Bio-IT Platform Germline Pipeline 3.4.12. Samples were sequenced to an average coverage of 37.9×.

### Analyzing mosaic aneuploidy using chromosome-wide WGS read-depth

The crux of our approach to detecting and quantifying mosaic aneuploidies was to compute highly accurate estimates of chromosome-wide sequencing coverage, from which mosaic aneuploidies could be ascertained as deviations from expected coverage. This conceptually simple approach of directly evaluating chromosomal abundances from genomic data was used in earlier studies that estimated LOY levels from SNP-array probe intensities^5, 7, 30, 31^. However, for diploid chromosomes, measurements of relative allelic representation at heterozygous sites (B-allele frequency; BAF) were observed to be less prone to technical artifacts and more suitable for calling mCAs^3, 4, 6^. Subsequent studies found that incorporating phase information further increased the power of BAF-based analysis^14, 77^, to the extent that even for LOY, BAF-based analysis (measuring allelic imbalance between only the PAR1 pseudoautosomal regions of chromosomes X and Y) became preferred^11, 12, 29^. In this spirit, we recently developed an allelic imbalance-based method for mCA analysis using phased BAF from WGS data^17^. This method included a module for denoising WGS read-depth measurements, which we used to assign copy-number state to BAF-based mCA calls^17^.

To our surprise, we subsequently discovered that the denoised WGS read-depth measurements were sufficiently accurate that, with further methodological development (described here), they could be used in a standalone—and much more powerful—read-depth-based method for detecting chromosomal aneuploidies, capable of identifying aneuploidies present in 0.2–0.4% of cells (Supplementary Tables 1–2). Moreover, beyond the increased detection sensitivity, this read-depth-based approach provided two additional advantages when analyzing LOX and LOY. First, it correctly handled polyclonality in female LOX (which we found commonly involves cell populations in which each of the two homologous X chromosomes is lost). Secondly, it enabled estimating LOX levels in all females and LOY levels in all males (not only those reaching thresholds for calling LOX/LOY), facilitating interpretation and improving GWAS power.

In the following sections, we detail the methods that we used to denoise chromosome-wide WGS read-depth measurements and to compute mosaic aneuploidy phenotypes optimized for GWAS. Supplementary Note 1 reports several benchmarking and validation analyses comparing this approach to BAF-based analyses using SNP-array and WGS data.

### Denoising WGS read-depth

Short-read WGS data from a single human DNA sample sequenced at high coverage (∼30× coverage by ∼150-bp paired-end reads) typically contains read pairs derived from ∼5–25 million DNA fragments per diploid chromosome (as human chromosomes range from ∼50–250 million base pairs). Most of these read pairs can be mapped uniquely to their chromosome of origin. A mosaic aneuploidy in which a chromosome is gained or lost in a small fraction of cells *α* causes the chromosome to be over- or underrepresented by a factor of 1 ± *α/*2 for diploid chromosomes (1 − *α* for LOY). In theory, if the expected number of DNA fragments attributable to the chromosome (in the absence of aneuploidy) were known—i.e., if chromosome-wide read-depth measurements were perfectly calibrated—then aneuploidy could be detected and quantified directly from the observed chromosomal coverage. This approach would have a relative error equal to the inverse square root of the number of sampled DNA fragments (the “shot noise” proportion), i.e., 0.0002–0.00045. For diploid chromosomes, knowing the relative (observed/expected) WGS read-depth of the chromosome to a precision (s.e.) of 0.0002–0.0045 would then allow estimating the aneuploid cell fraction *α* to a precision (s.e.) of 0.0004–0.0009. This represents a theoretical limit on the informativeness of WGS for aneuploidy estimation: in an ideal scenario of perfect calibration, direct quantification of chromosomal abundance from fragment counts is the optimal approach (and is much more precise than allelic imbalance-based analysis).

In practice, multiple biological and technical confounders (including germline copy-number variation, blood cell type proportions, and sequencing batch effects) generate variation in sequencing coverage that far exceeds shot noise from sampling DNA fragments. However, we found here that by carefully filtering and denoising WGS read counts, we could reduce noise in chromosome-wide read-depth to a level near the theoretical limit (s.e. within ∼1.5-fold; Supplementary Tables 1–2 and Supplementary Note 1). The key steps of the pipeline are as previously described^17^: (1) defining a genome mask that eliminates low-complexity, low-mappability, and common CNV regions; (2) profiling read-depth at 1-kb resolution and computing empirical GC-bias profiles^78^; (3) identifying and masking individual-level germline CNVs; and (4) correcting for remaining systematic biases by removing top principal components (PCs) of genome-wide read-depth profiles computed in sex-specific sets of reference individuals.

Here, to better handle pervasive LOX and LOY, we made one important modification to the approach used to adjust for read-depth PCs. Previously, we observed that beyond a certain number of PCs (12 PCs for chromosome X in females, 24 PCs for chromosome Y in males), the read-depth PCs began to noticeably capture LOX or LOY itself (even in reference sets restricted to the youngest UKB participants), such that removing subsequent PCs removed signal of LOX and LOY. We previously ameliorated this problem by imposing an orthogonality constraint that effectively disabled PC-adjustment of chromosome-wide read-depths for X and Y beyond these points^17^; however, this was suboptimal because earlier PCs still captured a small amount of LOX and LOY signal, attenuating estimates of LOX and LOY levels, and later PCs were no longer useful for normalizing X and Y depth. Here, we devised a different approach that circumvented this issue entirely: after computing genome-wide GC-adjusted, CNV-masked read-depth profiles (at ∼50-kb resolution), we collapsed the read-depth profiles within the X and Y chromosomes into 25 bins per sex chromosome before computing PCs. This produced read-depth vectors containing 49,716 elements, of which 49,666 represented ∼50-kb autosomal bins and 50 elements represented the sex chromosomes. Because the sex chromosomes only contributed 0.1% of the coordinates of these collapsed read-depth vectors, common but low-level LOX and LOY now contributed a negligible amount of overall read-depth variance, such that top read-depth PCs no longer captured LOX or LOY (while still capturing ways in which genome-wide “genomic waves” in read-depth profiles were informative of technical variation in chromosome-wide X and Y coverage, enabling precise calibration). The modified approach was particularly helpful for denoising X-chromosome coverage; it was helpful but less crucial for the Y chromosome, for which shot noise was relatively high (imposing a theoretical minimum s.e. of 0.0013 from ∼600,000 read pairs aligned to 12 Mb of the haploid Y chromosome that remained after excluding the masked pseudoautosomal regions, ampliconic repeats, and heterochromatin regions^79^).

We computed read-depth PCs within sex-specific reference panels containing younger individuals least likely to have mosaic aneuploidies: 7,684 female and 6,852 male UKB participants (selected as before^17^) and 7,976 female and 3,742 male AoU participants aged 18–22 years at blood draw. This produced four sets of 50 PCs and corresponding baseline read-depth vectors^17^. The PCs computed in UKB differed somewhat from those computed in AoU (reflecting cohort-specific technical variation), such that adjusting for within-cohort PCs was more effective than computing PCs in only one cohort and reusing them to denoise read-depth profiles from the other.

We used the PCs to denoise read-depth profiles (with X and Y collapsed to 25 bins each) for 490,089 UKB participants and 350,764 AoU participants with blood-derived WGS data (restricting to the EUR, AMR, and AFR genetic ancestry groups in the AoU v8 release^28^). For each individual, we determined genetic sex and applied the corresponding baseline normalization and PC adjustment (along with a final normalization setting the median of autosomal median coverages to 1)^17^.

The final output of this procedure (in each of UKB and AoU) was a matrix of denoised, normalized chromosome-wide median coverages: 24 values corresponding to chromosomes 1–22, X, and Y, each typically very close to 1 (except for Y chromosomal coverage in females, which was typically very close to 0). Additionally, we computed an overall read-depth noise metric for each sample (specifically, the median read-depth deviation of ∼5-Mb autosomal chunks from their respective autosomal median coverages); this noise metric was useful for subsequent outlier removal.

### Mosaic X and Y loss phenotypes

To define sample sets for LOX and LOY analyses, we started by assigning genetic sex based on Y chromosome ploidy (from denoised Y chromosome coverage). We used a custom pipeline rather than using DRAGEN sex calls because ∼1,000 AoU participants with evidence of high-cell-fraction LOY or LOX had DRAGEN sex calls of NA; these individuals had very low Y chromosome or X chromosome coverage and were much older than average.

We classified individuals with estimated Y chromosome ploidy *>*0.05 as male and those with estimated Y chromosome ploidy *<*0.0015 as female, handling constitutional sex chromosome aneuploidies later. A small fraction of samples (1–2% of each cohort) had estimated Y chromosome ploidy between 0.0015 and 0.05. Such values indicated potential contamination of female blood samples with small amounts of male blood DNA, which would create the appearance of X loss (because males have only one X chromosome). We therefore excluded these samples from our LOX/LOY analyses, ensuring that female samples included in LOX analyses had male contamination levels of at most ∼0.1%.

We estimated LOX cell fractions in females and LOY cell fractions in males from X- and Y-chromosome-wide coverage. We verified that sex chromosome coverage profiles showed minimal intrachromosomal heterogeneity (Supplementary Fig. 5), consistent with nearly all mosaic sex chromosome aneuploidies being losses of whole sex chromosomes^5, 6, 14, 30^. In AoU, we computed female LOX cell fractions as 2×(1–X_depth_) and male LOY cell fractions as (1–Y_depth_), where X_depth_ and Y_depth_ denote the denoised, normalized chromosome-wide coverages of X and Y relative to their baselines in reference individuals, as described above. In UKB, the younger individuals that we included in the reference panel (40–41 years of age) were old enough to have modest amounts of LOX and LOY (Supplementary Fig. 3), such that the baseline coverages of X and Y in UKB were slightly miscalibrated. To adjust for this slight miscalibration, we applied offsets corresponding to the mean deviations in X and Y depth observed in 40–41-year-old AoU female and male participants: we computed LOX and LOY cell fractions in UKB as 2×(1.002–X_depth_) and (1.003–Y_depth_), respectively.

Some LOX and LOY cell fraction estimates were slightly negative, as expected given remaining noise (Supplementary Figs. 1–2). Some of the more-negative estimates could reflect constitutional or mosaic gains of X or Y; such events are much rarer^5, 6, 14, 80^, but to minimize their representation in the data set, we excluded individuals with estimated cell fractions of LOX (for females) or LOY (for males) below –0.1, corresponding to full or *>*10% mosaic XXX (UKB n=143, AoU n=154) and full or *>*10% mosaic XYY (UKB n=180, AoU n=155). We did not attempt to distinguish and exclude non-mosaic cases of Turner syndrome (X0), as these are very rare^80^. Lastly, in UKB, we restricted to individuals with SNP-array genotyping data^75^ (which we previously used to assign genetic ancestries^81^), producing our final cohorts for analyzing LOX (UKB n=259,088, AoU n=206,847) and LOY (UKB n=221,857, AoU n=138,703).

We also defined log-transformed LOX and LOY cell fraction phenotypes, which were more suitable for GWAS because cells with LOX or LOY sometimes undergo clonal expansion, causing the distributions of LOX and LOY cell fractions to be right-skewed (Supplementary Figs. 1–2). In AoU, we computed:

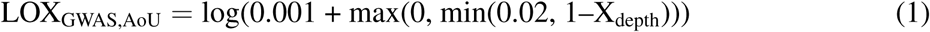

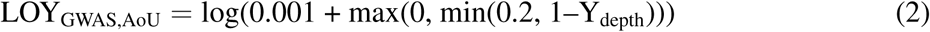

corresponding to clipping LOX and LOY cell fractions to the ranges 0–4% and 0–20%, respectively, and adding constant offsets of 0.2% and 0.1% before taking the natural logarithm.

In UKB, we used slightly different formulas because of the minimum age of 40 years, which had led to the need to recalibrate the baseline coverages of X and Y. Here, instead of clipping at 0 and adding a constant offset, we simply clipped from below at the offset value:

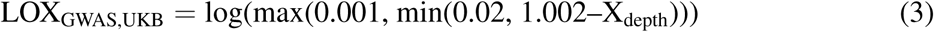

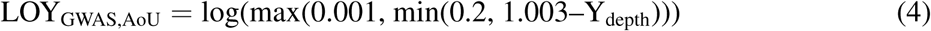

corresponding to clipping LOX and LOY cell fractions to the ranges 0.2–4% and 0.1–20%, respectively, before taking the natural logarithm. Clipping at small positive values was sensible in UKB given that LOX and LOY were typically present at low levels in individuals over age 40. We selected the constants used in the above formulas to maximize association strengths of lead variants from previous GWAS^11–13^.

### Mosaic autosomal aneuploidy phenotypes

Denoised chromosome-level WGS coverage also enabled powerful detection of autosomal mosaic aneuploidies, but deriving phenotypes from this information required different techniques owing to the lower frequency and greater heterogeneity of autosomal mCAs compared to LOX and LOY. First, we restricted analyses to 10 autosomal mCAs (gains on chromosomes 3, 8, 9, 12, 14, 15, 18, 21, and 22 and losses on chromosome 21) that have previously been observed at higher frequencies, and for which most mCAs are whole-chromosome events^17, 20^. Second, we dropped outlier samples with overall read-depth noise in the top 0.5% of each cohort (leaving UKB EUR n=451,924, AoU EUR+AMR+AFR n=348,956). This prevented false-positive-prone, noisy samples from diluting analyses of rarer autosomal aneuploidies; it was not necessary for LOX and LOY, which are very common. Third, rather than analyzing cell fractions (or log-transformed cell fractions) of mosaic aneuploidies, we generated binary and probabilistic calls of autosomal mosaic aneuploidies. Analyzing cell fractions was sensible for LOX and LOY, which were present at measurable levels in most individuals (Supplementary Fig. 15), but autosomal aneuploidy levels were in the noise for most individuals.

We first defined binary calls of autosomal aneuploidies based on deviations of chromosome-level read-depth beyond certain thresholds. To control FDR at ∼0.05, we needed to identify read-depth thresholds that varied across chromosomes (as smaller chromosomes had noisier read-depth measurements, and aneuploidy rates varied across chromosomes). To do so, we empirically estimated the standard error *σ* of denoised, normalized read-depth for each chromosome as 1.4826 times the median absolute deviation (MAD) of chromosome-level read-depth estimates in younger individuals (in AoU, age *<*22 years). We then assumed a Gaussian null distribution *N* (1*, σ*^2^) for chromosome-level read-depth and determined the read-depth deviation threshold yielding an FDR of 0.05 (using the Benjamini-Hochberg method). This produced binary calls that exhibited strong age-enrichment (Supplementary Figs. 13–14), confirming adequate false positive control. To facilitate comparing aneuploidy rates across different chromosomes, we also computed binary calls using read-depth deviation thresholds that corresponded to a fixed cell fraction threshold (*>*0.4%, which provided adequate false positive control for all chromosomes), and we applied the same methods to compute binary calls for LOX and LOY (Fig. 1h and Supplementary Table 2).

To increase power in GWAS of autosomal mosaic aneuploidies, we also defined probabilistic-call phenotypes that incorporated information from chromosome-level read-depth, age, and sex. The rationale for this approach was twofold: (1) to use a participant’s age and sex when evaluating the likelihood of a borderline read-depth deviation to reflect aneuploidy rather than noise (Supplementary Fig. 13); and (2) to generate a quantitative spectrum of phenotype values capturing uncertainty in detection (Supplementary Fig. 17). To implement this approach, we first stratified each cohort by sex. Within each sex, for each of the 10 aneuploidy types, we did the following:

1. **Fit a model estimating a prior on the odds of having the aneuploidy** as a function of age. To do so, we estimated the standard error *σ* of chromosome-level read-depth as 1.4826 times the MAD across individuals aged ≤41 years, and we fit a logistic model regressing case status (defined as a *>*4*σ* read-depth deviation in the appropriate direction) on log(age) and log(age)^2^, clipping age to be ≤80 years.
2. **For each individual, compute a read-depth *z*-score** *z* by dividing the observed read-depth deviation by *σ*. If the read-depth deviation was in the opposite direction of the aneuploidy, we set *z* to zero if |*z*|≤2; if the read-depth deviation was *>*2*σ* in the opposite direction, we set the phenotype to missing to avoid potential confounding from cases of the opposite aneuploidy (e.g., confounding of the chromosome 21 loss phenotype from non-cases being enriched for chromosome 21 gain).
3. **Estimate the posterior log odds of aneuploidy** as logit(*p*) = *z*^2^/2 + (prior log odds). The estimated posterior probability *p* of having the aneuploidy could then be obtained by applying the inverse logit function.
4. **Clip posterior probabilities** *<***0.05 to zero** to minimize potential confounding from subtle read-depth variation unrelated to aneuploidy.

A few aspects of this procedure are theoretically loose (e.g., the choice of the 4*σ* case threshold when fitting the prior and the flat prior on *z* among cases); however, empirically, the phenotype transformation achieved the desired effects and was robust to adjustments to the model that we considered.

### GWAS of mosaic aneuploidy phenotypes

We ran a total of 34 GWAS of 12 mosaic aneuploidy phenotypes in up to 4 cohorts defined by genetic ancestry groupings (UKB EUR, AoU EUR, AoU AMR, and AoU AFR). We analyzed all 12 aneuploidies (LOX, LOY and 10 autosomal aneuploidies) in the UKB EUR and AoU EUR cohorts. We analyzed subsets of aneuploidies in the smaller AoU AMR and AFR cohorts owing to limited power to analyze rarer aneuploidies: we analyzed LOX, LOY, and chromosome 21 gain and loss in both the AoU AMR and AoU AFR cohorts, and we also analyzed chromosome 15 and 22 gain in AoU AMR.

We ran GWAS using linear mixed models (for LOX and LOY) or linear regression (for autosomal aneuploidies) implemented in BOLT-LMM^82^ v2.5, including age, age^2^, smoking status, genetic PCs (20 for UKB, 16 for AoU), and sex (for autosomal aneuploidies) as covariates. We tested SNP and indel variants previously imputed with INFO score *>*0.3 using the TOPMed R2 reference panel^83, 84^ or called directly from WGS with *<*10% missingness. For female LOX and male LOY (log-transformed, clipped cell fraction phenotypes; Supplementary Fig. 15), we tested all variants with minor allele frequency (MAF) *>*0.1%, constituting 14–23 million variants per cohort. For autosomal aneuploidies (probabilistic calls on a 0–1 scale; Supplementary Fig. 17), we applied more stringent MAF filters (0.35% to 3.5%, depending on aneuploidy prevalence; Supplementary Table 3) to avoid test statistic inflation^85^, leaving 7–11 million variants. Details of the association analyses are provided in Supplementary Note 2, and GWAS sample sizes are provided in Supplementary Table 3. Follow-up analyses testing for allele-specific associations of inherited variants with gain of the homolog in *cis* (i.e., the haploid chromosome containing the variant) are also described in Supplementary Note 2, as are gene-based burden tests of aneuploidy phenotypes.

### Identification and annotation of lead variants at GWAS loci

We used the canonical genome-wide significance threshold *P<*5×10^-8^ to identify significant associations, reasoning that most of the mosaic aneuploidy GWAS we conducted found several hits at *P<*5×10^-8^ (Supplementary Figs. 20–30, Supplementary Tables 4–6), such that the FDR was well-controlled. To define loci and identify index variants for GWAS of LOX and LOY in UKB and AoU, we first identified a subset of pairwise-independent associations by iterating through variants in descending order of association strength and retaining a variant only if it remained significantly associated (*P<*5×10^-8^) in conditional analysis with each previously-retained nearby variant^86^. We then defined loci and index variants by iterating through the remaining variants (again in descending order of association strength) and retaining a variant only if it was *>*500 kb away from any previously retained variant.

We used a simpler approach to define loci and index variants for GWAS of autosomal aneuploidy phenotypes. These GWAS identified fewer loci (up to 64 loci, compared to hundreds for LOX and LOY) harboring associations that were generally not exceptionally strong (10 strongest *P*-values: 4.8×10^-46^–4.1×10^-167^). The higher MAF thresholds (0.35% to 3.5%) that we imposed for these GWAS also limited the range of linkage disequilibrium (LD) with tagging variants. Consequently, defining loci based on requiring a genetic distance of *>*1 cM between lead variants largely sufficed. Upon inspecting loci defined in this way, we found only two instances in AoU in which this approach retained nearby loci that did not represent independent associations (the Duffy locus, at which African American cohorts exhibit long-range admixture LD, and the chromosome 15 centromere, which may have inaccurate genetic map coordinates). We therefore post-processed the initial lists of lead variants from AoU using the pairwise conditional significance criterion^86^ to obtain final sets of lead variants.

We annotated index variants with nearby genes using GENCODE^87^ v39 definitions for protein-coding genes, long non-coding RNAs, and microRNAs. We also annotated index variants as expression quantitative trait loci (eQTLs) or splicing quantitative trait loci (sQTLs) using GTEx v10 summary statistics^88^. Finally, if any nearby protein-altering variants had association strengths nearly as strong as the index variant, we annotated this information as well (Supplementary Tables 4–9). Specifically, we extracted annotations for protein-coding variants with high or moderate Variant Effect Predictor (VEP) consequence^89^ from the gnomAD v4.1.0 exomes release^90^, and we reported variants with association strengths *>*90% of the nearest index variant. In supplementary Manhattan plots (Supplementary Figs. 20–30), to improve readability, we annotated lead variants with the nearest protein-coding gene (within 500 kb) in GENCODE v47. Additionally, to facilitate plotting, we downsampled non-significant variants with *P>*1×10^-4^ by a factor of *P* / 1×10^-4^.

### Rescaling effect sizes to units of natural log fold change

To facilitate interpretation and comparison of variant effect sizes (*β*), we rescaled *β* to the scale of natural log fold change when studying effect sizes (Fig. 3b,f and Supplementary Figs. 31–32). For LOX and LOY, the GWAS phenotypes were natural log-transformed, clipped cell fractions, such that *β* was already on this scale (up to modest attenuation from clipping, which affected a minority of samples; Supplementary Fig. 15). For autosomal aneuploidies, the GWAS phenotype was a probabilistic call, so we transformed *β* to the scale of natural log fold change in phenotype prevalence:

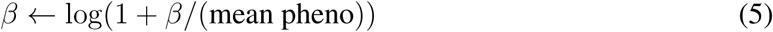

These units are not fully harmonized because fold change in autosomal aneuploidy prevalence has a different meaning from fold change in LOX and LOY cell fractions, but we found them helpful for understanding absolute effect sizes and for comparing the cross-aneuploidy effect-size profiles of different inherited variants (Fig. 3b,f and Supplementary Fig. 32); being able to compare effect sizes of variants across multiple related phenotypes mitigated some of the complexities of interpreting GWAS findings^91^.

### Estimation of heritability and genetic correlations

We estimated the heritability of female LOX and male LOY levels (log-transformed, clipped cell fractions) in UKB using two approaches: variance components analysis in unrelated individuals and sibling-pair analysis. For both approaches, to reduce potential confounding from shared environmental effects, we restricted analyses to EUR-ancestry individuals and excluded current smokers^30, 31^. We also restricted analyses to individuals aged ≥60 years, by which point the large majority of individuals had measurable levels of sex chromosome loss (Supplementary Fig. 2).

To estimate SNP-heritability^33^ (*h*_g_^2^), we first pruned to unrelated sets of individuals by dropping one individual per second-degree or closer relationship pair (kinship *>*0.0884 computed by UKB^75^). We estimated *h*_g_^2^ using BOLT-REML^92^ (as implemented in BOLT-LMM v2.5), including smoking status and 20 genetic PCs as covariates. This yielded LOX *h*_g_^2^ ≈ 35% (s.e., 1%; n=95,817 females) and LOY *h*_g_^2^ ≈ 44% (1%; n=85,092 males) in the age-controlled subcohort.

We estimated broad-sense heritability (*H*^2^) as twice the sib-sib correlation (using sisters aged ≥60 years for LOX and brothers aged ≥60 years for LOY, dropping monozygotic twins and adjusting log-transformed, clipped cell fractions for age, age^2^, and smoking status). This gave LOX *H*^2^ ≈ 52% [45%–60%] and LOY *H*^2^ ≈ 62% [52%–71%].

The estimates from these two approaches were reasonably concordant, as SNP-heritability is a lower bound on narrow-sense heritability (as it does not capture effects of untagged variants), whereas doubling the sib-sib correlation gives an upper bound on broad-sense heritability (as it is susceptible to inflation from shared environmental effects, and here, siblings of the same sex are required to inherit the same sex chromosome from their father). Both approaches gave heritability estimates much higher than previously obtained (e.g., *h*_g_^2^ ≈ 10% for LOX^13, 14^ and 31.7% for LOY^11^), probably reflecting greatly reduced noise in the LOX and LOY phenotypes analyzed here.

To estimate genetic correlations between aneuploidy phenotypes in UKB, we ran LDSC^93^ (v2.0.0) on pairs of GWAS summary statistics. We also estimated genetic correlations with CLL and skin cancer (as a control phenotype) using summary statistics from meta-analyses of FinnGen R12, MVP, and UKB (gs://finngen-public-data-r12/meta analysis/mvp ukbb/summary stats).

### Haplotype-resolved analyses of LOX in UKB

Haplotype-resolved analysis of LOX in females was possible in UKB because phased haplotypes could be statistically estimated in this cohort (representing ∼1% of the UK population) with chromosome-scale accuracy. We used SHAPEIT5^94^ `phase_common` to phase WGS-derived genotypes (called using the GraphTyper pipeline^27^) for a scaffold of X chromosome variants from the SNP array used by OneK1K^67^ that passed QC (Supplementary Note 7). This facilitated subsequent reference-based phasing of OneK1K samples, and phasing a scaffold of SNP-array sites can also improve long-range phase accuracy^95^.

We evaluated phasing accuracy in 4,213 female UKB participants for whom BAF-based analysis^17^ identified LOX with *>*2% cell fraction, such that phase switch errors disrupting long-range phase were easily detectable. Among these individuals, 83.6% had no detectable phase switch errors, 12.8% had only one switch, and 3.6% had two or more switches. Phase switch errors were rarer in regions with lower recombination rates, as expected.

To resolve read-depth-derived estimates of total LOX cell fractions into haplotype-specific estimates for loss of each homologous X chromosome, we computed the relative abundance of the two homologs by summing phased allelic depths across chrX:68.5–115 Mb (GRCh38), after which we calculated LOX cell fractions for haplotype 1 and haplotype 2 that summed to the total LOX cell fraction and produced the observed relative abundance of non-lost X homologs:

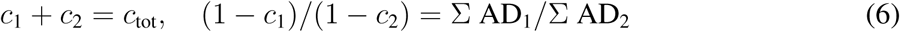

The haplotype-specific LOX estimates *c*_1_ and *c*_2_ were sometimes slightly negative, which we allowed to avoid introducing upward bias. We selected the region chrX:68.5–115 Mb for two reasons: (1) because it is relatively recombination-cold (spanning only 35 cM, i.e., 18% of the recombination length of the X chromosome), such that we expected ∼97% of female UKB participants to have no switch errors within it; and (2) to eliminate any possibility of confounding from effects of variation at *DXZ4* and *ICCE*. We computed phased allelic depths from common SNPs (MAF*>*1%) throughout this region using our deduping and CNV-masking pipeline^17^, mapping previously computed haplotypes^15^ onto the SHAPEIT5-phased scaffold haplotypes. We adjusted allelic depths for variant-specific reference bias^17^, which we estimated in heterozygotes among 20,133 female UKB participants with negligible LOX (age *<*45 years and minimal read-depth deviation).

To test common variants across the X chromosome for association with loss of the homolog in *cis* or *trans*, we also generated haplotype-resolved LOX cell fraction phenotypes specific to each 1-cM window on the X chromosome. For each 1-cM window (defined using the genetic maps available at https://github.com/odelaneau/GLIMPSE/tree/master/maps/genetic maps.b38), we estimated fractions of cells with loss of each haplotype in the same way as above, but using allelic depths of common SNPs located *>*1 cM and *<*30 cM from the focal window (i.e., 58 cM that remained within a 61-cM region centered at the 1-cM window after excluding the middle 3 cM). Similar to GWAS of LOX, we restricted to EUR-ancestry UKB participants and tested haploid genotypes for association with log(max(0.001, min(0.02, haplotype-specific LOX cell fraction))) for the homolog in *cis* or *trans*. We performed linear regression adjusting for age, age^2^, smoking status, 20 genetic PCs, and an LOX PRS (computed using autosomal lead variants from the previous LOX GWAS^13^) as covariates.

### Digenic risk score for susceptibility of X chromosomes to loss

We used the following procedure to build a digenic risk score using phased repeat-length genotypes at *ICCE* and *DXZ4* to estimate the susceptibility of a haploid X chromosome to loss. Based on the allelic series observed at *ICCE* and *DXZ4* (Fig. 4d,e), we fit a linear model regressing haploid LOX levels (log(max(0.001, LOX cell fraction))) on *ICCE* copy number in *cis* and natural log-transformed *DXZ4* wild-type copy number in *cis*, clipped to at least 10 copies. We also included age, age^2^, smoking status, and 20 genetic PCs as covariates. We fit the regression on haplotypes of female EUR-ancestry UKB participants. This resulted in the model 0.05265×(*ICCE* hapCN) + 0.3327×(log(max(10, *DXZ4* wild-type hapCN))), where hapCN denotes the copy number of the haplotype in *cis*.

Supplementary Note 6 describes the analyses that supported this model: copy-number and PSV genotyping and phasing at *ICCE* and *DXZ4*, analyses of LOX associations and allelic series, and analyses of CTCF and YY1 binding in *DXZ4*.

### Mendelian randomization analyses

We performed bidirectional MR to assess causality of associations between hematopoietic sex chromosome loss and health outcomes. We selected 10 health outcomes for analysis, taking into consideration reported epidemiological associations^5, 7–10, 13^ and availability of GWAS summary statistics (Supplementary Table 17). We used sex chromosome loss as either the exposure or the outcome, and we tested LOY with all 10 clinical traits and LOX with 9 traits (excluding prostate cancer, which is male-specific). We ran four MR methods (inverse-variance weighted, weighted median, MR-Egger^96^, and MR-PRESSO^97^) implemented in genal^98^ v1.6 to evaluate sensitivity to different assumptions of these models.

For each exposure, we selected instrumental variables from the set of genetic variants significantly associated with the exposure (*P<*5×10^-8^). We restricted to common variants (MAF*>*0.01 in 1000 Genomes EUR^99^) and performed LD clumping to *R*^2^*<*0.01 using plink^100^ v2.0. For MR-PRESSO, which is more computationally intensive, we capped the number of instrumental variables at the 500 most significant.

### Polygenic scores for LOY

We computed out-of-sample PRS for LOY using five-fold cross-validation within the UKB male cohort. For each cross-validation fold in turn, we restricted the cohort to the other folds and ran BOLT-LMM on log-transformed, clipped LOY cell fractions (using the `--predBetasFile` option to generate PRS weights for SNP-array variants) with age, age^2^, smoking status, and 20 genetic PCs as covariates. We used these PRS coefficients to compute scores for the left-out fold.

### Disease and longevity analyses

We studied the relationships of LOY PRS with binary disease phenotypes (“all-source” phenotypes curated as previously described^81^) and longevity phenotypes in UKB from two sources. First, we analyzed death registry data to compute proportions (per LOY PRS quartile) of participants surviving to age 70, 75, 80, and 85. For each age threshold *T*, we limited analyses to male participants born at least *T* years before 2025. Second, to obtain direct estimates of longevity (rather than survival probabilities), we analyzed paternal ages at death reported by UKB participants during repeat imaging visits (“Instance 3”) in 2019 or later. We analyzed data from 10,691 non-sibling UKB participants, comprising 9,994 individuals who reported “father’s age at death” (field 1807) and 697 individuals who reported ages of fathers still living; for the latter group, we imputed age at death as the conditional expectation based on the empirical distribution from the 9,994 reported ages at death. We analyzed these data by using the child’s LOY PRS as a proxy for the father’s LOY risk, which also allowed including female UKB participants in these analyses.

## Code availability

Code for denoising WGS read-depth profiles and computing aneuploidy phenotypes will be publicly released after completion of peer review.

## Data Availability

Genomic and phenotypic data analyzed in this study are available by application to UK Biobank (http://www.ukbiobank.ac.uk/) and All of Us (https://allofus.nih.gov/). Full summary statistics from GWAS of mosaic aneuploidy phenotypes in UKB and AoU will be made available at the NHGRI-EBI GWAS Catalog after completion of peer review. Individual-level mosaic aneuploidy phenotypes in AoU v8 will be released in a community workspace, and aneuploidy phenotypes in UKB will be returned when a mechanism for release on the UKB-RAP platform becomes available.

## Acknowledgements

We thank B. Handsaker and G. Genovese for helpful discussions. This research was conducted using the UK Biobank Resource under application number 40709. Y.S. was supported by funding from the Eric and Wendy Schmidt Center at the Broad Institute of MIT and Harvard. N.K. was supported by US NIH fellowship F31 DE034283. D.T. was supported by US NIH training grant T32 HG002295. P.-R.L. was supported by US NIH grants R56 HG012698 and R01 HG013110 and a Burroughs Wellcome Fund Career Award at the Scientific Interfaces. This research was supported by the NIH Common Fund, through the Office of Strategic Coordination and/or Office of the NIH Director under award UM1 DA058230. The funders had no role in study design, data collection and analysis, decision to publish or preparation of the manuscript. The content is solely the responsibility of the authors and does not necessarily represent the official views of the National Institutes of Health. Computational analyses were performed on the UKB Research Analysis Platform, the All of Us Researcher Workbench, and the O2 High Performance Compute Cluster supported by the Research Computing Group at Harvard Medical School. The All of Us Research Program is supported by the NIH, Office of the Director: Regional Medical Centers: 1 OT2 OD026549; 1 OT2 OD026554; 1 OT2 OD026557; 1 OT2 OD026556; 1 OT2 OD026550; 1 OT2 OD026552; 1 OT2 OD026553; 1 OT2 OD026548; 1 OT2 OD026551; 1 OT2 OD026555; IAA no. AOD 16037; Federally Qualified Health Centers: HHSN 263201600085U; Data and Research Center:5 U2C OD023196; Biobank: 1 U24 OD023121; The Participant Center: U24 OD023176; Participant Technology Systems Center: 1 U24 OD023163; Communications and Engagement: 3 OT2 OD023205; 3 OT2 OD023206; and Community Partners: 1 OT2 OD025277; 3 OT2 OD025315; 1 OT2 OD025337; and 1 OT2 OD025276. In addition, the All of Us Research Program would not be possible without the partnership of its participants.

## Author contributions

Y.S., N.K., and P.-R.L. conceptualized the project and performed analyses. D.T. generated allelic depth data and gene-level burden masks. Y.S. and P-R.L. drafted the manuscript. All authors reviewed and edited the manuscript.

## References

1. Jacobs, P. A., Court Brown, W. & Doll, R. Distribution of human chromosome counts in relation to age. Nature 191, 1178–1180 (1961).

2. Jacobs, P. A., Brunton, M., Court Brown, W., Doll, R. & Goldstein, H. Change of human chromosome count distributions with age: evidence for a sex difference. Nature 197, 1080–1081 (1963).

3. Jacobs, K. B. et al. Detectable clonal mosaicism and its relationship to aging and cancer. Nature Genetics 44, 651–658 (2012).

4. Laurie, C. C. et al. Detectable clonal mosaicism from birth to old age and its relationship to cancer. Nature Genetics 44, 642–650 (2012).

5. Forsberg, L. A. et al. Mosaic loss of chromosome Y in peripheral blood is associated with shorter survival and higher risk of cancer. Nature Genetics 46, 624–628 (2014).

6. Machiela, M. J. et al. Female chromosome X mosaicism is age-related and preferentially affects the inactivated X chromosome. Nature Communications 7, 11843 (2016).

7. Dumanski, J. P. et al. Mosaic loss of chromosome Y in blood is associated with Alzheimer disease. The American Journal of Human Genetics 98, 1208–1219 (2016).

8. Sano, S. et al. Hematopoietic loss of Y chromosome leads to cardiac fibrosis and heart failure mortality. Science 377, 292–297 (2022).

9. Sato, G. et al. Genetic regulation across germline and somatic variation on the Y chromosome contributes to type 2 diabetes. Nature Medicine 32, 894–905 (2026).

10. Bruhn-Olszewska, B. et al. The effects of loss of Y chromosome on male health. Nature Reviews Genetics 26, 320–335 (2025).

11. Thompson, D. J. et al. Genetic predisposition to mosaic Y chromosome loss in blood. Nature 575, 652–657 (2019).

12. Francis, M., et al. Multi-ancestry genome-wide association meta-analysis of mosaic loss of chromosome Y in the Million Veteran Program identifies 240 novel loci. medRxiv (2024).

13. Liu, A. et al. Genetic drivers and cellular selection of female mosaic X chromosome loss. Nature 631, 134–141 (2024).

14. Loh, P.-R. et al. Insights into clonal haematopoiesis from 8,342 mosaic chromosomal alterations. Nature 559, 350–355 (2018).

15. Loh, P.-R., Genovese, G. & McCarroll, S. A. Monogenic and polygenic inheritance become instruments for clonal selection. Nature 584, 136–141 (2020).

16. Terao, C. et al. Chromosomal alterations among age-related haematopoietic clones in Japan. Nature 584, 130–135 (2020).

17. Tang, D., Kamitaki, N., Mukamel, R. E., Rubinacci, S. & Loh, P.-R. Patterns and drivers of 43,617 mosaic chromosomal alterations in blood. Nature Genetics 58, 1308–1319 (2026).

18. Zhao, K. et al. Genetic drivers and clinical consequences of mosaic chromosomal alterations in 1 million individuals. medRxiv (2025).

19. Mitchell, E. et al. Clonal dynamics of haematopoiesis across the human lifespan. Nature 606, 343–350 (2022).

20. Jakubek, Y. A. et al. Mosaic chromosomal alterations in blood across ancestries using whole-genome sequencing. Nature Genetics 55, 1912–1919 (2023).

21. Worrall, J. T. et al. Non-random mis-segregation of human chromosomes. Cell Reports 23, 3366–3380 (2018).

22. Dumont, M. et al. Human chromosome-specific aneuploidy is influenced by DNA-dependent centromeric features. EMBO Journal 39, e102924 (2020).

23. Klaasen, S. J. et al. Nuclear chromosome locations dictate segregation error frequencies. Nature 607, 604–609 (2022).

24. Logsdon, G. A. et al. The variation and evolution of complete human centromeres. Nature 629, 136–145 (2024).

25. Logsdon, G. A. et al. Complex genetic variation in nearly complete human genomes. Nature 644, 430–441 (2025).

26. Gao, S. et al. A global view of human centromere variation and evolution. Nature (2026).

27. UK Biobank Whole-Genome Sequencing Consortium et al. Whole-genome sequencing of 490,640 UK Biobank participants. Nature 645, 692–701 (2025).

28. The All of Us Research Program Genomics Investigators. Genomic data in the All of Us Research Program. Nature 627, 340–346 (2024).

29. Jakubek, Y. A. et al. Genomic and phenotypic correlates of mosaic loss of chromosome Y in blood. The American Journal of Human Genetics 112, 276–290 (2025).

30. Dumanski, J. P. et al. Smoking is associated with mosaic loss of chromosome Y. Science 347, 81–83 (2015).

31. Zhou, W. et al. Mosaic loss of chromosome Y is associated with common variation near *TCL1A*. Nature Genetics 48, 563–568 (2016).

32. Lin, S.-H. et al. Incident disease associations with mosaic chromosomal alterations on autosomes, X and Y chromosomes: insights from a phenome-wide association study in the UK Biobank. Cell & Bioscience 11, 143 (2021).

33. Yang, J. et al. Common SNPs explain a large proportion of the heritability for human height. Nature Genetics 42, 565–569 (2010).

34. Ge, T., Chen, C.-Y., Neale, B. M., Sabuncu, M. R. & Smoller, J. W. Phenome-wide heritability analysis of the UK Biobank. PLoS Genetics 13, e1006711 (2017).

35. Sacristan, C. et al. Vertebrate centromeres in mitosis are functionally bipartite structures stabilized by cohesin. Cell 187, 3006–3023 (2024).

36. Bick, A. G. et al. Inherited causes of clonal haematopoiesis in 97,691 whole genomes. Nature 586, 763–768 (2020).

37. Kar, S. P. et al. Genome-wide analyses of 200,453 individuals yield new insights into the causes and consequences of clonal hematopoiesis. Nature Genetics 54, 1155–1166 (2022).

38. Kessler, M. D. et al. Common and rare variant associations with clonal haematopoiesis phenotypes. Nature 612, 301–309 (2022).

39. Stacey, S. N. et al. Genetics and epidemiology of mutational barcode-defined clonal hematopoiesis. Nature Genetics 55, 2149–2159 (2023).

40. Poeschla, M. & Sankaran, V. G. Genetic influences on haematopoiesis. Nature Reviews Genetics 27, 603–621 (2026).

41. Chen, C.-Z., Li, L., Lodish, H. F. & Bartel, D. P. MicroRNAs modulate hematopoietic lineage differentiation. Science 303, 83–86 (2004).

42. Schmitz, R., et al. *TNFAIP3* (A20) is a tumor suppressor gene in Hodgkin lymphoma and primary mediastinal B cell lymphoma. Journal of Experimental Medicine 206, 981–989 (2009).

43. Weinstock, J. S. et al. Aberrant activation of *TCL1A* promotes stem cell expansion in clonal haematopoiesis. Nature 616, 755–763 (2023).

44. Zhao, Y., et al. *GIGYF1* loss of function is associated with clonal mosaicism and adverse metabolic health. Nature Communications 12, 4178 (2021).

45. Fujita, Y. et al. Priming of centromere for CENP-A recruitment by human hMis18α, hMis18β, and M18BP1. Developmental Cell 12, 17–30 (2007).

46. Yatskevich, S. et al. Structure of the human inner kinetochore bound to a centromeric CENP-A nucleosome. Science 376, 844–852 (2022).

47. Polley, S. et al. Structure of the human KMN complex and implications for regulation of its assembly. Nature Structural & Molecular Biology 31, 861–873 (2024).

48. Yatskevich, S., Yang, J., Bellini, D., Zhang, Z. & Barford, D. Structure of the human outer kinetochore KMN network complex. Nature Structural & Molecular Biology 31, 874–883 (2024).

49. Kixmoeller, K., Tarasovetc, E. V., Mer, E., Chang, Y.-W. & Black, B. E. Centromeric chromatin clearings demarcate the site of kinetochore formation. Cell 188, 1280–1296 (2025).

50. Ariyoshi, M. & Fukagawa, T. An updated view of the kinetochore architecture. Trends in Genetics 39, 941–953 (2023).

51. McAinsh, A. D. & Kops, G. J. Principles and dynamics of spindle assembly checkpoint signalling. Nature Reviews Molecular Cell Biology 24, 543–559 (2023).

52. Jaganathan, K. et al. Predicting expression-altering promoter mutations with deep learning. Science 389, eads7373 (2025).

53. Waye, J. S. & Willard, H. F. Nucleotide sequence heterogeneity of alpha satellite repetitive DNA: a survey of alphoid sequences from different human chromosomes. Nucleic Acids Research 15, 7549–7569 (1987).

54. Altemose, N. et al. Complete genomic and epigenetic maps of human centromeres. Science 376, eabl4178 (2022).

55. Gershman, A. et al. Epigenetic patterns in a complete human genome. Science 376, eabj5089 (2022).

56. Jaggi, K. E., Hoyt, S. J., O’Neill, R. J. & Sullivan, B. A. A genomic and epigenomic view of human centromeres. Nature Reviews Genetics (2026).

57. Aldrup-MacDonald, M. E., Kuo, M. E., Sullivan, L. L., Chew, K. & Sullivan, B. A. Genomic variation within alpha satellite DNA influences centromere location on human chromosomes with metastable epialleles. Genome Research 26, 1301–1311 (2016).

58. Byrska-Bishop, M. et al. High-coverage whole-genome sequencing of the expanded 1000 Genomes Project cohort including 602 trios. Cell 185, 3426–3440 (2022).

59. Salinas-Luypaert, C. et al. DNA methylation influences human centromere positioning and function. Nature Genetics 57, 2509–2521 (2025).

60. Carty, B. L. et al. Heterochromatin boundaries maintain centromere position, size and number. Nature Structural & Molecular Biology 33, 220–234 (2026).

61. Xu, Y., et al. Haplotype-resolved centromeric chromatin organization from a complete diploid human genome. bioRxiv (2026).

62. Giacalone, J., Friedes, J. & Francke, U. A novel GC-rich human macrosatellite VNTR in Xq24 is differentially methylated on active and inactive X chromosomes. Nature Genetics 1, 137–143 (1992).

63. Horakova, A. H., Moseley, S. C., McLaughlin, C. R., Tremblay, D. C. & Chadwick, B. P. The macrosatellite *DXZ4* mediates CTCF-dependent long-range intrachromosomal interactions on the human inactive X chromosome. Human Molecular Genetics 21, 4367–4377 (2012).

64. Rao, S. S. et al. A 3D map of the human genome at kilobase resolution reveals principles of chromatin looping. Cell 159, 1665–1680 (2014).

65. Darrow, E. M. et al. Deletion of *DXZ4* on the human inactive X chromosome alters higher-order genome architecture. Proceedings of the National Academy of Sciences 113, E4504–E4512 (2016).

66. Miga, K. H. et al. Telomere-to-telomere assembly of a complete human X chromosome. Nature 585, 79–84 (2020).

67. Yazar, S. et al. Single-cell eQTL mapping identifies cell type–specific genetic control of autoimmune disease. Science 376, eabf3041 (2022).

68. Tukiainen, T. et al. Landscape of X chromosome inactivation across human tissues. Nature 550, 244–248 (2017).

69. Sanderson, E. et al. Mendelian randomization. Nature Reviews Methods Primers 2, 6 (2022).

70. Hain, C., Rausch, T., Consortium, H. G. S. V., Consortium, H. P. R. & Korbel, J. O. HOROSCOPE: Decoding human centromere architecture from short reads using k-mer signatures. bioRxiv (2026).

71. Lu, S. et al. Pangenome-based human genome analysis improves trait association and genomic prediction. bioRxiv 2026–07 (2026).

72. Watson, C. J. & Blundell, J. R. Mutation rates and fitness consequences of mosaic chromo-somal alterations in blood. Nature Genetics 55, 1677–1685 (2023).

73. Halldorsson, B. V. et al. Characterizing mutagenic effects of recombination through a sequence-level genetic map. Science 363, eaau1043 (2019).

74. Carioscia, S. A. et al. Common variation in meiosis genes shapes human recombination and aneuploidy. Nature 651, 146–153 (2026).

75. Bycroft, C. et al. The UK Biobank resource with deep phenotyping and genomic data. Nature 562, 203–209 (2018).

76. All of Us Research Program Investigators. The “All of Us” research program. New England Journal of Medicine 381, 668–676 (2019).

77. Vattathil, S. & Scheet, P. Extensive hidden genomic mosaicism revealed in normal tissue. The American Journal of Human Genetics 98, 571–578 (2016).

78. Handsaker, R. E. et al. Large multiallelic copy number variations in humans. Nature Genetics 47, 296–303 (2015).

79. Rhie, A. et al. The complete sequence of a human Y chromosome. Nature 621, 344–354 (2023).

80. Tuke, M. A. et al. Mosaic Turner syndrome shows reduced penetrance in an adult population study. Genetics in Medicine 21, 877–886 (2019).

81. Hujoel, M. L. et al. Insights into DNA repeat expansions among 900,000 biobank partici-pants. Nature 650, 920–929 (2026).

82. Loh, P. R. et al. Efficient Bayesian mixed-model analysis increases association power in large cohorts. Nature Genetics 47, 284–290 (2015).

83. Taliun, D. et al. Sequencing of 53,831 diverse genomes from the NHLBI TOPMed Program. Nature 590, 290–299 (2021).

84. Das, S. et al. Next-generation genotype imputation service and methods. Nature Genetics 48, 1284–1287 (2016).

85. Loh, P.-R., Kichaev, G., Gazal, S., Schoech, A. P. & Price, A. L. Mixed-model association for biobank-scale datasets. Nature Genetics 50, 906–908 (2018).

86. Barton, A. R., Sherman, M. A., Mukamel, R. E. & Loh, P.-R. Whole-exome imputation within UK Biobank powers rare coding variant association and fine-mapping analyses. Nature Genetics 53, 1260–1269 (2021).

87. Frankish, A. et al. GENCODE: Reference annotation for the human and mouse genomes in 2023. Nucleic Acids Research 51, D942–D949 (2023).

88. GTEx Consortium. The GTEx Consortium atlas of genetic regulatory effects across human tissues. Science 369, 1318–1330 (2020).

89. McLaren, W. et al. The Ensembl Variant Effect Predictor. Genome Biology 17, 122 (2016).

90. Chen, S. et al. A genomic mutational constraint map using variation in 76,156 human genomes. Nature 625, 92–100 (2024).

91. Spence, J. P. et al. Specificity, length and luck drive gene rankings in association studies. Nature 649, 918–925 (2026).

92. Loh, P.-R. et al. Contrasting genetic architectures of schizophrenia and other complex diseases using fast variance-components analysis. Nature Genetics 47, 1385–1392 (2015).

93. Bulik-Sullivan, B. et al. An atlas of genetic correlations across human diseases and traits. Nature Genetics 47, 1236–1241 (2015).

94. Hofmeister, R. J., Ribeiro, D. M., Rubinacci, S. & Delaneau, O. Accurate rare variant phasing of whole-genome and whole-exome sequencing data in the UK Biobank. Nature Genetics 55, 1243–1249 (2023).

95. Loh, P.-R. et al. Reference-based phasing using the Haplotype Reference Consortium panel. Nature Genetics 48, 1443–1448 (2016).

96. Bowden, J., Davey Smith, G. & Burgess, S. Mendelian randomization with invalid instru-ments: effect estimation and bias detection through Egger regression. International Journal of Epidemiology 44, 512–525 (2015).

97. Verbanck, M., Chen, C.-Y., Neale, B. & Do, R. Detection of widespread horizontal pleiotropy in causal relationships inferred from Mendelian randomization between complex traits and diseases. Nature Genetics 50, 693–698 (2018).

98. Rivier, C. A. et al. Genal: a Python toolkit for genetic risk scoring and Mendelian randomization. Bioinformatics Advances 5, vbae207 (2025).

99. 1000 Genomes Project Consortium et al. A global reference for human genetic variation. Nature 526, 68 (2015).

100. Chang, C. C. et al. Second-generation PLINK: rising to the challenge of larger and richer datasets. Gigascience 4, s13742–015 (2015).

