## Supplementary Information for "Genetic mechanisms of mitotic error in 840,853 blood genomes"

**References** **31**

**Supplementary Figures** **40**

**Supplementary Tables** **89**

### Supplementary Notes

#### 1 Benchmarking and validation of WGS depth-based aneuploidy quantification

The mosaic aneuploidy phenotypes that we obtained from WGS read-depth were strongly age-enriched and nearly absent in younger individuals, suggesting tight estimates of aneuploid cell fractions and low false positive rates of aneuploidy calls (Fig. 1 and Supplementary Figs. 1–2 and 13–14). To further evaluate the performance and robustness of aneuploidy quantification from WGS read-depth, we performed several additional analyses detailed below.

##### 1.1 Mosaic X and Y chromosome loss

**Benchmarks of LOX and LOY cell fraction measurement noise (Supplementary Table 1).** We compared the standard errors of four approaches for estimating LOX and LOY cell fractions:

1. **WGS read-depth, denoised by adjusting for 50 PCs** (the approach we used here). We estimated the remaining noise in chromosome-level WGS read-depth estimates by computing the median absolute deviation (MAD, which is robust to outliers) of chrX read-depth among female AoU participants $<$22 years old (n=7,053) and chrY read-depth among male AoU participants $<$22 years old (n=3,299). Among these youngest participants, variation in read-depth primarily reflects measurement error rather than LOX or LOY (Supplementary Fig. 1). We multiplied the MAD by 1.4826 (the ratio of s.d./MAD for a Gaussian distribution) and by an additional factor of 2 for chrX (to convert relative WGS depth deviations to the scale of LOX cell fraction in diploid cell populations). This produced error estimates s.e.(LOX cell fraction) = 0.09%, s.e.(LOY cell fraction) = 0.17%.

2. **WGS read-depth, unadjusted for PCs.** We estimated noise in WGS read-depth unadjusted for PCs (but still GC-corrected and CNV-masked) in the same way, obtaining error estimates s.e.(LOX cell fraction) = 0.37%, s.e.(LOY cell fraction) = 0.25%.

3. **WGS allelic imbalance in PAR1** (for LOY analysis[29]). We estimated the noise of allelic imbalance computed within the PAR1 pseudoautosomal region for 6,852 male UKB participants selected to have at most modest LOY (based on having age 40–41 years, no SNP-array-based mCA calls, and minimal DNA contamination[17]). We computed allelic depths using our WGS BAF analysis pipeline[17] and summed allelic depths for each haplotype across PAR1. We computed the MAD of the allelic imbalance within PAR1, multiplied by 1.4826 to estimate s.e.(PAR1 imbalance), and multiplied by 4 to convert to the scale of LOY cell fraction. This produced an error estimate s.e.(LOY cell fraction) = 1.1%. We did not attempt to evaluate

the noise of BAF-based LOX cell fraction estimates because allelic imbalance analysis underestimates LOX (owing to the presence of competing clones having lost each of the two homologous X chromosomes).

4. **SNP-array allelic imbalance in PAR1** (for LOY analysis[11]). We previously estimated s.e.(BAF deviation) for each LOY call ascertained from allelic imbalance in SNP-array intensities within PAR1[15], so we computed the median s.e.(BAF deviation) across all calls and multiplied by 4 to convert to the scale of LOY cell fraction. This produced an error estimate s.e.(LOY cell fraction) = 1.4%. (This error estimate is specific to the UKB SNP array; SNP arrays vary in probe density and in signal-to-noise of allelic intensities.)

These benchmarks, together with the results below validating the calibration of denoised WGS read-depth, show that careful analysis of read-depth from high-coverage (∼30x) short-read WGS produces much more precise estimates of aneuploid cell fractions than analysis of allelic imbalance. This may seem surprising in light of the widespread adoption of allelic imbalance-based analysis over the past decade[11–18,20,29,77]. However, the reason that allelic imbalance-based analysis was advantageous was that measurements of DNA abundance (from SNP-array probe intensities or WGS read-depth) were difficult to calibrate owing to technical noise. Comparing the relative abundances of pairs of alleles (which were likely to be equally affected by technical noise) circumvented this difficulty, and the reduction in technical noise was sufficiently large to be worth the reduction in signal inherent in limiting analysis to heterozygous sites informative of allelic imbalance (and in the case of LOY, further limiting analysis to PAR1).

Here, we observed that in two large WGS data sets generated from uniformly processed blood-derived DNA (UKB and AoU), the main components of technical noise in WGS read-depth could be learned and eliminated using principal component analysis[17]. Adjusting for read-depth PCs in combination with minimizing biological confounding from germline copy-number variation[17] produced read-depth measurements clean enough that the allelic imbalance-based approach was no longer advantageous. In fact, the relative errors we observed in chromosome-level WGS read-depth are near the theoretical limit from shot noise: e.g., we observed s.e.(chrX depth) = 0.05%, modestly higher than shot noise of 0.03% from sampling ∼13.6 million DNA fragments derived from 136 Mb of unmasked X-chromosome sequence, and we observed s.e.(chrY depth) = 0.17%, modestly higher than shot noise of 0.13% from sampling ∼ 600,000 DNA fragments derived from 12 Mb of unmasked haploid Y-chromosome sequence (excluding the pseudoautosomal regions, ampliconic repeats, and heterochromatin regions[79]).

It is worth noting that the read-depth-based approach does have caveats: in particular, the PCs we computed in UKB and AoU differed somewhat, such that adjusting for read-depth PCs computed in-sample achieved better denoising. Computing read-depth PCs in-sample was possible here but requires having a large reference set of read-depth profiles, which may not be available for other data sets. In such instances, allelic imbalance-based analysis may still be more effective.

**Validation of robustness of WGS read-depth-based LOX and LOY analyses.** We ran several analyses to verify the robustness of our estimates of LOX and LOY cell fractions from chromosome-level read-depth. These included both checks for internal consistency and comparisons to LOX and LOY quantifications using orthogonal methods:

- **Consistency between UKB and AoU (Supplementary Fig. 3)**. The ranges of LOX cell fractions estimated in age-matched participants (restricted to EUR-ancestry never-smokers) were nearly identical in UKB and AoU, and the ranges of LOY cell fractions were also concordant.
- **Homogeneity of read-depth deviations within chromosomes (Supplementary Fig. 5)**. Sex chromosome coverage profiles showed minimal intrachromosomal heterogeneity, indicating that observed deviations in chromosome-level read-depth were not confounded by technical noise and were consistent with whole-chromosome losses.
- **Consistency with SNP-array-based quantification of LOX and LOY (Supplementary Fig. 6)**. Individual-level estimates of LOX and LOY cell fractions in UKB obtained from WGS read-depth were broadly consistent with estimates previously generated using allelic imbalance-based analyses of SNP-array probe intensities[11,14,15]. Differences between the two approaches appeared to be driven by high-cell-fraction LOX or LOY causing drop-out of heterozygous genotypes used in SNP-array allelic imbalance analyses, resulting in deflation of cell fraction estimates or failure to detect LOX or LOY.
- **Consistency with chromosome-level coverage computed by DRAGEN (Supplementary Fig. 7)**. Individual-level estimates of LOX and LOY cell fractions in UKB were also broadly consistent with estimates computed from chromosomal coverages reported in the "Whole genome diagnostics files (DRAGEN)" provided by UKB, with the DRAGEN coverage estimates exhibiting some technical biases between and within sequencing centers (evidenced by vertically-offset bands).

**Benchmarks against WGS coverage analysis using DRAGEN.** To compare our WGS read-depth normalization pipeline to a standard approach, we performed further benchmarks against LOX and LOY cell fractions estimated using chromosomal coverages reported by DRAGEN:

- **Comparison of distributions of estimated LOX and LOY cell fractions (Supplementary Fig. 8)**. The DRAGEN-based estimates exhibited much broader distributions (indicating higher noise) that were multimodal (indicating batch effects) and slightly miscalibrated.
- **Comparison of noise in LOX and LOY cell fraction estimates and GWAS association strengths (Supplementary Fig. 9)**. The DRAGEN-based estimates exhibited several-fold

higher MAD, and LOX and LOY GWAS phenotypes derived from DRAGEN-estimated coverage had several-fold weaker associations with top GWAS hits.

#### 1.2 Autosomal mosaic aneuploidies

We performed similar benchmarking and validation for WGS read-depth-based analyses of autosomal mosaic aneuploidies:

- **Benchmarks of accuracy of denoised autosomal WGS read-depth (Supplementary Table 2)**. For each of the autosomal aneuploidies we analyzed, we computed the MAD of chromosome-level denoised read-depth measurements among AoU participants $<22$ years old (for whom variation in read-depth primarily reflects measurement error) and multiplied the MAD by 1.4826 to estimate the standard error. These error estimates were again near the theoretical limit from shot noise (0.02%–0.045% relative error from sampling $\sim$5–25 million DNA fragments depending on chromosome length). Note that the error estimates in Supplementary Table 2 are on the normalized read-depth scale (such that a value of 1 corresponds to absence of aneuploidy); to obtain standard errors on the cell fraction scale, they need to be multiplied by 2 for aneuploidies of diploid chromosomes.

- **Comparison of read-depth-based and allelic imbalance-based autosomal aneuploidy calls (Supplementary Fig. 18)**. WGS read-depth-based analysis detected several-fold more autosomal mosaic aneuploidies than allelic imbalance-based analysis[17] whether using a call threshold of the probabilistic phenotype being $>0.5$ or $>0.9$ (Supplementary Fig. 18a,b). For individuals with BAF-based mCA calls, the BAF-based calls were predominantly whole-chromosome mosaic aneuploidies (Supplementary Fig. 18c) as expected for the 10 autosomal aneuploidies we analyzed[17,20]. As expected, for 5 of the 10 aneuploidies, read-depth deviations almost exclusively reflected whole-chromosome mosaic aneuploidies; the others included a noticeable minority of sub-chromosomal mCAs.

- **Evaluation of intrachromosomal read-depth variability for autosomal aneuploidy calls (Supplementary Fig. 19)**. For the 5 autosomes for which read-depth deviations arose from a mix of full-chromosome mosaic aneuploidies and sub-chromosomal mCAs, read-depth measurements within subchromosomal regions (e.g., p-arm-only) confirmed that most non-full-chromosome mCAs were arm-level gains, as expected[17,20]. We did not attempt to identify and filter out these events, as this was usually not possible for low-cell-fraction events, and arm-level gains were still potentially informative in GWAS searching for genetic effects on mitotic missegregation (e.g., if they are generated by merotelic attachments that result in splitting of bipartite kinetochores[35]).

### 2 GWAS of mosaic aneuploidy phenotypes

#### 2.1 Details of genome-wide association analyses

**UKB GWAS of LOX and LOY.** We ran GWAS of log-transformed, clipped LOX and LOY cell fractions in EUR-ancestry UKB participants using the non-infinitesimal linear mixed model (LMM) implemented in BOLT-LMM[82] v2.5, including age, age$^2$, smoking status, and 20 genetic PCs as covariates. We fit the LMM using UKB SNP-array genotypes.

In addition to testing TOPMed-imputed variants[83,84] (MAF>0.1%, INFO>0.3), we also tested variants in the pseudoautosomal regions (PAR1 and PAR2) called by the DRAGEN WGS pipeline[27], as the pseudoautosomal regions had been excluded from imputation. We applied the following QC filters to variants from the DRAGEN WGS genotyping: male-female AF difference <0.002, Hardy-Weinberg $P>1\times10^{-6}$ (computed using plink[100] v2.0 within the EUR-ancestry sample set from the LOY GWAS), and genotyping rate >0.9. This QC was important; without the QC filters, technical artifacts in the DRAGEN genotype calls (perhaps driven by high-cell-fraction LOY biasing genotype calls for some poorly genotyped variants) generated many false-positive associations of PAR1/PAR2 variants with LOY.

**AoU GWAS of LOX and LOY.** We ran GWAS of log-transformed, clipped LOX and LOY cell fractions in each of the EUR, AMR, and AFR ancestry groups in AoU v8 (restricting to the individuals with blood-derived WGS data that we profiled for aneuploidies). We used the infinitesimal LMM implemented in BOLT-LMM v2.5, fitting it using AoU SNP-array variants with ancestry-specific MAF>0.01, and we included age, age$^2$, smoking status, and 16 genetic PCs as covariates.

We tested variants from the AoU WGS "ACAF threshold" call set that passed ancestry-specific allele frequency thresholds and were present in externally curated variant sets, providing a QC filter. For the EUR-ancestry GWAS, we filtered to variants with MAF>0.1% in the TOPMed-imputed UKB data set. For the AMR and AFR ancestries, we filtered to variants with MAF>0.5% in the gnomAD[90] v4.1 AMR and AFR ancestry groups, respectively. In each GWAS, we further restricted to variants with allele count ≥40, MAF>0.1%, and genotyping rate >0.9 within the set of samples analyzed. Finally, we dropped the X chromosome from the AoU LOY GWAS because of a known issue with the encoding of male chrX genotypes in the AoU v8 ACAF bgen file; this issue created false-positive associations with LOY throughout the X chromosome.

**UKB GWAS of autosomal aneuploidies.** We ran GWAS of probabilistic calls of 10 autosomal aneuploidies in EUR-ancestry UKB participants using linear regression implemented in BOLT-LMM v2.5, including age, age$^2$, smoking status, and 20 genetic PCs as covariates. We restricted each GWAS to unrelated individuals by dropping one individual per second-degree or closer relationship pair (kinship >0.0884 computed by UKB[75]), prioritizing individuals to retain based on aneuploidy phenotype value, breaking ties in favor of older age.

We again tested TOPMed-imputed variants (MAF>0.1%, INFO>0.3) that we subsequently filtered more stringently on MAF to avoid test statistic inflation[85]. This was necessary because the probabilistic aneuploidy call phenotypes that we analyzed were not binary, such that saddlepoint approximation and Firth regression techniques[101,102] could not be used. We imposed phenotype-specific thresholds of MAF>50/(# cases), where we treated the sum of the probabilistic phenotype values across the GWAS sample set as the number of cases. This resulted in MAF thresholds ranging from 0.4% to 3.5% depending on phenotype frequency (Supplementary Table 3).

We also tested pericentromeric variants from the DRAGEN WGS genotype call set after observing significant associations of TOPMed-imputed pericentromeric variants on chromosomes 15, 21, and 22 with mosaic aneuploidies involving these chromosomes. We further observed that imputation accuracy was low in these regions, such that associations might be underestimated or missed by analyzing imputed data. For each autosomal aneuploidy GWAS in UKB, we therefore also included DRAGEN WGS variant calls by within 1 Mb of the centromere of the affected chromosome, filtering to variants with FILTER=PASS.

**AoU GWAS of autosomal aneuploidies.** We ran GWAS of probabilistic calls of 10 autosomal aneuploidies in each of the EUR, AMR, and AFR ancestry groups in AoU using linear regression implemented in BOLT-LMM v2.5, including age, $age^2$, smoking status, and 16 genetic PCs as covariates. We restricted to unrelated individuals within each ancestry group based on the AoU v8 sample relatedness file (containing relationship pairs with kinship coefficient >0.1), prioritizing individuals to retain based on total aneuploidy phenotype value across the 10 aneuploidies, breaking ties in favor of older age.

As in UKB, we applied phenotype-specific MAF thresholds to the AoU autosomal aneuploidy GWAS. We set the minimum MAF threshold for each GWAS as min(0.02, 25/(# cases)), where we again treated the sum of the probabilistic phenotype values across the GWAS sample set as the number of cases. This resulted in MAF thresholds of 0.35% to 2% for the AoU EUR-ancestry autosomal aneuploidy GWAS and 0.66% to 2% for the AoU AMR and AFR ancestry groups (Supplementary Table 3). We restricted to variants from the AoU WGS “ACAF threshold” call set with MAF>0.1% in the TOPMed-imputed UKB data set (with MAF computed in UKB participants of all ancestries). Finally, we restricted to variants with allele count ≥40 and genotyping rate >0.9 within the set of samples analyzed in each GWAS.

#### 2.2 Comparison between aneuploidy GWAS results in UKB and AoU

**UKB versus AoU EUR-ancestry GWAS.** The EUR-ancestry GWAS in UKB and AoU were broadly concordant for all 12 aneuploidy phenotypes. Sign tests for replication showed that lead variants from the UKB EUR GWAS replicated robustly in AoU EUR (97% consistent effect directions; Supplementary Fig. 31 and Supplementary Tables 4–6). In the reverse direction, 100% of lead variants from the AoU EUR GWAS (201 GWAS hits across the 12 aneuploidies) exhibited consistent effect directions in UKB.

Some differences in relative association strengths in AoU versus EUR were likely driven by allele frequency differences in Northern versus Southern European ancestries. For example, the top association in the AoU EUR GWAS of mosaic chromosome 3 gain was rs144339562 (chr6:32632142:A:G), which is several times more common in Southern European compared to Northern European populations, explaining the lower relative contribution of the HLA locus in the UKB EUR GWAS (Supplementary Fig. 21).

**AFR- versus EUR-ancestry GWAS.** GWAS of non-European ancestries in AoU identified several associations driven by ancestry-specific or ancestry-enriched variants. For LOX, the top two associations in AoU AFR were rs2814778 at *ACKR1* ($P$=$3.2\times10^{-134}$) and rs115126447 at *SP140L* ($P$=$7.0\times10^{-101}$; Supplementary Table 7). The rs2814778-CC genotype generates the malaria-protective Duffy-null phenotype[103] and also strongly associates with reduced neutrophil count[104]); the latter effect probably mediates the association of rs2814778 with LOX levels measured in blood-derived DNA, which increase with lymphocyte percentage (Supplementary Fig. 11). At *SP140L*, rs115126447 represents a distinct genetic effect from those observed in European ancestries: rs115126447 is an African-specific variant (AFR AF=2%) in the promoter region of *SP140L*. Another top LOX association in AoU AFR involved rs531991719, an African-specific (AFR AF=0.5%) 25-bp deletion causing loss of the start codon of *OIP5*. *OIP5* encodes MIS18B, a subunit of the MIS18 complex, which facilitates CENP-A loading at centromeres[45]. Similarly, LOY GWAS in AoU AFR identified several AFR-specific variants; these included missense variants in *MPL* (rs2840287), *XPC* (rs3731152), and *CHEK2* (rs17886163) (AFR AF=1–5%), each of which was the lead variant at its locus (Supplementary Table 8).

#### 2.3 Allelic shift tests for autosomal gains

Several common-variant associations with autosomal aneuploidies (specifically, gains) involved variants located within the chromosome affected by the aneuploidy. For these variants, we ran "allelic shift" analyses that tested whether the homolog that was gained tended to carry the REF or the ALT allele more frequently in heterozygous carriers. We did so using the same approach we used in our recent analysis of CN-LOH allelic shifts[17]. Briefly, we identified UKB participants with mosaic gains called on the chromosome under consideration (using our allelic imbalance-based pipeline[17]); for these calls, we determined the phase of the mosaic gain relative to the phase of the associated variant, and we ran a binomial test to determine whether the REF or the ALT allele was more often gained (Supplementary Table 10).

We used a separate approach to run allele-specific association tests on pericentromeric variants obtained from DRAGEN genotype calls, as these variants had not been included in our earlier analysis pipeline and were also more difficult to phase (Supplementary Note 4).

#### 2.4 Gene-based burden tests of aneuploidy phenotypes

We performed gene-based burden tests to identify genes for which a burden of rare protein-altering variants associated with mosaic aneuploidy. We used burden masks that collapsed together variants predicted to cause loss of function (LoF) and optionally included missense variants satisfying PrimateAI-3D score[105] thresholds (>0.6, 0.7, 0.8). These burden masks also applied varying options for the choice of allele frequency threshold, inclusion or exclusion of CNVs[106], and choice of transcript(s), altogether defining 64 burden masks per gene[17].

We ran burden tests on log-transformed, clipped LOX and LOY cell fractions in UKB using BOLT-LMM[82] under the same setup we used for the common-variant GWAS. This was appropriate even for testing very rare burden masks because the LOX and LOY GWAS phenotypes did not have heavy tails or outliers (Supplementary Fig. 15), so we only required a minimum allele count of 5 within each GWAS. To reduce multiple testing burden, we excluded singleton-only burden masks (leaving the allele frequency thresholds AF<0.01, 0.001, 0.0001), and to reduce the potential for burden masks to tag common variants, we limited the AF<0.01 masks to variants with LoF annotations; this left 36 masks per gene. Finally, we excluded burden masks incorporating CNVs for genes within regions that commonly contain somatic CNVs:

- *IGH*: chr14:105.5-106.9 Mb (GRCh38)
- *IGK*: chr2:88.8-90.3 Mb
- *IGL*: chr22:22.0-23.0 Mb
- *TRA*, *TRD*: chr14:21.6-22.6 Mb
- *TRB*: chr7:142.2-142.9 Mb
- *TRG*: chr7:38.2-38.4 Mb
- 16p11.2: chr16:29.5-30.2 Mb
- 13q14: chr13:45-52 Mb

This left ∼600,000 burden masks across ∼18,500 genes. However, these comprised only ∼280,000 unique variant sets (as different variant inclusion criteria often resulted in selection of exactly the same variants), and even among the unique variant sets, many were very similar. We therefore defined a significance threshold $P<1\times10^{-5}$ that controlled FDR at 0.011 (Supplementary Fig. 20 and Supplementary Tables 11–12).

The two strongest burden associations with LOY involved protein-altering mutations in *TET2* ($P$=$9.9\times10^{-78}$) and *DNMT3A* ($P$=$7.6\times10^{-76}$), the two most common drivers of clonal hematopoiesis with indeterminate potential (CHIP)[107,108]. Previous studies have observed associations of CHIP mutations with decreased LOY[12,29,109], suggesting that some burden associations might be driven by somatic mutations. To identify such associations, we evaluated the extent to which restricting the burden masks to variants with confident statistical phase[94] (reflecting likely germline origin) diminished association strengths. This identified nine burden associations with LOY (*TET2*, *DNMT3A*, *JAK2*, *CALR*, *SRSF2*, *ASXL1*, *PPM1D*, *GNB1*, and *SF3B1*) and one with reduced LOX (*DNMT3A*) that were likely to be driven by somatic mutations. All of these genes are common drivers of myeloid CHIP[110], and all associated with reduced LOY, perhaps reflecting clonal competition between myeloid CHIP clones and cell populations with LOY[111,112].

We also ran burden tests for mosaic autosomal aneuploidies in UKB. In order to perform rare-variant tests on these rarer phenotypes, we binarized the probabilistic call phenotypes (rounding values >0.5 to 1) and performed association testing using REGENIE[102] `--firth --approx --pThresh 0.01`. We included age, age$^2$, smoking status, and 20 genetic PCs as covariates and restricted to unrelated EUR-ancestry UKB participants, prioritizing individuals to retain based on total aneuploidy phenotype value across the 10 autosomal aneuploidies, breaking ties in favor of older age. We applied a significance threshold of $P<1\times10^{-7}$ to control FDR at $\sim$0.01.

The effect size estimates for *SETDB2* LoF/missense burden shown in Fig. 3f correspond to the SETDB2.LoF.missense8.0.001 mask, which collapsed LoF SNP/indel variants and missense variants with PrimateAI-3D score[105] >0.8 in the MANE Select[113] transcript for *SETDB2*, restricting to variants with allele frequency <0.1%. This mask did not include LoF CNVs to ensure that the association results were not affected by somatic del(13q14) events.

### 3 Putative functional variants in kinetochore and centromere genes

In Fig. 3b, we display effect sizes from GWAS across mosaic aneuploidy phenotypes for 12 variants in kinetochore and centromere genes at GWAS loci for one or more aneuploidies. For each of these variants, we had high confidence in the target gene and could identify a putative functional effect of the variant or haplotype:

- *MAD1L1* R558H (chr7:1936821:C:T): common missense variant in near-perfect LD with top SNP ($R^2$=1.00).
- *MAD2L1* sQTL (chr4:120063721:A:G): lead haplotype; $R^2$=0.95 with top sQTL for *MAD2L1* in GTEx; causal variant is unclear.
- *SPDL1* R20Q (chr5:169588475:G:A): lead variant at locus.
- *ZWILCH* S344G (chr15:66528912:A:G): common missense variant in near-perfect LD with top SNP ($R^2$=0.99).
- *KNTC1* L317F (chr12:122547931:C:T): lead variant at locus.

- *NDC80* E348D (chr18:2595444:G:T): common missense variant in strong LD with top SNP ($R^2$=0.89; $P$=2.0×10$^{-8}$ compared to $P$=2.8×10$^{-9}$ for lead variant at locus for LOY).

- *PMF1* Q75R (chr1:156232382:A:G): lead variant at locus.

- *CENPW* promoter (chr6:126340008:T:C): lead variant at locus; PromoterAI[52] score −0.32.

- *CENPQ* D266G (chr6:49492265:A:G): common missense variant in near-perfect LD with top SNP ($R^2$=1.00); alternatively, could potentially function as a cryptic splice donor gain (Pangolin[114] score 0.50).

- *CENPU* eQTL (chr4:184731717:A:T): lead haplotype; $R^2$=1.00 with top eQTL for *CENPU* in GTEx, associated with increased expression.

- *CENPR* eQTL (chr1:63492775:A:C): lead haplotype; $R^2$=0.99 with top eQTL for *ITGB3BP* (the official name for *CENPR*) in GTEx, associated with increased expression.

- *CENPT* R507C (chr16:67828517:G:A): lead variant at locus.

We also searched for a potential functional variant at the *SETDB2* locus, which appears to affect missegregation of chromosomes 21 and 22, probably by modulating H3K9me3 methylation of centromeric chromatin (Fig. 3g). In Fig. 3f, we display effect sizes for the lead variant chr13:49518324:CTA:C ($P$=2.4×10$^{-72}$) from the GWAS of chromosome 21 gain in UKB EUR. This index variant is probably not causal; in UKB EUR, it sits on a haplotype containing 15 variants with $P$=8.3×10$^{-68}$–2.4×10$^{-72}$. Examining the same GWAS locus for chromosome 21 loss in AoU AFR (in which LD was much shorter) provided some clarity: the top 3 variants (chr13:49501793:T:C, $P$=8.3×10$^{-30}$; chr13:49506711:A:G, $P$=1.9×10$^{-29}$; chr13:49500616:A:G, $P$=2.4×10$^{-27}$) clustered ∼5–10kb downstream of *SETDB2* (in the first intron of *PHF11*). Among these three variants, chr13:49500616:A:G seems most likely to be functional based on functional annotations[115].

### 4 Associations of centromere haplotypes with mosaic aneuploidy

We performed several follow-up analyses to further investigate and validate the centromeric GWAS loci on chromosomes 15, 21, and 22 associated with mosaic gains of these chromosomes.

#### 4.1 Allele-specific association tests for gain of the homolog in *cis* or *trans*

Under the model in which centromere alleles have varying propensities to missegregate during mitosis, centromeric risk alleles should increase risk of aneuploidy involving the homolog in *cis* and have no effect on risk of aneuploidy involving the homolog in *trans*. To evaluate whether each of the three lead variants (chr15:19900978:T:C, chr21:13216980:A:G, and chr22:15432903:C:T) associated with gain of the homolog in *cis* or gain of the homolog in *trans*, we restricted analysis to autosomal mosaic gains in UKB called from allelic imbalance[17], as the phase of the gained homolog relative to heterozygous SNPs could be confidently assigned for such gains. Specifically, we identified gain calls that spanned all or nearly all of the chromosome (>95% the length of the longest call), and for each such call, we extracted the genotype of the lead variant as well as the phase of the gain relative to the individual's SNP-haplotype scaffold at the centromere (i.e., whether haplotype 1 or haplotype 2 had been gained).

For heterozygous carriers of the lead variant, we next determined the phase of the variant relative to the SNP-haplotype scaffold (which did not include these pericentromeric variant calls from DRAGEN). To do so, we used a phasing approach similar to our previous work (Supplementary Note 4.2.2 of ref.[81]). For each UKB participant, we identified the 10 longest identical-by-descent (IBD) matches ("haplotype neighbors"[106]) for each of the individual's two SNP-haplotypes at the centromere. We then ran the following iterative phasing algorithm:

- Initialize each haplotype to have dosage equal to half the diploid genotype.
- For each of 10 iterations:
  - For each individual in turn, partition the individual's diploid genotype into haploid dosages proportional to values imputed from the 10 "haplotype neighbors" of each haplotype. We computed each imputed haploid dosage as the weighted mean of the current estimates of the dosages of the 10 haplotype neighbors, using weights proportional to exp(–$c$/(IBD length)) to prioritize more recent IBD-sharing.

We tuned the $c$ parameter to maximize the correlation between observed diploid genotypes and sums of imputed haploid genotypes (evaluating $c = \{0, 0.5, 1, \ldots, 10\}$ cM in each of the first three iterations and using the optimal value from the third iteration for subsequent iterations). We dropped heterozygous individuals for which the phase of the lead variant was uncertain at the end of this procedure (based on the imputed dosages of the two haplotypes differing by $<0.2$ for the rarer chr15 and chr22 variants and by $<0.5$ for the common chr21 variant).

We performed haplotype-specific tests for association of each variant with gain of the homolog in *cis* or the homolog in *trans* using Fisher's exact test on 2x2 contingency tables in which the rows corresponded to haploid alleles of the lead variant and the columns corresponded to whether or not the homolog in *cis* (respectively, *trans*) was somatically gained. For the row corresponding to the REF allele, we used observations from hom-REF individuals: for $n$ mosaic gains observed among $N$ participants with $2N$ haplotypes, we counted $n$ instances in which the haplotype in *cis* was gained and $2N - n$ instances in which the haplotype in *cis* was not gained (with the same counts for the haplotype in *trans* being gained vs. not gained). For the row corresponding to the ALT allele, we used observations from confidently phased heterozygotes. For the *cis* association test, we counted the numbers of carriers for which the ALT allele was gained vs. not gained, and for the *trans* association test, we counted the numbers of carriers for which the REF allele (in *trans* to the ALT allele) was gained vs. not gained. We restricted these association analyses to EUR-ancestry UKB participants.

These tests confirmed that the associations of centromeric haplotypes with mosaic gains were specific to the homolog in *cis* (insets of Fig. 3c–e).

##### 4.2 Stepwise conditional analyses of pericentromeric variants

The strong associations of the lead pericentromeric variants ($P$=$8.4\times10^{-32}$–$1.3\times10^{-119}$; Fig. 3c–e) suggested the possibility that other centromere haplotypes (cenhaps) might independently confer risk of mosaic aneuploidy, particularly given that the lead variants for gains of 15 and 22 had low frequencies (AF=0.66% and 0.85% in UKB EUR). To search for additional cenhap effects, we performed stepwise conditional analyses using the same GWAS setup that we used to test DRAGEN pericentromeric variants in our primary analyses. Specifically, we iteratively identified a lead variant, added it to the set of covariates, and repeated the analysis, iterating until the lead variant no longer reached $P<5\times10^{-8}$ in either the conditional or original analysis. This procedure identified the following independent associations ($P$-values corresponding to the original analysis):

- Chromosome 15 gain:
  - chr15:19900978:T:C (AF=0.0066, $P$=$1.3\times10^{-119}$)
  - chr15:19781862:A:C (AF=0.0027, $P$=$3.1\times10^{-53}$)
    This position is in an $\alpha$-satellite region, such that the DRAGEN variant calls may reflect misalignments; however, the variant calls showed good consistency between IBD2 siblings in UKB, such that they probably tag a real cenhap.
- Chromosome 21 gain:
  - chr21:13216980:A:G (AF=0.42, $P$=$4.8\times10^{-46}$)
  - chr21:13437212:A:G (AF=0.05, $P$=$1.2\times10^{-18}$)
  - chr21:10501901:G:A (AF=0.04, $P$=$6.5\times10^{-25}$)
- Chromosome 22 gain:
  - chr22:15432903:C:T (AF=0.0085, $P$=$8.4\times10^{-32}$)
  - chr22:15493984:T:C (AF=0.49, $P$=$2.1\times10^{-13}$)

##### 4.3 Evidence of robustness of cenhap associations

Centromeric regions are prone to genotyping artifacts and contain unusually long haplotypes (sometimes spanning several megabases[116]), so we took precautions and ran several follow-up analyses to confirm that the associations that we identified with mosaic autosomal aneuploidies were robust:

- **Rounded autosomal aneuploidy phenotypes to zero for most individuals.** We rounded probabilistic call phenotypes below 0.05 down to 0, such that for each autosomal aneuploidy phenotype, the large majority of individuals had phenotypes of exactly zero (Supplementary Fig. 17; in each panel, the percentage of nonzero phenotypes is indicated in the top-right). This reduced the potential for the read-depth-derived aneuploidy phenotype to capture slight deviations in read-depth driven by structural variation in or near the centromere that escaped our CNV-masking procedure and could be tagged by pericentromeric SNPs.

- **Replicated associations in AoU.** We confirmed that the chr15, chr21, and chr22 pericentromeric associations with mosaic gains of these chromosomes in UKB replicated in AoU (Supplementary Table 6). The lead variant for the chromosome 21 locus in UKB was not genotyped in AoU, but we replicated the associations of other variants on its haplotype.

- **Checked consistency of associations with gain and loss of chromosome 21.** We verified that in both UKB and AoU, the ALT allele of the lead pericentromeric variant, which associated with decreased gain of chromosome 21, also associated with decreased loss of chromosome 21 (Supplementary Fig. 34). This is consistent with the ALT haplotype being in linkage disequilibrium with a centromere allele with improved mitotic fidelity (such that missegregation, which can produce either gain or loss of the chromosome, is broadly decreased). In contrast, a technical artifact involving a spurious decrease in read-depth in ALT allele carriers would have an opposite-direction, risk-increasing effect on the read-depth-derived loss of chromosome 21 phenotype.

- **Verified that associations were *cis*-specific and robust to restricting analysis to higher-cell-fraction gains called from allelic imbalance analysis.** These analyses are described in Supplementary Note 4.1.

- **Verified genotyping quality of lead variants.** We checked that all three lead variants were genotyped in gnomAD v4.1.1 with EUR AF concordant with UKB, and we further verified that within UKB, genotypes of IBD2 siblings showed good consistency.

### 5 Identification of hybrid chromosomes from short-read WGS

The strong associations of centromeric haplotypes of chromosomes 15, 21, and 22 with somatic gains of the chromosomes containing these haplotypes led us to wonder whether any of these associations could be driven by structural variation within centromeres. Centromeric sequence variation is generally inaccessible to short reads, but recent advances in long-read sequencing and assembly techniques have enabled assembly of complete human centromeres[24–26]. Such analysis has been performed for a few hundred individuals to date, mostly from the 1000 Genomes Project (1KGP)[58,99] as part of the HGSVC and HPRC projects[25,26,117].

We were particularly curious about whether the low-frequency haplotypes at the centromeres of chromosomes 15 and 22 (which associated with several-fold increased risk of gains of the chromosomes containing these cenhaps; OR=10.8 [6.9–16.3] and 3.7 [2.0–6.4], Fig. 3c,e) might tag centromere alleles with unusual properties. To investigate this possibility, we searched for carriers of these haplotypes among the HGSVC3 and HPRC2 cohorts. We did not find any carriers of chr15:19900978:T:C, but chr22:15432903:C:T had one carrier in HGSVC3 (NA20355, ASW) and two in HPRC2 (HG00235, GBR; HG00639, PUR)—exactly matching the three individuals recently observed to carry a hybrid centromere on chromosome 22 created by ectopic recombination with chromosome 13 (ref.[26]).

This led us to undertake further analyses to genotype the hybrid 13p–22q structural variant in biobank WGS data sets, characterize its global allele frequency distribution, and search for other arm-level structural variations formed by ectopic recombination of acrocentric p-arms.

#### 5.1 Hybrid chromosome 13p–22q in UKB

We used two approaches to identify UKB participants carrying the hybrid chromosome 13p–22q based on short-read WGS data: read-depth analysis and analysis of chimeric sequence generated by the ectopic recombination.

**Read-depth analysis.** The 13p–22q hybrid chromosome can be thought of as a chromosome 22 in which the p-arm has been replaced by the p-arm of chromosome 13 (ref.[26]). As such, heterozygous carriers of the hybrid chromosome have three copies of 13p and one copy of 22p. In theory, this should produce a signature of elevated read-depth of short-read WGS alignments to 13p and reduced read-depth in 22p, allowing genotyping of the 13p–22q recombination.

In practice, read-depth analysis in the p-arms of acrocentric chromosomes is very challenging because of the high homology within and between these arms, compounded with the approximate representation of these arms in the GRCh38 reference (used for read-alignment of most existing biobank WGS data[27,28]). The GRCh38 reference only contains p-arm sequence for chromosomes 21 and 22, whereas chr13:1–16000000, chr14:1–16000000, and chr15:1–17000000 are all N's. As such, short read pairs from p-arms of the acrocentrics are often mismapped, and in lieu of having reference sequence for 13p, reads from 13p are often aligned to 21p (owing to higher homology between these p-arms). However, despite these challenges, GRCh38 read-depth based analysis of p-arm structural variation is still possible with sufficient care, as recently demonstrated for genotyping of Robertsonian translocations[118].

We took an empirical approach to identifying GRCh38 regions within 21p (used as a proxy for

13p) and 22p with read-depth sufficiently well-behaved in the UKB WGS data set to be suitable for analysis:

- For 22p, our overall strategy was to find regions in which read-depth (relative to genome-wide coverage) was close to 1 and had low variability across samples (indicating limited effects of copy-number variation and sequence variation affecting read alignment). We used mosdepth[119] v0.3.1 to compute read-depth profiles across 100-bp bins in 22p. Based on a pilot analysis across the full length of 22p, we prioritized the region chr22:12.5–12.6 Mb for profiling across the UKB cohort. Within this region, we selected 54 100-bp bins within the sub-region chr22:12531901–12582900 with mean read-depth 0.9–1.1 (relative to genome-wide coverage) and s.d.$<$0.2.

- For 21p, 100-bins generally seemed to capture reads from both 13p and 21p. Based on a pilot analysis, we prioritized the region chr21:96.6–96.8 Mb for profiling across the UKB cohort. Within this region, we selected 14 100-bp bins within chr21:9662501–9671900 with lower variation (s.d./mean $<$ 1/7).

Plotting read-depth in the 21p target regions (attempting to measure copy-number in 13p+21p) against read-depth in the 22p target regions (attempting to measure copy-number in 22p) showed that UKB participants heterozygous for the aneuploidy-associated lead variant chr22:15432903:C:T almost perfectly matched a cluster of individuals with roughly halved read-depth in 22p (consistent with loss of 22p) and roughly $\sim$1.25-fold increased read-depth in 21p (consistent with gain of 13p, increasing 13p+21p copy number from 4 to 5; Fig. 3i and Supplementary Fig. 35d). Similarly, individuals homozygous for chr22:15432903:C:T formed a cluster with twice these read-depth deviations. This indicated that chr22:15432903:C:T is a near-perfect tag for the hybrid 13p–22q chromosome 22 structural variant in UKB.

**Chimeric sequence analysis.** The full centromeric sequence of the hybrid 13/22 centromere was recently assembled for three carriers, demonstrating a shared chimeric sequence generated by ectopic recombination[26]. This enabled an orthogonal approach to identifying carriers of 13p–22q hybrid chromosomes based on presence of the chimeric sequence in short reads. To implement this approach, we identified at 65-bp sequence spanning the breakpoint (TTCTGTGAAACTTGTTTGT-GATGTGTGTACTCAACTAACAGAGTTGAACCTTTCTTTTGATGCAG) that did not have any close matches in T2T-CHM13. We then searched for reads containing exact matches to this sequence among UKB WGS reads that had been aligned to the chr13 centromere in GRCh38.

This confirmed that chr22:15432903:C:T is a very good tag for the hybrid 13/22 centromere: $\sim$90% of carriers of the tag SNP had chimeric reads, and conversely, $\sim$99.4% of individuals with chimeric reads carried the tag SNP (Supplementary Fig. 35c). The $\sim$10% of tag SNP carriers for which no chimeric reads were observed still tended to have read-depth deviations consistent with loss of 22p and gain of 13p, suggesting that $\sim$10% of hybrid 13/22 centromeres have lost the chimeric breakpoint sequence due to mutation.

##### 5.2 Hybrid chromosomes in 1KGP

To assess the extent to which chr22:15432903:C:T tags the hybrid 13/22 centromere in other populations, and to search for other arm-level swaps of acrocentric p-arms generated by ectopic recombination, we also performed read-depth analyses of the 1000 Genomes 30x WGS data set[58]. Here, alignments to the T2T-CHM13 reference were available, and 500-bp mosdepth coverage profiles had previously been generated and released[120]. We therefore analyzed the available 500-bp depth profiles, identifying reasonably well-behaved bins on 13p, 14p, 15p, 21p, and 22p based on having mean depth 0.9–1.1 (relative to median autosomal coverage) and s.d.$<$0.14. We then estimated p-arm coverage for each 1KGP participant (n=3,202) based on mean relative coverage across selected 500-bp bins within each of the five acrocentric chromosomes (Supplementary Table 15).

These analyses confirmed that chr22:15432903:C:T is a near-perfect tag for the hybrid 13p–22q chromosome: among 35 carriers in 1KGP (across all populations), 34 formed a cluster with aberrant read-depth supporting the ectopic recombination (Supplementary Fig. 35b). Moreover, 10 of the 34 individuals belonged to five parent-child duos, confirming germline inheritance of this structural variant. As expected, the read-depth deviations measured from selected 500-bp bins were smaller in magnitude than the deviations we had observed in our analyses of UKB WGS data using 100-bp bins that could more precisely target informative regions; however, they still provided better separation of genotype clusters, reflecting the benefits of aligning reads to T2T-CHM13 rather than GRCh38[121] (Supplementary Fig. 35b,d). We also tried analyzing alignments of 1KGP WGS data to GRCh38 using the 100-bp bins we had used for UKB, but we observed that these bins did not accurately measure p-arm ploidy in the 1KGP alignments, presumably reflecting differences in short-read alignment pipelines.

We also looked more generally for evidence of other p-arm swaps in the read-depth data for 1KGP participants, observing one other putative event in three ESN participants (HG03116 and a duo, HG03270 and HG03272). All three samples exhibited decreased 13p read-depth and increased 14p read-depth (Supplementary Table 15). This suggests that another ancient ectopic recombination in an African population may have replaced part or all of 13p with a corresponding portion of 14p; however, further characterization of this event will require analyses of long-read sequencing data. We did not observe evidence of any large-scale p-arm swaps involving chromosome 15, suggesting that the low-frequency variant chr15:19900978:T:C tags a different form of variation that underlies its association with increased somatic gain of chromosome 15.

These analyses were only able to detect p-arm swaps generated by ectopic recombination in or near centromeres, such that read-depth across each affected p-arm was increased or decreased to an extent detectable from short-read coverage in 500-bp windows. Based on these data, such swaps appear to be much rarer than ectopic recombinations in subtelomeric regions and within p-arms of acrocentric chromosomes[117,122–125].

##### 5.3 Population allele frequencies and likely origin of hybrid chromosome 13p–22q

The near-perfect LD between chr22:15432903:C:T and the hybrid 13p–22q chromosome allowed us to explore the allele frequency of this structural variation across different populations using chr22:15432903:C:T as a proxy. Examining the allele frequencies of the tag SNP across genetic ancestry groups in AoU, 1KGP Phase 3 (n=2,504), and UKB showed that the tag SNP had highest frequency in European and admixed American ancestries ($\sim$1% in each group), lower frequency in South Asian populations ($\sim$0.3–0.5%), and was absent in East Asian and unadmixed African populations (Supplementary Fig. 35a). Within European-ancestry populations, the allele frequency was higher in AoU EUR than in UKB EUR; this, together with the similarly higher frequency in AoU AMR, suggested that the hybrid chromosome might have even higher frequencies in individuals of Southern European ancestry, comprising only a small fraction of the UKB EUR cohort.

To estimate allele frequencies within Southern and Northern European ancestry groups, we clustered the AoU EUR cohort into TSI-like, IBS-like, and GBR+CEU-like subgroups using previously computed coordinates along 16 genetic principal components[28]. Specifically, we computed Euclidean distances from AoU EUR-ancestry participants (in `ancestry_preds.tsv`) to the centroids of the TSI, IBS, and GBR+CEU reference sets (in `training_pca.tsv`), and we assigned each AoU participant to the group with the closest centroid. Computing the allele frequency of the tag SNP within each group showed that the recombined haplotype was indeed enriched in the Southern European ancestry groups (TSI and IBS; Supplementary Fig. 35a):

- TSI-like AF = 1.7% (n=32,964)
- IBS-like AF = 1.6% (n=12,579)
- GBR+CEU-like AF = 1.0% (n=188,810)

Collectively, these analyses suggest that the hybrid 13p–22q chromosome originated from a single ancient ectopic recombination event in an ancestral Southern European population, and that it has since been transmitted (through germline inheritance) throughout European and European-admixed populations, where it is now observed at up to $\sim$2% allele frequency. The initial observation of this event in centromere assemblies from three distinct 1KGP populations (HG00235, GBR; HG00639, PUR; NA20355, ASW)[26] presumably reflects admixed EUR-ancestry tracts in the PUR and ASW participants at their chromosome 22 centromeres.

The commonness of the hybrid chromosome suggests that it is a benign structural variation that does not reduce fitness. Consistent with this, we did not observe an association of the tag SNP with any clinical phenotype in AoU (Supplementary Note 8). This is reasonable, as the acrocentric p-arms contain similar components (e.g., ribosomal DNA arrays) that are interchangeable and even dispensable in the case of Robertsonian translocations, for which carriers are observed in healthy population cohorts[118] and are generally healthy aside from having reduced fertility. The 13p–22q hybrid chromosome has the additional feature of containing a recombined centromere that we observed associates with ∼4-fold increased somatic aneuploidy (relative to other chromosome 22 centromeres); however, mitotic segregation may be sufficiently robust or the consequences of occasional missegregation of chromosome 22 sufficiently minor that this effect does not noticeably affect health.

### 6 Genotyping and association analyses of *ICCE* and *DXZ4* copy-number variation

#### 6.1 *ICCE* and *DXZ4* copy number estimation

We estimated copy number of the *ICCE* and *DXZ4* repeats in UKB and AoU by counting WGS read pairs aligned to each repeat region in GRCh38, correcting these counts for GC bias using GC profiles estimated during genome-wide read-depth profiling[17]. As in our GC-profiling pipeline, we counted reverse-strand reads with mates mapped to the forward strand, excluding non-primary alignments, reads failing QC, and supplementary alignments (i.e., requiring SAM flag 16 and excluding SAM flags 2860). We did not exclude duplicate reads in order to avoid potential bias from increased duplicate-marking in individuals with high copy numbers. We also did not attempt to adjust copy number estimates for read-depth PCs, as measurement noise in locus-level copy number estimates is driven more by shot noise, and further optimization of accuracy was not necessary. We analyzed WGS data from male participants as well as females; data from males were useful for studying haploid copy number distributions and for helping to phase copy number estimates in females.

We counted reads that mapped to the following regions in GRCh38:

- *ICCE*: chrX:56767701–56781000. This 13.3kb region contains ∼2.5 copies of the core *ICCE* 5.4-kb repeat; its flanks contain diverged repeat units[63]. Reads originating from the copy-number-variable region of the *ICCE* repeat array align to the central 13.3kb region of highest homology.

- *DXZ4*: chrX:115721401–115889220. This 168-kb region, which contains the copies of the core 3.0-kb *DXZ4* repeat in GRCh38, spans a 100-kb assembly gap (Supplementary Fig. 44). Most of the *DXZ4* repeat array is assembled on the right side of the gap. A few repeat units are also situated immediately to the left of the gap, including a 5-kb sequence that is inverted relative to the rest of the array (Supplementary Fig. 44). The inversion probably reflects an assembly error, but reads from the *DXZ4* array also align to it, such that it was important to include in read-counting.

We used the following procedure to calibrate GC-corrected read counts to absolute estimates of repeat copy numbers (haploid for males, diploid sums for females):

1. Divide by the length of the repeat unit (5,400 bp for *ICCE*, 2,985 bp for *DXZ4*).
2. Divide by chromosome-level chrX coverage (computed as described in Methods). This accounted for decreases in read counts at *ICCE* and *DXZ4* caused by X loss in females; this adjustment was typically slight ($<1\%$), and *ICCE* and *DXZ4* copy number associated strongly with LOX levels regardless of whether the adjustment was applied. To limit outlier effects, we restricted analysis to females with chrX coverage between 0.8–1.05 (relative to expectation) and to males with chrX coverage between 0.95–1.05; values outside these ranges were very rare.
3. Fine-tune calibration:
    - *ICCE*: For males, divide by 1.043 and subtract 0.55; for females, divide by 1.035 and subtract 1.03 ($= 2\times0.515$).
      These values were chosen to calibrate copy number estimates to integer modes (Supplementary Fig. 36a), which required slight rescaling (by 1.035 and 1.043) and subtraction of an offset corresponding to the $\sim$0.5-unit partial repeat unit in the *ICCE* array. The same calibration produced integer modes in both UKB and AoU.
    - *DXZ4*: For males, divide by 1.043; for females, divide by 1.02.
      Copy number estimates of the much longer *DXZ4* array were not sufficiently accurate to distinguish integer modes, so in lieu of a direct means of fine-tuning calibration, we reused the calibration factor of 1.043 that we had estimated for males for *ICCE*, and we calibrated females to have mean copy number twice that of males. We did not apply a partial-repeat-unit offset as this comprised a negligible fraction of the *DXZ4* array (typically 10–120 repeat units per allele).

We statistically phased *ICCE* and *DXZ4* copy number estimates onto SNP-haplotypes using our previously described approach for phasing continuous-valued copy number estimates[126]. We generated the SNP-haplotypes by running SHAPEIT5[94] on a haplotype scaffold in UKB (as described in Supplementary Note 7) and on AoU SNP-array genotypes in chrX:110–120 Mb with MAF$>$0.01 and missingness$<$0.02.

##### 6.2 Paralogous sequence variant (PSV) copy number estimation

We were initially surprised to find that while *DXZ4* copy number associated strongly with LOX levels, the lead SNP at *DXZ4* had low correlation with *DXZ4* copy number and appeared to reflect an independent association. This led us to suspect that the lead SNP might tag sequence variation within the *DXZ4* repeat, which might also have a strong effect on X loss. We therefore undertook PSV profiling for both *ICCE* and *DXZ4* using our previously described approach for PSV copy number estimation[106].

Briefly, for each locus, we extracted reads that mapped to the repeat region in GRCh38 and realigned them to a reference sequence containing one copy of the repeat plus a 1-kb "wrap-around" sequence containing the beginning of a second copy of the same repeat; the wrap-around buffer allowed reads that spanned the ends of the repeat unit to be properly aligned. We identified PSVs based on observing non-reference alleles in these alignments. We estimated PSV allele fractions from relative allelic depths, and we obtained absolute estimates of PSV copy number by multiplying allele fractions by total copy number of the repeat (using copy number estimates refined by phasing, and adding a "bonus" 1 copy for males and 2 copies for females for PSV positions within the partial repeat unit of *ICCE*).

For *ICCE*, we used the 5,400-bp sequence chrX:56770001–56775400 as the reference sequence. For *DXZ4*, we used the 2,954-bp sequence chrX:115727764–115730717 as the reference sequence. This region is within the inverted portion of the *DXZ4* assembly in GRCh38 (Supplementary Fig. 44); while the inversion presumably reflects an assembly error, we used the inverted sequence as a reference for consistency with the strand orientation of the originally reported *DXZ4* monomer (GenBank S60754)[62].

##### 6.3 X loss association and fine-mapping analyses at *ICCE* and *DXZ4*

**Stepwise conditional analyses in UKB.** To fine-map associations of variation at *ICCE* and *DXZ4* with LOX levels, we performed stepwise conditional analyses at the *ICCE* and *DXZ4* loci, testing TOPMed-imputed SNP and indel variants, repeat copy number, and copy numbers of non-major PSV alleles for association with log-transformed, clipped LOX cell fractions. We analyzed female EUR-ancestry UKB participants using linear regression implemented in BOLT-LMM v2.5, including age, age$^2$, smoking status, and 20 genetic PCs as covariates.

At *ICCE*, copy number of the *ICCE* repeat produced the strongest association (Supplementary Fig. 38a), and conditioning on *ICCE* copy number greatly diminished the associations of nearby SNPs and indels ($\sim$10-fold; Supplementary Fig. 38b). Associations observed in the conditional analysis centered at the X chromosome centromere (Supplementary Fig. 38b). This may reflect an independent effect of centromeric variation on LOX that is still partially tagged by variation several megabases away owing to the particularly long centromere haplotype structure of the X chromosome[116]. We verified that this secondary association signal was robust to excluding the centromeric region from estimation of LOX cell fraction (specifically, by excluding chrX:47–70 Mb from the computation of chrX read-depth).

At *DXZ4*, copy number of the *DXZ4* repeat produced the strongest association (Supplementary Fig. 38c). After conditioning on *DXZ4* copy number, the top association among flanking

SNPs had $P$=5.0×10$^{-301}$, while the top two PSVs were chrX:115728524:G:A ($P$=9.1×10$^{-269}$) and chrX:115728581:T:C ($P$=1.6×10$^{-259}$; Supplementary Fig. 38d), with the latter being a clear candidate causal variant given its location within the CTCF binding motif of *DXZ4*. After conditioning on chrX:115728581:T:C, the lead variant was the YY1 motif-disrupting PSV represented by chrX:115728322:T:G (Supplementary Fig. 38e), and little signal remained after further conditioning on chrX:115728322:T:G (Supplementary Fig. 38f).

**Replication in AoU.** We replicated the associations of *ICCE* and *DXZ4* copy number and PSV copy number with LOX levels in female AoU participants. We also included *DXZ4* wild-type copy number (defined as total *DXZ4* copy number minus the copy numbers of the PSVs represented by chrX:115728581:T:C and chrX:115728322:T:G) in the association tests. We ran linear regression on log-transformed, clipped LOX cell fractions using BOLT-LMM v2.5 with the same covariates and variant filters as in our primary analyses. We ran analyses within each of the EUR, AMR, and AFR ancestry groups in AoU (Supplementary Fig. 37) as well as on all three ancestries combined (Fig. 4a). We restricted each analysis to unrelated individuals, prioritizing individuals to retain based on older age.

#### 6.4 Allelic series at *ICCE* and *DXZ4* for haplotype-resolved X loss cell fractions

To further explore the relationship of LOX levels with *ICCE* copy number and copy numbers of *DXZ4* repeat types (with and without the PSVs disrupting the CTCF and YY1 binding sites in *DXZ4*), and to assess the extent to which allelic variation associated with loss of the homolog in *cis* (carrying the allele) or the homolog in *trans* (carrying an individual's other haplotype), we estimated allelic series for haplotype-resolved LOX levels in female EUR-ancestry UKB participants. We phased PSV copy number estimates onto SNP haplotypes in the same way that we phased total copy number estimates[126], and we partitioned LOX cell fractions into haplotype-specific estimates as described in Methods.

For *ICCE*, we used the following procedure to compute an allelic series of effect sizes of common *ICCE* allele lengths. We fit a linear model regressing haploid LOX levels (log(max(0.001, LOX cell fraction))) on *ICCE* copy number in *cis* (as a categorical variable), *ICCE* copy number in *trans* (to evaluate the possibility of an effect of an allele on loss of the other homolog), and age, age$^2$, smoking status, and 20 genetic PCs as covariates. In this regression, each individual appeared twice, once for each haplotype. We clipped *ICCE* allele lengths to the copy number range [0, 14] and then rounded copy numbers to the nearest integer. In the linear model, we included an indicator variable for each value other than the mode (copy number 6). These analyses demonstrated a *cis*-specific linear relationship of *ICCE* copy number with log-transformed, clipped LOX cell fraction (Fig. 4d).

We used a similar approach to compute *cis* and *trans* allelic series for *DXZ4*, now subdividing *DXZ4* repeats into allelic copy numbers of wild-type *DXZ4* repeat units, *DXZ4* repeats with the chrX:115728581:T:C PSV ("CTCF-mut"), and *DXZ4* repeats with the chrX:115728322:T:G PSV ("YY1-mut"). We observed that the two PSVs were almost never observed on the same allele; for example, in AoU:

- 137,798 male AoU participants
- 115,845 estimated to have 2+ copies of the CTCF-mut PSV
- 13,420 estimated to have 2+ copies of the YY1-mut PSV
- 49 estimated to have 2+ copies of both.

As such, we partitioned copy numbers of *DXZ4* alleles into wild-type copies, CTCF-mut copies, and YY1-mut copies (assuming that CTCF-mut and YY1-mut PSVs almost never appear on the same repeat unit), and for each of the three *DXZ4* repeat types, we included copy number of the repeat type on the allele in *cis* as a categorical variable. We also included the number of wild-type copies in *trans* as a categorical variable (again to evaluate the possibility of an effect of an allele on loss of the other homolog). We clipped wild-type copy number to the range [0, 120], CTCF-mut copy number to the range [0, 50], and YY1-mut copy number to the range [0, 30], after which we binned copy numbers into 5-copy ranges. We included indicator variables for each bin other than the modal bin ([50, 55) for wild-type repeats and [0, 5) for the CTCF-mut and YY1-mut repeat types).

These analyses demonstrated a *cis*-specific nonlinear relationship of *DXZ4* wild-type copy number with log-transformed, clipped LOX cell fraction (Fig. 4e). The copy numbers of the CTCF-mut and YY1-mut PSVs associated with near-zero but slightly negative effects (Fig. 4e). We are unsure of the interpretation of this latter observation, but one hypothesis could be that interrupting the contiguity of CTCF/YY1-binding *DXZ4* copies has an effect (e.g., if *DXZ4* copy number affects missegregation of the inactive X chromosome due to some physical property relating to CTCF/YY1-density).

##### 6.5 CTCF ChIP-seq data analyses

To test whether the paralogous sequence variant (PSV) predicted to affect the CTCF binding site in *DXZ4* associates with decreased CTCF binding, we analyzed previously generated CTCF ChIP-seq data sets, counting the number of ChIP-seq reads arising from *DXZ4* repeat units with each of the two alleles. If CTCF binds less to *DXZ4* repeat units containing the motif-disrupting alternate allele (C), the allelic representation of the reference allele (T/(C+T)) should be higher in CTCF ChIP-seq data relative to whole-genome sequencing data (as allelic depths in WGS reflect the numbers of *DXZ4* repeat units with each allele in an individual's genome). (Here, the reference and alternate alleles are in the strand orientation of the original monomer[62], with the PSV represented by chrX:115728581:T:C in GRCh38; Supplementary Fig. 44.)

We identified 33 female 1KGP participants for whom we could measure both allelic ratios: for these individuals, CTCF ChIP-seq data was available from previously sequenced lymphoblastoid cell line (LCL) libraries[127,128] and high-coverage WGS data was also available[58]. For CTCF ChIP-seq data, we realigned FASTQ reads to GRCh38 using samtools[129] (v1.21) and bwa[130] (v0.7.18), and for both ChIP-seq and WGS, we summed allelic depths across PSV positions within GRCh38, reverse-complementing alleles as necessary according to strand orientation of repeats in GRCh38 (Supplementary Fig. 44). We computed allelic depths at each PSV position using htsbox pileup (r345; `https://github.com/lh3/htsbox`) with a minimum base quality of 20 and minimum query length of 30 bp. Counts of reads with each allele were merged across available CTCF ChIP-seq replicates. Allelic ratios in CTCF ChIP-seq and WGS were compared using a paired t-test. This identified a significant over-representation of the reference allele in CTCF ChIP-seq ($P$=0.012; Supplementary Fig. 38g), supporting the hypothesis that the alternate allele decreases CTCF binding to *DXZ4*, and that CTCF binding is involved in the mechanism by which wild-type *DXZ4* copy number associates with LOX.

We note that the ratio of reads with each allele in WGS reflects contributions from both X chromosomes, whereas only *DXZ4* repeats on the inactive X chromosome are bound by CTCF[131]. For samples with approximately equal numbers of cells with each X chromosome inactive, the allelic ratios we computed from CTCF ChIP-seq and WGS can be directly compared as we did. However, some LCLs have a bias toward one inactive X chromosome[132], such that the allelic ratio in CTCF ChIP-seq should ideally be compared against an allelic ratio closer to the allelic representation of *DXZ4* repeats on this X chromosome, and the WGS-derived ratio (based on both X chromosomes) may be too high or too low. This may explain why some samples exhibited higher reference allele fractions in WGS versus CTCF ChIP-seq (Supplementary Fig. 38g).

##### 6.6 CTCF and YY1 motif analyses

To generate coverage plots for CTCF and YY1 binding at *DXZ4* (Fig. 4f,h), we analyzed ChIP-seq data from ENCODE[128] generated using the K562 cell line (as YY1 ChIP-seq data from NA12878 LCLs did not appear to have peaks at *DXZ4* despite previous evidence of YY1 binding[133]). We realigned ChIP-seq reads to GRCh38 and computed per-base depth using mosdepth[119] v0.3.9 and merged two available replicates for each transcription factor target.

Best-matched motifs for each factor in *DXZ4* (`M08367_3.00` for CTCF and `M08315_3.00` for YY1) were identified using the CIS-BP database[134]. Relative position weight plots (Fig. 4g,i) were generated using the R package ggseqlogo[135] v0.1.

#### 7 Single-cell analysis of X chromosome inactivation in OneK1K

We analyzed single-cell RNA sequencing (scRNA-seq) data and SNP-array genotypes for 565 female participants in OneK1K[67] to assess whether or not allelic bias in X loss could be explained by potential allelic effects on skewing of X chromosome inactivation (XCI) as previously proposed[13].

##### 7.1 Phasing and imputing X chromosome haplotypes

To estimate phased genotypes of OneK1K participants at common variants across the X chromosome (including those without SNP-array genotypes), we used the following procedure. First, we downloaded the publicly available SNP-array data from NCBI Gene Expression Omnibus (GEO) accession GSE196829 and performed QC on the SNP-array genotypes. Specifically, we dropped variants with missingness>0.05, Hardy-Weinberg $P<1\times10^{-6}$, MAF<0.01, or allele frequency deviation >0.05 versus gnomAD `AF_nfe_XX` (allele frequency in non-Finnish European female individuals). On the X chromosome, 21,998 out of 29,191 variants remained after QC.

We used the TOPMed imputation server[84] to impute from SNP-array genotypes to genome-wide variants, also obtaining provisional phase information (from Eagle2[95]) for the imputed variants. This phase information was locally accurate but imperfect; for example, because phasing and imputation were performed on 10-Mb chunks, phase was randomized across chunk boundaries.

To improve phase accuracy, we used the UKB cohort as a phasing reference panel. To do so, we first phased X chromosomes in UKB by running SHAPEIT5 on WGS-derived genotypes for the X chromosome variants from the OneK1K SNP-array that remained after QC. We used the phased haplotypes as a reference panel to phase OneK1K, obtaining accurate long-range phase for variants on the SNP-array scaffold. We then integrated this phase information into the dense TOPMed-imputed genotypes, flipping phase as necessary to be consistent with UKB-phased haplotype scaffold.

Finally, to obtain accurate phase across the entire X chromosome, we incorporated information from single-cell XCI analyses (described in the next section). These analyses provided information about the phase of a small set of heterozygous variants across the X chromosome (varying across individuals) that were covered by scRNA-seq reads. We integrated this phase information to obtain a final set of dense haplotypes by flipping phase as necessary to obtain consistency with the scRNA-seq-determined phase.

##### 7.2 Calling X chromosome inactivation direction in single cells

To call XCI direction in single cells (i.e., which of the two homologous X chromosomes is inactivated in a given cell), we first downloaded raw scRNA-seq data in FASTQ format from GEO (accession number GSE196830), aligned the scRNA-seq data to GRCh38 using STARsolo[136], and identified single nucleotide variants (SNVs) in the aligned reads.

For each donor, we ran an iterative algorithm to phase highly expressed heterozygous sites on the X chromosome and assign XCI direction to single cells. Since only one of the two X chromosomes is activated in any single cell, germline alleles present in the transcripts of a single cell are all from the same X chromosome (haplotype) with the exception of transcripts of the minority of genes that escape XCI[68]. As such, scRNA-seq provides a source of phase information across the X chromosome, and conversely, knowledge of phase information allows XCI direction to be assessed in single cells[137].

For each individual, we first identified germline heterozygous sites with variant allele frequency (VAF) in the range [0.1, 0.9] (in scRNA-seq data aggregated across all cells) and with unique molecular identifier (UMI) counts of at least 5 for both the reference and alternate alleles. We then iteratively built a pair of haplotypes comprising a set of heterozygous sites with known phase relative to each other, updating provisional assignments of XCI direction per cell in each iteration.

We initialized the algorithm by identifying an "anchor" site selected to be maximally informative for XCI assignment. For each candidate site, we calculated a log-likelihood ratio (LLR) score evaluating how well its allelic distribution across all cells fit a model of monoallelic expression versus a diploid model (escaping XCI), incorporating Phred-scaled base quality scores to account for sequencing errors. We selected the site with the highest LLR as the "anchor" for the two haplotypes to be built. We defined haplotype 1 to contain the reference allele at this anchor site and haplotype 2 to contain the alternate allele.

We then performed the following two-step iteration:

1. **Assign cells to haplotypes.** For each cell, we calculated a log-likelihood score for each of the two haplotypes being inactivated based on the observed UMI counts and base qualities. We assigned the cell to a haplotype if the LLR comparing the two haplotype scores exceeded a threshold of 2.5. Cells not meeting this threshold were considered to have ambiguous XCI direction.

2. **Augment haplotypes with an additional heterozygous variant.** We selected an unphased heterozygous site to add to the haplotype based on having the most significant association between the allele observed in an scRNA-seq read (UMI) and the haplotype assignment of the cell from which the read originated. Specifically, for each candidate site, we constructed a $2 \times 2$ contingency table summarizing the UMI counts for the reference and alternate alleles of the candidate site against the cells-of-origin assigned to haplotype 1 versus haplotype 2, and we computed the Fisher's exact test $P$-value for this table. If at least one site had $P<0.05$, we augmented the haplotype with the variant with the lowest $P$-value; otherwise, we terminated the algorithm.

This procedure yielded a set of phased heterozygous sites for each individual along with an assignment of XCI status for 79% of cells (excluding those with ambiguous XCI direction, either due to low coverage at the phased sites or observations of alleles with inconsistent phase). For 91% of individuals, we determined XCI status in >500 cells. Skewed XCI within an individual was common, as previously observed[68]; 25% of individuals had >70% of cells with the majority haplotype inactivated (i.e., skewness >0.2) (Supplementary Fig. 40a). The amount of skewing did not associate with age ($P$=0.43; Supplementary Fig. 40a).

##### 7.3 Testing for allelic bias in XCI skew direction

To test whether any variants on the X chromosome exhibited evidence of biased X chromosome inactivation (i.e., a tendency for XCI to skew in favor of one allele over the other, which could generate allelic biases in X loss[13]), we performed an association analysis similar to "allelic shift" tests for LOX[13,14], making use of the chromosome-wide phasing and XCI skewing data we obtained from the previous analyses. For each individual $i$, let $y_i$ be the fraction of cells expressing haplotype 1 minus 0.5. To test a given variant for allelic bias in XCI skew direction, we restricted to individuals heterozygous for the variant. Among such individuals $i$, let $g_i$ be $\pm 1$ according to whether haplotype 1 carries the reference or the alternate allele. With these definitions, we computed a $z$-test to test whether the mean of $g_i \cdot y_i$ (i.e., the amount of XCI skew, oriented according to the phase of the variant under consideration) was different from zero.

We ran this test on 239,054 imputed and array-genotyped variants on the X chromosome with minor allele count $\geq$20 and did not observe any alleles significantly associated with the direction of XCI skewing (Supplementary Fig. 40b). We looked more closely at the *MIR223HG–VSIG4* and *ITM2A* loci, which showed the strongest allelic shifts in X loss (Fig. 5d and Supplementary Fig. 40c,d) after *ICCE* and *DXZ4* (which probably affect missegregation rather than XCI), but we did not observe any evidence of allelic biases in XCI skewing despite having reasonable statistical power at these two loci (Supplementary Fig. 40e). However, these results do not preclude the possibility of weaker allelic effects on XCI skewing that we were underpowered to observe; larger data sets will be needed to further evaluate this hypothesis.

##### 7.4 No evidence of cells with loss of the active X chromosome

Lastly, we analyzed the single-cell RNA-seq data from OneK1K to evaluate the extent to which X loss in females is restricted to the inactive X chromosome. X loss appears not to be tolerated in peripheral leukocytes in males, suggesting that cells may require an active X chromosome to survive[138]. However, previous literature has been uncertain about the extent to which X loss in females affects the inactive X: studies have suggested that the inactive X chromosome is "preferentially" lost[6,139,140] and left open the possibility that mosaic LOX could involve cell populations with loss of either the active or the inactive X chromosome, providing differing fitness benefits[72].

Here, single-cell data from OneK1K (an older cohort likely to contain several individuals with moderate- to high-cell-fraction LOX) provided an opportunity to more definitively evaluate this question. To do so, we first ran MoChA[14,15] (version 2024-09-27) on the OneK1K SNP-array intensity data (using TOPMed-phased haplotypes) to identify female donors with substantial levels of LOX. The top 10 individuals had estimated LOX cell fractions of 2.8%–37.5%, such that their scRNA-seq data was expected to contain tens to hundreds of cells with LOX.

We ran two analyses of scRNA-seq data to see whether any cells from these 10 female donors with highest LOX cell fractions appeared to have lost the active X chromosome. First, we searched for cells with abnormally low expression of genes that do not escape XCI. Any cell with loss of the active X chromosome would have near-zero expression of these non-escape genes. To adjust for varying amounts of transcription in different cells, we computed (for each cell) the ratio of the combined expression level (number of UMIs) of non-escape genes to the combined expression level of all genes on the X chromosome. We used the classification of genes as escaping vs. not escaping XCI previously reported by Tukiainen et al. 2017 (Supplementary Table 13)[68], comprising 89 escape genes and 386 non-escape genes. This analysis did not detect any cells with unusually low expression of non-escape genes, suggesting that in these 10 donors with 2.8%–37.5% LOX, all sequenced cells with LOX had lost the inactive X chromosome (Supplementary Fig. 41; blue histograms).

Second, we performed gene-level analyses counting (for each cell) the number of non-escape genes with nonzero expression (at least 1 UMI). Again, in every sequenced cell of each of the 10 donors, expression was observed from at least one non-escape gene; transcripts from several non-escape genes were observed in nearly all cells (Supplementary Fig. 41; red histograms).

These results suggest that loss of the active X chromosome in leukocytes is either not tolerated or substantially reduces clonal fitness, such that only leukocytes with loss of the inactive X are present at observable frequencies in blood cell populations.

### 8 Association analyses of centromeric, *ICCE*, and *DXZ4* structural variants

We tested the hybrid chromosome 13/22 tag SNP (22:15432903:C:T), *ICCE* copy number, and *DXZ4* copy number for association with 3,447 binary phenotypes curated in the "All by All"[141] analysis of AoU v8. To minimize potential confounders, we restricted analysis to EUR-ancestry individuals (n=35,228–176,550 depending on phenotype and genotype missingness). We ran association tests using logistic regression with Firth correction implemented in the logistf R package (v1.26.1), including age, age$^2$, sex, and 16 genetic PCs as covariates. We encoded *ICCE* and *DXZ4* copy number genotypes as diploid for females (i.e., total copy number across the two haplotypes) and haploid for males. These analyses identified a single significant association ($P<0.05/(3\times3{,}447)$) between *DXZ4* copy number and self-reported history of hemorrhoids from the Personal and Family Health History (PFHH) survey ($P=3.1\times10^{-10}$; n=130,070; 32,152 cases and 97,918 controls).

We followed up on the *DXZ4*-hemorrhoids association by performing GWAS on hemorrhoids in AoU v8, restricting to unrelated EUR-ancestry individuals with PFHH survey data and blood-derived WGS data (such that *DXZ4* copy number estimates were available from our read-depth analyses). We pruned to unrelated individuals based on the AoU v8 sample relatedness file (containing relationship pairs with kinship coefficient >0.1), prioritizing individuals to retain based on case status and then on older age; this left 125,764 individuals (31,832 cases and 93,932 controls). We ran linear regression using BOLT-LMM with age, $age^2$, sex, smoking status, and 16 genetic PCs as covariates, analyzing SNP and indel variants with MAF>0.1% and *ICCE*, *DXZ4* ($P$=$1.8\times10^{-9}$), and *DXZ4* wild-type copy number (Supplementary Fig. 43a). We also ran an analogous analysis in which we restricted to females (n=79,220; 20,684 cases and 58,536 controls; $P$=$1.1\times10^{-9}$, Supplementary Fig. 43b). We observed no association of *DXZ4* copy number with hemorrhoids in males ($P$=0.22; n=47,887, 11,240 cases and 36,647 controls).

We replicated the *DXZ4*-hemorrhoids association in 43,684 additional EUR-ancestry female AoU participants (12,631 cases and 31,053 controls who completed the PFHH survey). These individuals were not included in our initial analysis either because they had saliva-derived WGS data (which we did not include in genome-wide read-depth profiling) or because they were subsequently included in the AoU v9 release. We estimated *DXZ4* copy number for these individuals in two independent ways: (1) by counting reads aligned to the *DXZ4* repeat region in GRCh38 and normalizing against genome-wide coverage; and (2) by imputing from our primary data set (the v8 blood-derived WGS; n=349,468 with *DXZ4* copy number estimates) using shared SNP-haplotypes[126]. We tested *DXZ4* copy number estimates for association with hemorrhoids adjusting for age, $age^2$, and 16 genetic PCs and observed that the association replicated ($P$=0.014 for (1), $P$=0.050 for (2), in each case with a risk-reducing effect direction).

Finally, we tested LOX cell fraction (clipped to the range 0–5%) for association with hemorrhoids. We ran linear regression with age, $age^2$, and 16 genetic PCs as covariates and did not observe an association ($P$=0.30) in EUR-ancestry female AoU v8 participants (n=80,218).

### References

1. Jacobs, P. A., Court Brown, W. & Doll, R. Distribution of human chromosome counts in relation to age. *Nature* **191**, 1178–1180 (1961).

2. Jacobs, P. A., Brunton, M., Court Brown, W., Doll, R. & Goldstein, H. Change of human chromosome count distributions with age: evidence for a sex difference. *Nature* **197**, 1080–1081 (1963).

3. Jacobs, K. B. *et al.* Detectable clonal mosaicism and its relationship to aging and cancer. *Nature Genetics* **44**, 651–658 (2012).

4. Laurie, C. C. *et al.* Detectable clonal mosaicism from birth to old age and its relationship to cancer. *Nature Genetics* **44**, 642–650 (2012).

5. Forsberg, L. A. *et al.* Mosaic loss of chromosome Y in peripheral blood is associated with shorter survival and higher risk of cancer. *Nature Genetics* **46**, 624–628 (2014).

6. Machiela, M. J. *et al.* Female chromosome X mosaicism is age-related and preferentially affects the inactivated X chromosome. *Nature Communications* **7**, 11843 (2016).

7. Dumanski, J. P. *et al.* Mosaic loss of chromosome Y in blood is associated with Alzheimer disease. *The American Journal of Human Genetics* **98**, 1208–1219 (2016).

8. Sano, S. *et al.* Hematopoietic loss of Y chromosome leads to cardiac fibrosis and heart failure mortality. *Science* **377**, 292–297 (2022).

9. Sato, G. *et al.* Genetic regulation across germline and somatic variation on the Y chromosome contributes to type 2 diabetes. *Nature Medicine* **32**, 894–905 (2026).

10. Bruhn-Olszewska, B. *et al.* The effects of loss of Y chromosome on male health. *Nature Reviews Genetics* **26**, 320–335 (2025).

11. Thompson, D. J. *et al.* Genetic predisposition to mosaic Y chromosome loss in blood. *Nature* **575**, 652–657 (2019).

12. Francis, M. *et al.* Multi-ancestry genome-wide association meta-analysis of mosaic loss of chromosome Y in the Million Veteran Program identifies 240 novel loci. *medRxiv* (2024).

13. Liu, A. *et al.* Genetic drivers and cellular selection of female mosaic X chromosome loss. *Nature* **631**, 134–141 (2024).

14. Loh, P.-R. *et al.* Insights into clonal haematopoiesis from 8,342 mosaic chromosomal alterations. *Nature* **559**, 350–355 (2018).

15. Loh, P.-R., Genovese, G. & McCarroll, S. A. Monogenic and polygenic inheritance become instruments for clonal selection. *Nature* **584**, 136–141 (2020).

16. Terao, C. *et al.* Chromosomal alterations among age-related haematopoietic clones in Japan. *Nature* **584**, 130–135 (2020).

17. Tang, D., Kamitaki, N., Mukamel, R. E., Rubinacci, S. & Loh, P.-R. Patterns and drivers of 43,617 mosaic chromosomal alterations in blood. *Nature Genetics* **58**, 1308–1319 (2026).

18. Zhao, K. *et al.* Genetic drivers and clinical consequences of mosaic chromosomal alterations in 1 million individuals. *medRxiv* (2025).

19. Mitchell, E. *et al.* Clonal dynamics of haematopoiesis across the human lifespan. *Nature* **606**, 343–350 (2022).

20. Jakubek, Y. A. *et al.* Mosaic chromosomal alterations in blood across ancestries using whole-genome sequencing. *Nature Genetics* **55**, 1912–1919 (2023).

21. Worrall, J. T. *et al.* Non-random mis-segregation of human chromosomes. *Cell Reports* **23**, 3366–3380 (2018).

22. Dumont, M. *et al.* Human chromosome-specific aneuploidy is influenced by DNA-dependent centromeric features. *EMBO Journal* **39**, e102924 (2020).

23. Klaasen, S. J. *et al.* Nuclear chromosome locations dictate segregation error frequencies. *Nature* **607**, 604–609 (2022).

24. Logsdon, G. A. *et al.* The variation and evolution of complete human centromeres. *Nature* **629**, 136–145 (2024).

25. Logsdon, G. A. *et al.* Complex genetic variation in nearly complete human genomes. *Nature* **644**, 430–441 (2025).

26. Gao, S. *et al.* A global view of human centromere variation and evolution. *Nature* (2026).

27. UK Biobank Whole-Genome Sequencing Consortium *et al.* Whole-genome sequencing of 490,640 UK Biobank participants. *Nature* **645**, 692–701 (2025).

28. The All of Us Research Program Genomics Investigators. Genomic data in the All of Us Research Program. *Nature* **627**, 340–346 (2024).

29. Jakubek, Y. A. *et al.* Genomic and phenotypic correlates of mosaic loss of chromosome Y in blood. *The American Journal of Human Genetics* **112**, 276–290 (2025).

30. Dumanski, J. P. *et al.* Smoking is associated with mosaic loss of chromosome Y. *Science* **347**, 81–83 (2015).

31. Zhou, W. *et al.* Mosaic loss of chromosome Y is associated with common variation near *TCL1A*. *Nature Genetics* **48**, 563–568 (2016).

32. Lin, S.-H. *et al.* Incident disease associations with mosaic chromosomal alterations on autosomes, X and Y chromosomes: insights from a phenome-wide association study in the UK Biobank. *Cell & Bioscience* **11**, 143 (2021).

33. Yang, J. *et al.* Common SNPs explain a large proportion of the heritability for human height. *Nature Genetics* **42**, 565–569 (2010).

34. Ge, T., Chen, C.-Y., Neale, B. M., Sabuncu, M. R. & Smoller, J. W. Phenome-wide heritability analysis of the UK Biobank. *PLoS Genetics* **13**, e1006711 (2017).

35. Sacristan, C. *et al.* Vertebrate centromeres in mitosis are functionally bipartite structures stabilized by cohesin. *Cell* **187**, 3006–3023 (2024).

36. Bick, A. G. *et al.* Inherited causes of clonal haematopoiesis in 97,691 whole genomes. *Nature* **586**, 763–768 (2020).

37. Kar, S. P. *et al.* Genome-wide analyses of 200,453 individuals yield new insights into the causes and consequences of clonal hematopoiesis. *Nature Genetics* **54**, 1155–1166 (2022).

38. Kessler, M. D. *et al.* Common and rare variant associations with clonal haematopoiesis phenotypes. *Nature* **612**, 301–309 (2022).

39. Stacey, S. N. *et al.* Genetics and epidemiology of mutational barcode-defined clonal hematopoiesis. *Nature Genetics* **55**, 2149–2159 (2023).

40. Poeschla, M. & Sankaran, V. G. Genetic influences on haematopoiesis. *Nature Reviews Genetics* **27**, 603–621 (2026).

41. Chen, C.-Z., Li, L., Lodish, H. F. & Bartel, D. P. MicroRNAs modulate hematopoietic lineage differentiation. *Science* **303**, 83–86 (2004).

42. Schmitz, R. *et al.* *TNFAIP3* (A20) is a tumor suppressor gene in Hodgkin lymphoma and primary mediastinal B cell lymphoma. *Journal of Experimental Medicine* **206**, 981–989 (2009).

43. Weinstock, J. S. *et al.* Aberrant activation of *TCL1A* promotes stem cell expansion in clonal haematopoiesis. *Nature* **616**, 755–763 (2023).

44. Zhao, Y. *et al.* *GIGYF1* loss of function is associated with clonal mosaicism and adverse metabolic health. *Nature Communications* **12**, 4178 (2021).

45. Fujita, Y. *et al.* Priming of centromere for CENP-A recruitment by human hMis18$\alpha$, hMis18$\beta$, and M18BP1. *Developmental Cell* **12**, 17–30 (2007).

46. Yatskevich, S. *et al.* Structure of the human inner kinetochore bound to a centromeric CENP-A nucleosome. *Science* **376**, 844–852 (2022).

47. Polley, S. *et al.* Structure of the human KMN complex and implications for regulation of its assembly. *Nature Structural & Molecular Biology* **31**, 861–873 (2024).

48. Yatskevich, S., Yang, J., Bellini, D., Zhang, Z. & Barford, D. Structure of the human outer kinetochore KMN network complex. *Nature Structural & Molecular Biology* **31**, 874–883 (2024).

49. Kixmoeller, K., Tarasovetc, E. V., Mer, E., Chang, Y.-W. & Black, B. E. Centromeric chromatin clearings demarcate the site of kinetochore formation. *Cell* **188**, 1280–1296 (2025).

50. Ariyoshi, M. & Fukagawa, T. An updated view of the kinetochore architecture. *Trends in Genetics* **39**, 941–953 (2023).

51. McAinsh, A. D. & Kops, G. J. Principles and dynamics of spindle assembly checkpoint signalling. *Nature Reviews Molecular Cell Biology* **24**, 543–559 (2023).

52. Jaganathan, K. *et al.* Predicting expression-altering promoter mutations with deep learning. *Science* **389**, eads7373 (2025).

53. Waye, J. S. & Willard, H. F. Nucleotide sequence heterogeneity of alpha satellite repetitive DNA: a survey of alphoid sequences from different human chromosomes. *Nucleic Acids Research* **15**, 7549–7569 (1987).

54. Altemose, N. *et al.* Complete genomic and epigenetic maps of human centromeres. *Science* **376**, eabl4178 (2022).

55. Gershman, A. *et al.* Epigenetic patterns in a complete human genome. *Science* **376**, eabj5089 (2022).

56. Jaggi, K. E., Hoyt, S. J., O'Neill, R. J. & Sullivan, B. A. A genomic and epigenomic view of human centromeres. *Nature Reviews Genetics* (2026).

57. Aldrup-MacDonald, M. E., Kuo, M. E., Sullivan, L. L., Chew, K. & Sullivan, B. A. Genomic variation within alpha satellite DNA influences centromere location on human chromosomes with metastable epialleles. *Genome Research* **26**, 1301–1311 (2016).

58. Byrska-Bishop, M. *et al.* High-coverage whole-genome sequencing of the expanded 1000 Genomes Project cohort including 602 trios. *Cell* **185**, 3426–3440 (2022).

59. Salinas-Luypaert, C. *et al.* DNA methylation influences human centromere positioning and function. *Nature Genetics* **57**, 2509–2521 (2025).

60. Carty, B. L. *et al.* Heterochromatin boundaries maintain centromere position, size and number. *Nature Structural & Molecular Biology* **33**, 220–234 (2026).

61. Xu, Y. *et al.* Haplotype-resolved centromeric chromatin organization from a complete diploid human genome. *bioRxiv* (2026).

62. Giacalone, J., Friedes, J. & Francke, U. A novel GC-rich human macrosatellite VNTR in Xq24 is differentially methylated on active and inactive X chromosomes. *Nature Genetics* **1**, 137–143 (1992).

63. Horakova, A. H., Moseley, S. C., McLaughlin, C. R., Tremblay, D. C. & Chadwick, B. P. The macrosatellite *DXZ4* mediates CTCF-dependent long-range intrachromosomal interactions on the human inactive X chromosome. *Human Molecular Genetics* **21**, 4367–4377 (2012).

64. Rao, S. S. *et al.* A 3D map of the human genome at kilobase resolution reveals principles of chromatin looping. *Cell* **159**, 1665–1680 (2014).

65. Darrow, E. M. *et al.* Deletion of *DXZ4* on the human inactive X chromosome alters higher-order genome architecture. *Proceedings of the National Academy of Sciences* **113**, E4504–E4512 (2016).

66. Miga, K. H. *et al.* Telomere-to-telomere assembly of a complete human X chromosome. *Nature* **585**, 79–84 (2020).

67. Yazar, S. *et al.* Single-cell eQTL mapping identifies cell type–specific genetic control of autoimmune disease. *Science* **376**, eabf3041 (2022).

68. Tukiainen, T. *et al.* Landscape of X chromosome inactivation across human tissues. *Nature* **550**, 244–248 (2017).

69. Sanderson, E. *et al.* Mendelian randomization. *Nature Reviews Methods Primers* **2**, 6 (2022).

70. Hain, C., Rausch, T., Consortium, H. G. S. V., Consortium, H. P. R. & Korbel, J. O. HOROSCOPE: Decoding human centromere architecture from short reads using k-mer signatures. *bioRxiv* (2026).

71. Lu, S. *et al.* Pangenome-based human genome analysis improves trait association and genomic prediction. *bioRxiv* 2026–07 (2026).

72. Watson, C. J. & Blundell, J. R. Mutation rates and fitness consequences of mosaic chromosomal alterations in blood. *Nature Genetics* **55**, 1677–1685 (2023).

73. Halldorsson, B. V. *et al.* Characterizing mutagenic effects of recombination through a sequence-level genetic map. *Science* **363**, eaau1043 (2019).

74. Carioscia, S. A. *et al.* Common variation in meiosis genes shapes human recombination and aneuploidy. *Nature* **651**, 146–153 (2026).

75. Bycroft, C. *et al.* The UK Biobank resource with deep phenotyping and genomic data. *Nature* **562**, 203–209 (2018).

76. All of Us Research Program Investigators. The "All of Us" research program. *New England Journal of Medicine* **381**, 668–676 (2019).

77. Vattathil, S. & Scheet, P. Extensive hidden genomic mosaicism revealed in normal tissue. *The American Journal of Human Genetics* **98**, 571–578 (2016).

78. Handsaker, R. E. *et al.* Large multiallelic copy number variations in humans. *Nature Genetics* **47**, 296–303 (2015).

79. Rhie, A. *et al.* The complete sequence of a human Y chromosome. *Nature* **621**, 344–354 (2023).

80. Tuke, M. A. *et al.* Mosaic Turner syndrome shows reduced penetrance in an adult population study. *Genetics in Medicine* **21**, 877–886 (2019).

81. Hujoel, M. L. *et al.* Insights into DNA repeat expansions among 900,000 biobank participants. *Nature* **650**, 920–929 (2026).

82. Loh, P. R. *et al.* Efficient Bayesian mixed-model analysis increases association power in large cohorts. *Nature Genetics* **47**, 284–290 (2015).

83. Taliun, D. *et al.* Sequencing of 53,831 diverse genomes from the NHLBI TOPMed Program. *Nature* **590**, 290–299 (2021).

84. Das, S. *et al.* Next-generation genotype imputation service and methods. *Nature Genetics* **48**, 1284–1287 (2016).

85. Loh, P.-R., Kichaev, G., Gazal, S., Schoech, A. P. & Price, A. L. Mixed-model association for biobank-scale datasets. *Nature Genetics* **50**, 906–908 (2018).

86. Barton, A. R., Sherman, M. A., Mukamel, R. E. & Loh, P.-R. Whole-exome imputation within UK Biobank powers rare coding variant association and fine-mapping analyses. *Nature Genetics* **53**, 1260–1269 (2021).

87. Frankish, A. *et al.* GENCODE: Reference annotation for the human and mouse genomes in 2023. *Nucleic Acids Research* **51**, D942–D949 (2023).

88. GTEx Consortium. The GTEx Consortium atlas of genetic regulatory effects across human tissues. *Science* **369**, 1318–1330 (2020).

89. McLaren, W. *et al.* The Ensembl Variant Effect Predictor. *Genome Biology* **17**, 122 (2016).

90. Chen, S. *et al.* A genomic mutational constraint map using variation in 76,156 human genomes. *Nature* **625**, 92–100 (2024).

91. Spence, J. P. *et al.* Specificity, length and luck drive gene rankings in association studies. *Nature* **649**, 918–925 (2026).

92. Loh, P.-R. *et al.* Contrasting genetic architectures of schizophrenia and other complex diseases using fast variance-components analysis. *Nature Genetics* **47**, 1385–1392 (2015).

93. Bulik-Sullivan, B. *et al.* An atlas of genetic correlations across human diseases and traits. *Nature Genetics* **47**, 1236–1241 (2015).

94. Hofmeister, R. J., Ribeiro, D. M., Rubinacci, S. & Delaneau, O. Accurate rare variant phasing of whole-genome and whole-exome sequencing data in the UK Biobank. *Nature Genetics* **55**, 1243–1249 (2023).

95. Loh, P.-R. *et al.* Reference-based phasing using the Haplotype Reference Consortium panel. *Nature Genetics* **48**, 1443–1448 (2016).

96. Bowden, J., Davey Smith, G. & Burgess, S. Mendelian randomization with invalid instruments: effect estimation and bias detection through Egger regression. *International Journal of Epidemiology* **44**, 512–525 (2015).

97. Verbanck, M., Chen, C.-Y., Neale, B. & Do, R. Detection of widespread horizontal pleiotropy in causal relationships inferred from Mendelian randomization between complex traits and diseases. *Nature Genetics* **50**, 693–698 (2018).

98. Rivier, C. A. *et al.* Genal: a Python toolkit for genetic risk scoring and Mendelian randomization. *Bioinformatics Advances* **5**, vbae207 (2025).

99. 1000 Genomes Project Consortium *et al.* A global reference for human genetic variation. *Nature* **526**, 68 (2015).

100. Chang, C. C. *et al.* Second-generation PLINK: rising to the challenge of larger and richer datasets. *Gigascience* **4**, s13742–015 (2015).

101. Zhou, W. *et al.* Efficiently controlling for case-control imbalance and sample relatedness in large-scale genetic association studies. *Nature Genetics* **50**, 1335–1341 (2018).

102. Mbatchou, J. *et al.* Computationally efficient whole-genome regression for quantitative and binary traits. *Nature Genetics* **53**, 1097–1103 (2021).

103. Tournamille, C., Colin, Y., Cartron, J. P. & Le Van Kim, C. Disruption of a GATA motif in the *Duffy* gene promoter abolishes erythroid gene expression in Duffy–negative individuals. *Nature Genetics* **10**, 224–228 (1995).

104. Reich, D. *et al.* Reduced neutrophil count in people of African descent is due to a regulatory variant in the Duffy antigen receptor for chemokines gene. *PLoS Genetics* **5**, e1000360 (2009).

105. Gao, H. *et al.* The landscape of tolerated genetic variation in humans and primates. *Science* **380**, eabn8153 (2023).

106. Hujoel, M. L. *et al.* Protein-altering variants at copy number-variable regions influence diverse human phenotypes. *Nature Genetics* **56**, 569–578 (2024).

107. Genovese, G. *et al.* Clonal hematopoiesis and blood-cancer risk inferred from blood DNA sequence. *New England Journal of Medicine* **371**, 2477–2487 (2014).

108. Jaiswal, S. *et al.* Age-related clonal hematopoiesis associated with adverse outcomes. *New England Journal of Medicine* **371**, 2488–2498 (2014).

109. Brown, D. W. *et al.* Shared and distinct genetic etiologies for different types of clonal hematopoiesis. *Nature Communications* **14**, 5536 (2023).

110. Niroula, A. *et al.* Distinction of lymphoid and myeloid clonal hematopoiesis. *Nature Medicine* **27**, 1921–1927 (2021).

111. Mattisson, J. *et al.* Loss of chromosome Y in regulatory T cells. *BMC Genomics* **25**, 243 (2024).

112. Dawoud, A., Green, L. & Rackham, O. Loss of chromosome Y associates with altered immune cell trajectories and X-inactivation features. *Aging Cell* **25**, e70528 (2026).

113. Morales, J. *et al.* A joint NCBI and EMBL-EBI transcript set for clinical genomics and research. *Nature* **604**, 310–315 (2022).

114. Zeng, T. & Li, Y. I. Predicting RNA splicing from DNA sequence using Pangolin. *Genome Biology* **23**, 103 (2022).

115. Ward, L. D. & Kellis, M. HaploReg v4: systematic mining of putative causal variants, cell types, regulators and target genes for human complex traits and disease. *Nucleic Acids Research* **44**, D877–D881 (2016).

116. Langley, S. A., Miga, K. H., Karpen, G. H. & Langley, C. H. Haplotypes spanning centromeric regions reveal persistence of large blocks of archaic DNA. *eLife* **8**, e42989 (2019).

117. Lucas, J. K. *et al.* HPRC2: A human pangenome reference with near-complete coverage of common genetic variation. *bioRxiv* (2026).

118. Rhie, A. *et al.* Biobank-scale genotyping of Robertsonian translocations reveals hidden structural variation on the human acrocentric chromosomes. *bioRxiv* (2026).

119. Pedersen, B. S. & Quinlan, A. R. Mosdepth: quick coverage calculation for genomes and exomes. *Bioinformatics* **34**, 867–868 (2018).

120. Aganezov, S. *et al.* A complete reference genome improves analysis of human genetic variation. *Science* **376**, eabl3533 (2022).

121. Nurk, S. *et al.* The complete sequence of a human genome. *Science* **376**, 44–53 (2022).

122. Linardopoulou, E. V. *et al.* Human subtelomeres are hot spots of interchromosomal recombination and segmental duplication. *Nature* **437**, 94–100 (2005).

123. Guarracino, A. *et al.* Recombination between heterologous human acrocentric chromosomes. *Nature* **617**, 335–343 (2023).

124. Lin, J. *et al.* Human acrocentric chromosome short-arm *de novo* mutation and recombination. *Cell* **189**, 4876–4890 (2026).

125. Guarracino, A., Gyamfi, A., Consortium, H. P. R. & Garrison, E. Concerted evolution and unorthodox recombination of human subtelomeres. *bioRxiv* 2026–07 (2026).

126. Mukamel, R. E. *et al.* Protein-coding repeat polymorphisms strongly shape diverse human phenotypes. *Science* **373**, 1499–1505 (2021).

127. Ding, Z. *et al.* Quantitative genetics of CTCF binding reveal local sequence effects and different modes of X-chromosome association. *PLoS Genetics* **10**, e1004798 (2014).

128. The ENCODE Project Consortium *et al.* Expanded encyclopaedias of DNA elements in the human and mouse genomes. *Nature* **583**, 699–710 (2020).

129. Li, H. *et al.* The sequence alignment/map format and SAMtools. *Bioinformatics* **25**, 2078–2079 (2009).

130. Li, H. Aligning sequence reads, clone sequences and assembly contigs with BWA-MEM. *arXiv* (2013).

131. Chadwick, B. P. DXZ4 chromatin adopts an opposing conformation to that of the surrounding chromosome and acquires a novel inactive X-specific role involving CTCF and antisense transcripts. *Genome Research* **18**, 1259–1269 (2008).

132. Sauteraud, R. *et al.* Inferring genes that escape X-Chromosome inactivation reveals important contribution of variable escape genes to sex-biased diseases. *Genome Research* **31**, 1629–1637 (2021).

133. Moseley, S. C. *et al.* YY1 associates with the macrosatellite DXZ4 on the inactive X chromosome and binds with CTCF to a hypomethylated form in some male carcinomas. *Nucleic Acids Research* **40**, 1596–1608 (2012).

134. Weirauch, M. T. *et al.* Determination and inference of eukaryotic transcription factor sequence specificity. *Cell* **158**, 1431–1443 (2014).

135. Wagih, O. ggseqlogo: a versatile R package for drawing sequence logos. *Bioinformatics* **33**, 3645–3647 (2017).

136. Kaminow, B., Yunusov, D. & Dobin, A. STARsolo: Accurate, fast and versatile mapping/quantification of single-cell and single-nucleus RNA-seq data. *bioRxiv* (2021).

137. Tomofuji, Y. *et al.* Quantification of escape from X chromosome inactivation with single-cell omics data reveals heterogeneity across cell types and tissues. *Cell Genomics* **4** (2024).

138. Zhou, W. *et al.* Detectable chromosome X mosaicism in males is rarely tolerated in peripheral leukocytes. *Scientific Reports* **11**, 1193 (2021).

139. Abruzzo, M., Mayer, M. & Jacobs, P. Aging and aneuploidy: evidence for the preferential involvement of the inactive X chromosome. *Cytogenetic and Genome Research* **39**, 275–278 (1985).

140. Tucker, J. D., Nath, J. & Hando, J. C. Activation status of the X chromosome in human micronucleated lymphocytes. *Human Genetics* **97**, 471–475 (1996).

141. Lu, W. *et al.* Systematic common and rare variant association testing in 392,030 whole genomes in All of Us. *medRxiv* (2026).

142. Altschul, S. F., Gish, W., Miller, W., Myers, E. W. & Lipman, D. J. Basic local alignment search tool. *Journal of Molecular Biology* **215**, 403–410 (1990).

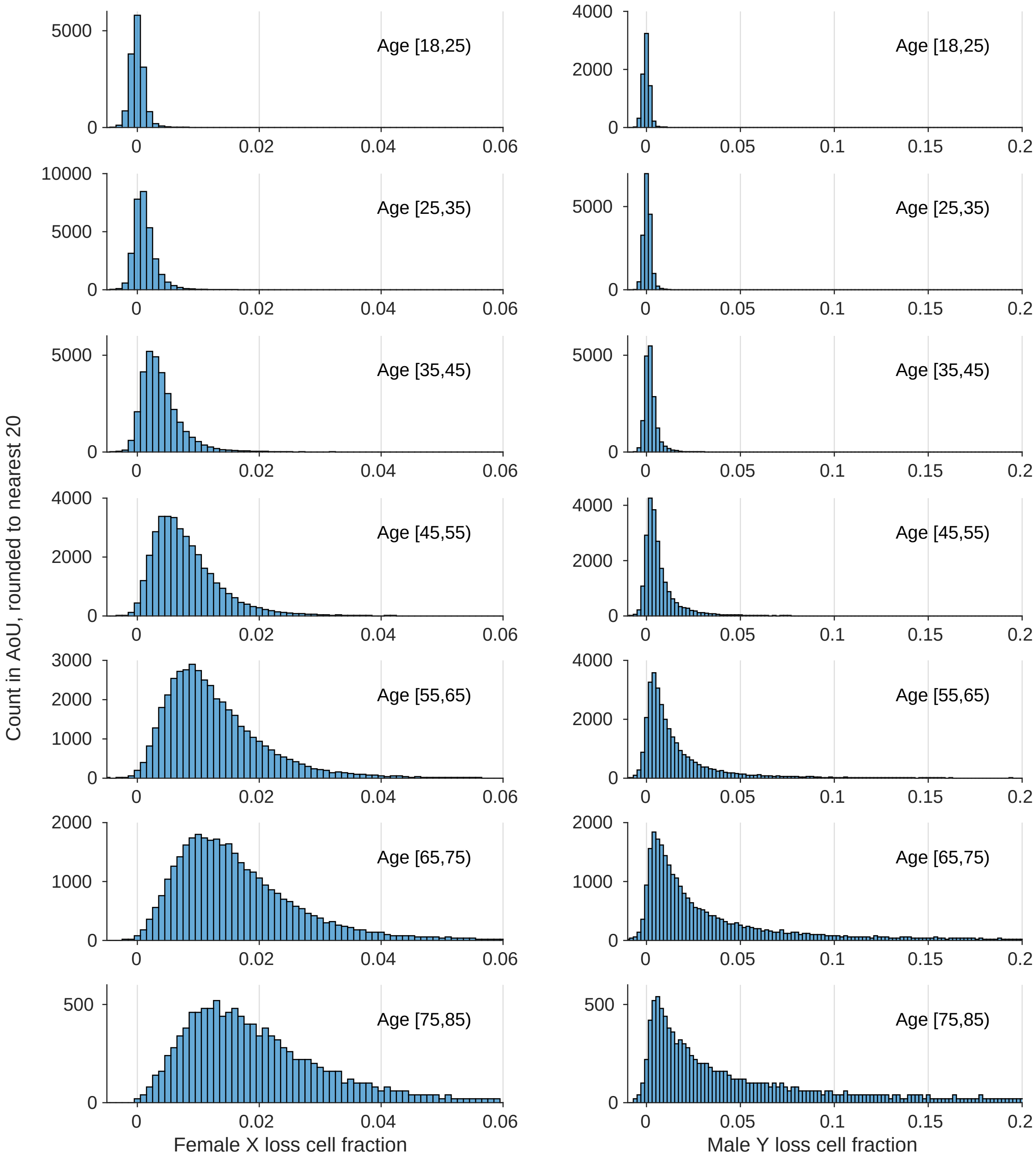

**Supplementary Figure 1. Distributions of estimated fractions of white blood cells harboring sex chromosome losses, across age tranches in AoU.** Left, X loss in females (n=205,620); right, Y loss in males (n=136,220). X loss and Y loss cell fractions both shift rightward with increasing age, but the distribution of Y loss cell fraction has a much longer tail, indicating greater propensity for clonal expansion. Negative estimates are plotted as negative values (rather than truncating them at 0) to provide an indication of the amount of measurement noise in these estimates derived from WGS read-depth. Participant counts were rounded to the nearest 20.

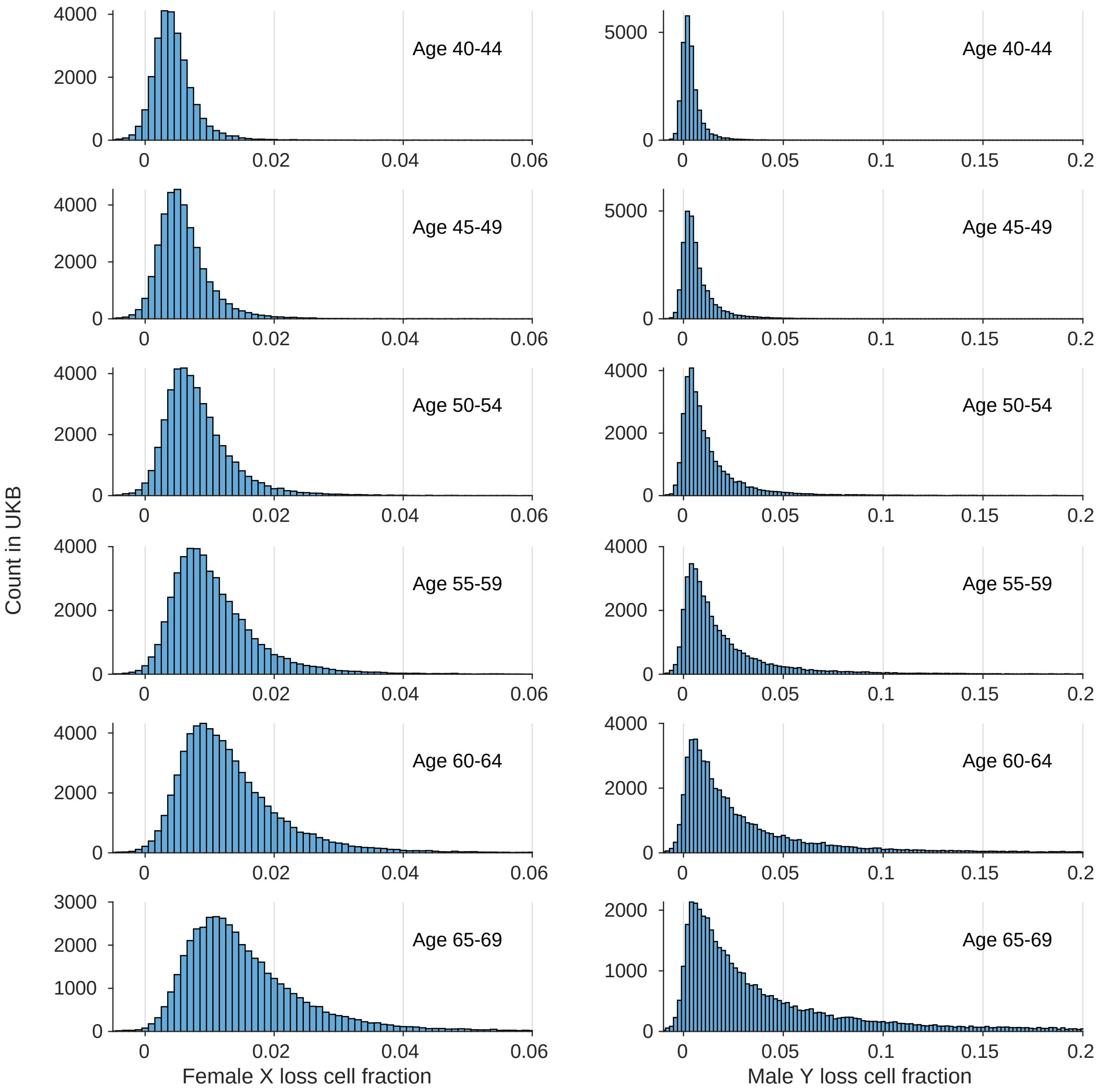

**Supplementary Figure 2. Distributions of estimated fractions of white blood cells harboring sex chromosome losses, across age tranches in UKB.** Left, X loss in females (n=259,088); right, Y loss in males (n=221,857). X loss and Y loss cell fractions both shift rightward with increasing age, but the distribution of Y loss cell fraction has a much longer tail, indicating greater propensity for clonal expansion. Negative estimates are plotted as negative values (rather than truncating them at 0) to provide an indication of the amount of measurement noise in these estimates derived from WGS read-depth.

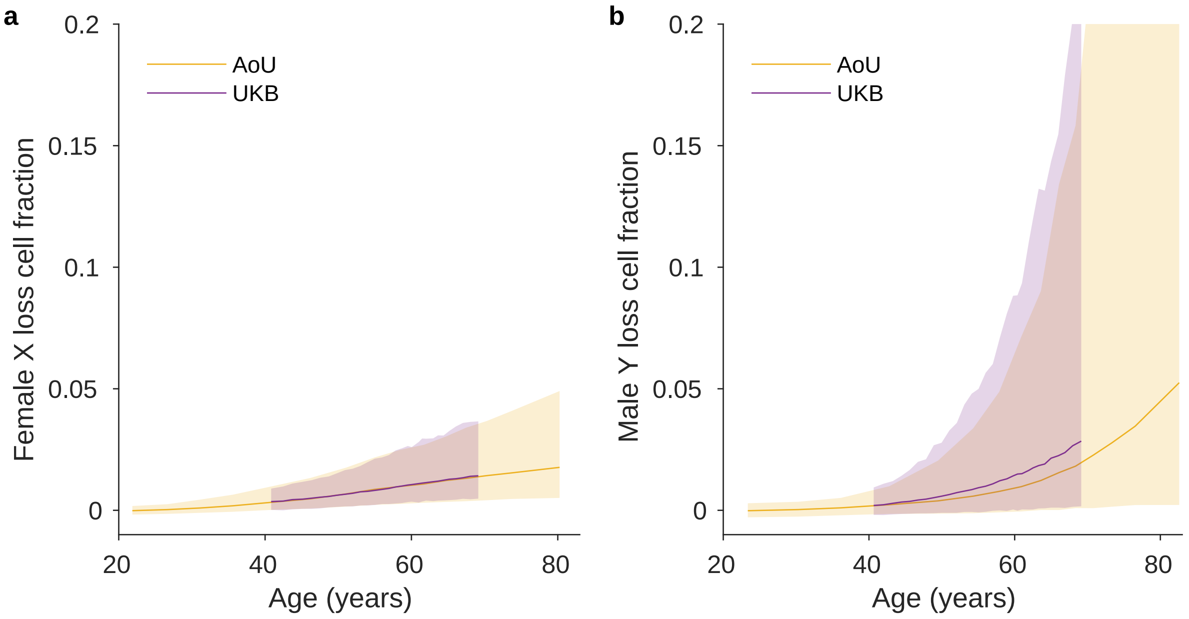

**Supplementary Figure 3. Cross-cohort consistency of estimated X loss and Y loss cell fractions. a**, X loss in female EUR-ancestry never-smokers in AoU and UKB. **b**, Y loss in male EUR-ancestry never-smokers in AoU and UKB. Participants were stratified into age tranches; curves indicate medians (shaded regions, $5^{th}$–$95^{th}$ percentiles) within each tranche. When computing percentiles of estimated cell fractions in AoU, we computed averages across the 10 individuals before and after the percentile boundary (n=20 total).

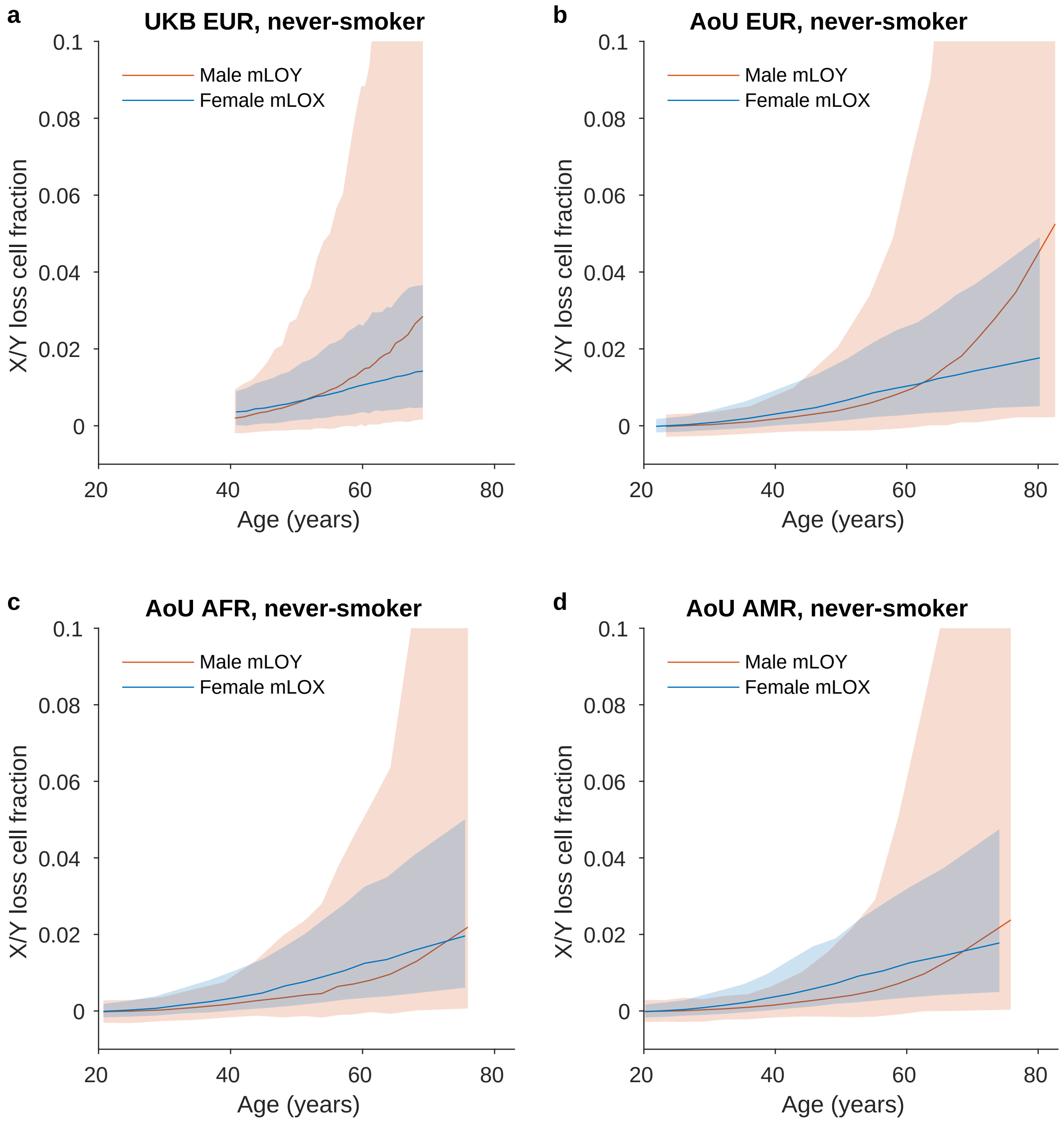

**Supplementary Figure 4. Comparison of female X loss and male Y loss cell fractions as a function of age in different sub-cohorts. a**, UKB never-smokers of EUR genetic ancestry. **b**, **c**, **d**, AoU never-smokers of EUR, AFR, and AMR genetic ancestry, respectively. Participants were stratified into age tranches; curves indicate medians (shaded regions, $5^{th}$–$95^{th}$ percentiles) within each tranche. When computing percentiles of estimated cell fractions in AoU, we computed averages across the 10 individuals before and after the percentile boundary (n=20 total).

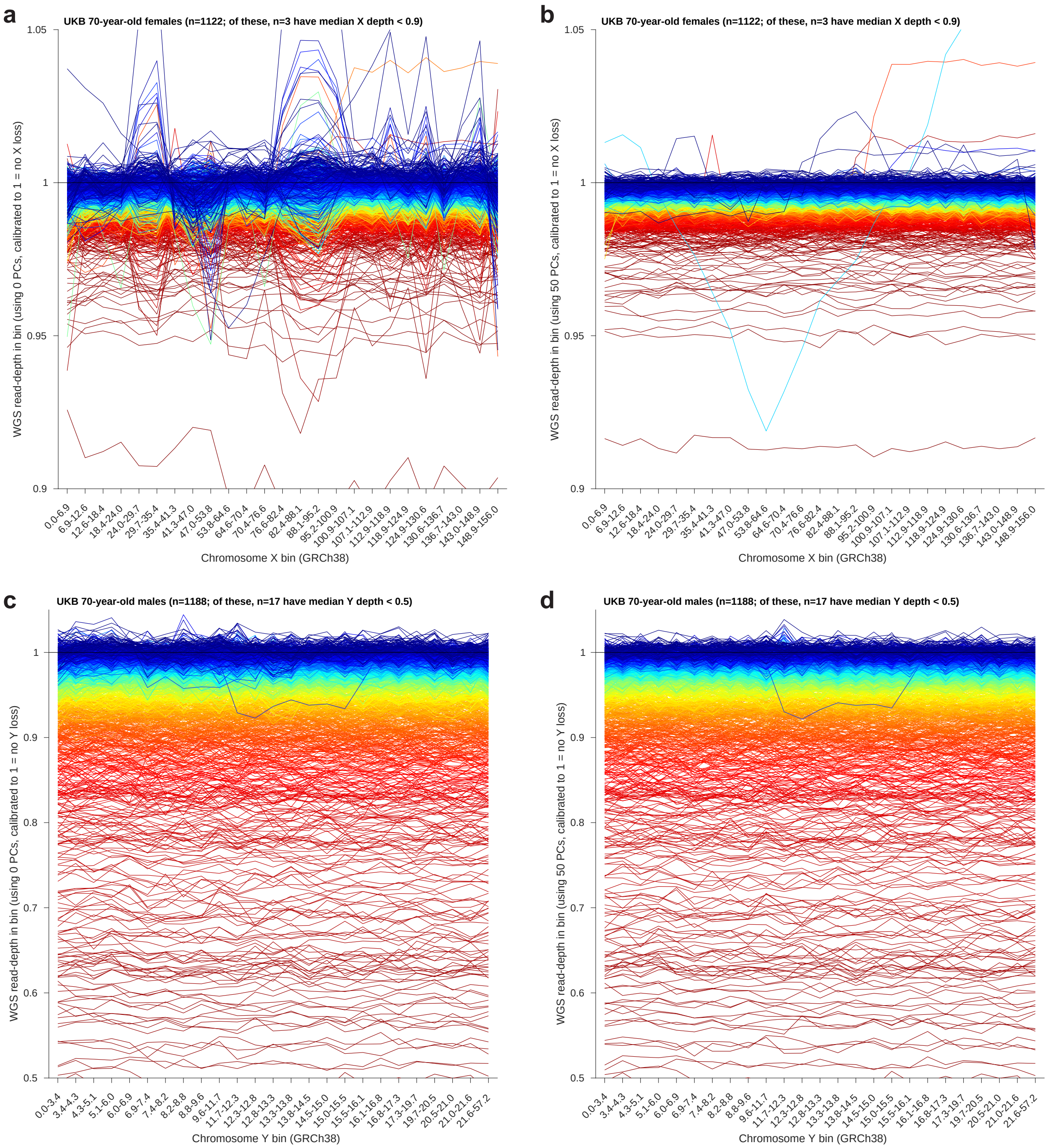

**Supplementary Figure 5. WGS read-depth profiles across chromosomes X and Y before and after denoising using 50 read-depth PCs.** Each chromosome was divided into 25 genomic bins (spanning roughly equal amounts of unmasked sequence) for read-depth computation. Each curve corresponds to a single 70-year-old UKB participant. Curves are colored by median read-depth within the chromosome. Nearly all deviations from the baseline read-depth at 1 are consistent with whole-chromosome losses. A few samples have aberrant read-depth profiles reflecting noisy sequencing, and a few have evidence of partial losses and/or gains on the X chromosome. A small fraction of samples with high levels of X or Y loss outside the plotted range are not depicted.

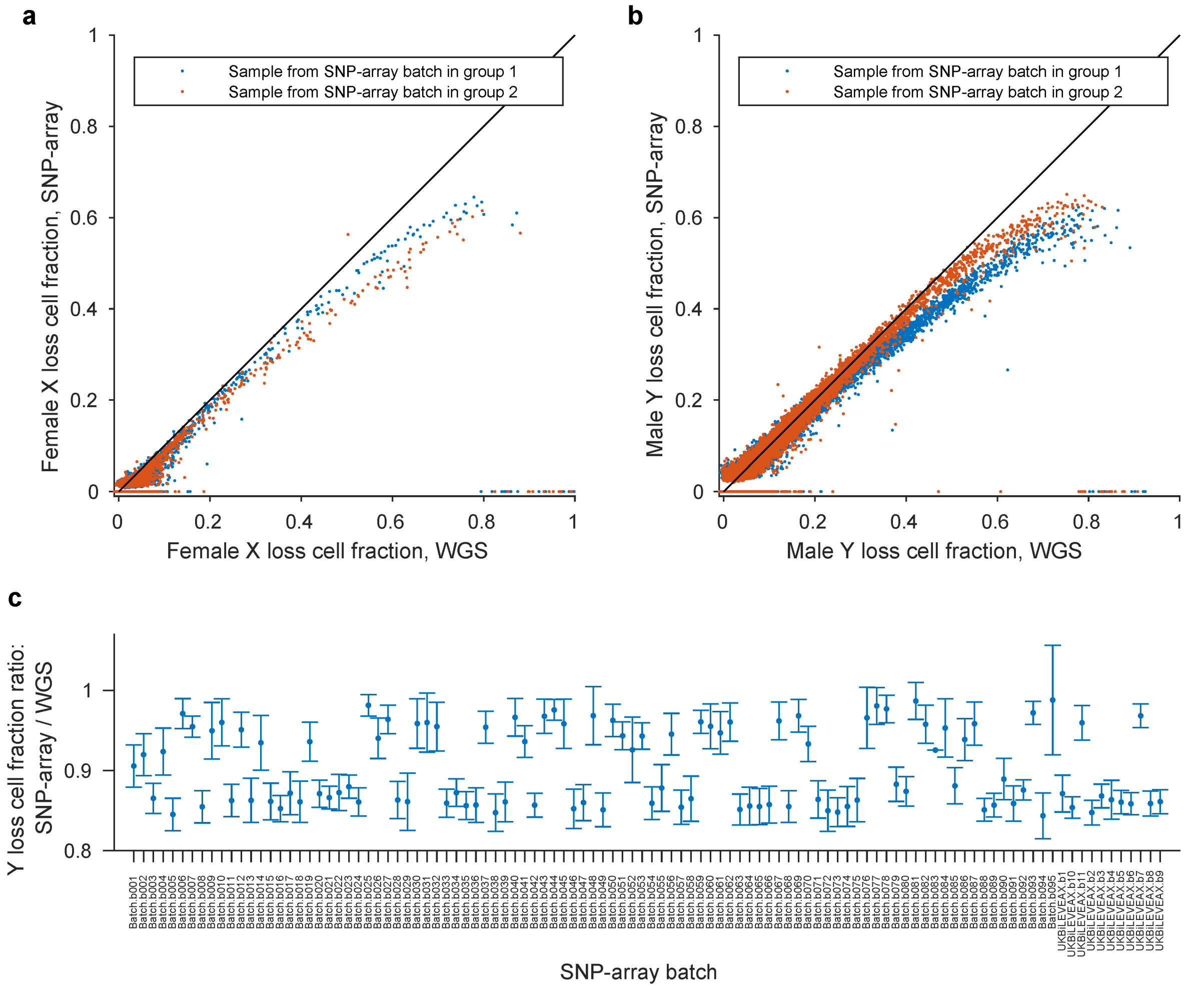

**Supplementary Figure 6. Comparison of SNP-array- and WGS-based X and Y loss detection.** **a**, Fractions of leukocytes with X loss in female UKB participants (n=251,343) estimated using SNP-array intensities[14,15] versus WGS read-depth. SNP-array-based X loss cell fractions were estimated for individuals with detectable allelic imbalance on the X chromosome[14,15] and were set to zero otherwise. A small fraction of individuals with ambiguous mCA calls on the X chromosome (not satisfying the criteria of being called as a loss with length >100Mb) were excluded. The fall-off on the upper-right of the plot and the points in the lower-right corner are consistent with X loss at very high cell fractions causing drop-out of heterozygous genotypes used in SNP-array allelic imbalance analyses, resulting in deflation of cell fraction estimates or failure to detect X loss. **b**, Analogous plot for Y loss in male UKB participants (n=197,126). A small fraction of individuals with ambiguous mCA calls on the Y chromosome (not called as a loss spanning >2Mb of PAR1) were excluded. **c**, Ratios of Y loss cell fractions estimated from SNP-array allelic imbalances versus WGS read-depth. Centers, means across individuals with SNP-array-based Y loss calls in each SNP-array batch; error bars, 95% CIs. The 106 SNP-array genotyping batches in UKB exhibit dichotomous behavior: roughly half of the batches ("group 1" in panels **a** and **b**) tended to produce higher estimates of X loss and lower estimates of Y loss, whereas the other half ("group 2") displayed the opposite bias.

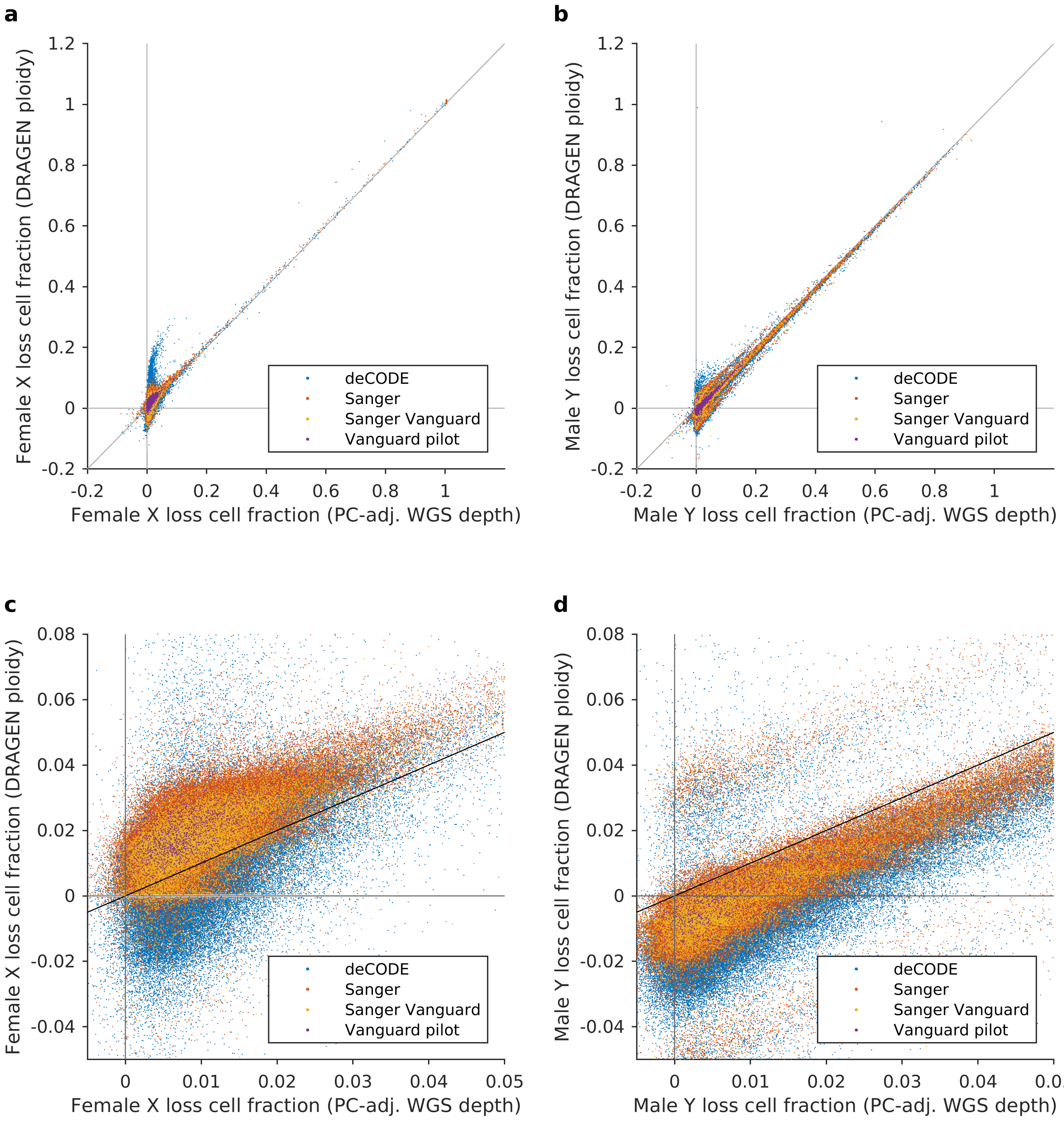

**Supplementary Figure 7. Comparison of WGS read-depth-based estimates of X and Y loss cell fractions using DRAGEN versus adjusting for 50 read-depth PCs.** **a**, Estimated fractions of X loss in female UKB participants (n=259,085). **b**, Estimated fractions of Y loss in male UKB participants (n=221,857). **c**, **d**, Zoomed-in views of panels **a** and **b** providing more detail near the origin. DRAGEN-based estimates of female X loss cell fraction were computed as 2×(1–(median X coverage)/(median autosomal coverage)). DRAGEN-based estimates of male Y loss cell fraction were computed as (1–2×(median Y coverage)/(median autosomal coverage)). The DRAGEN-based coverage estimates appear to exhibit some technical biases between and within sequencing centers.

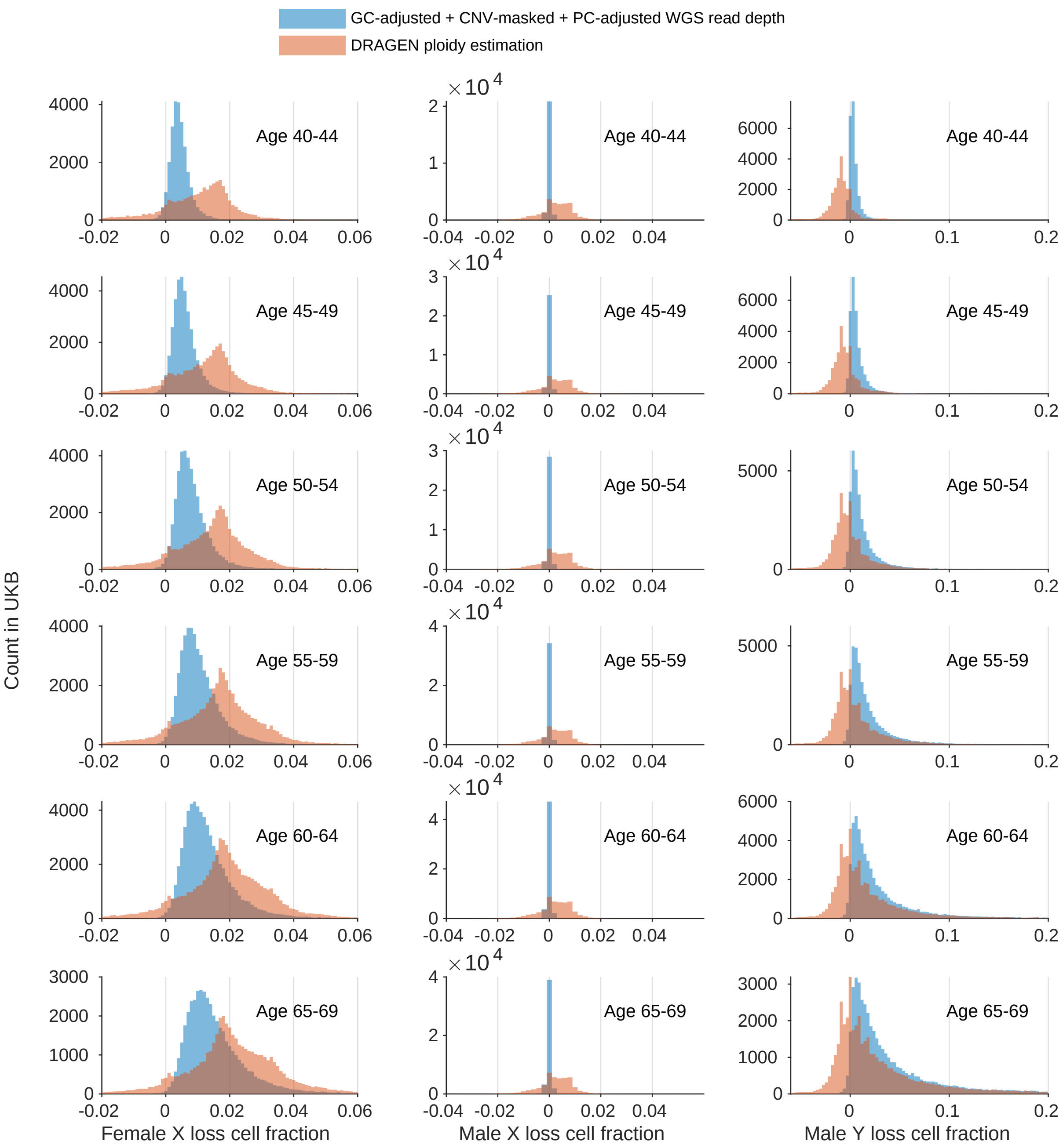

**Supplementary Figure 8. Comparison of distributions of WGS read-depth-based estimates of X and Y loss cell fractions using DRAGEN versus adjusting for 50 read-depth PCs, across age tranches in UKB.** Left, X loss in females (n=259,085); middle, X loss in males (n=221,857); right, Y loss in males (n=221,857). Negative estimates are plotted as negative values (rather than truncating them at 0) to provide an indication of the amount of measurement noise. Estimates of X loss in males (which appears not to be tolerated in peripheral leukocytes[138]) were included to further evaluate amounts of measurement noise. DRAGEN-based estimates of male X loss cell fraction were computed as (1–2×(median X coverage)/(median autosomal coverage)). The multimodal distributions from DRAGEN indicate batch effects.

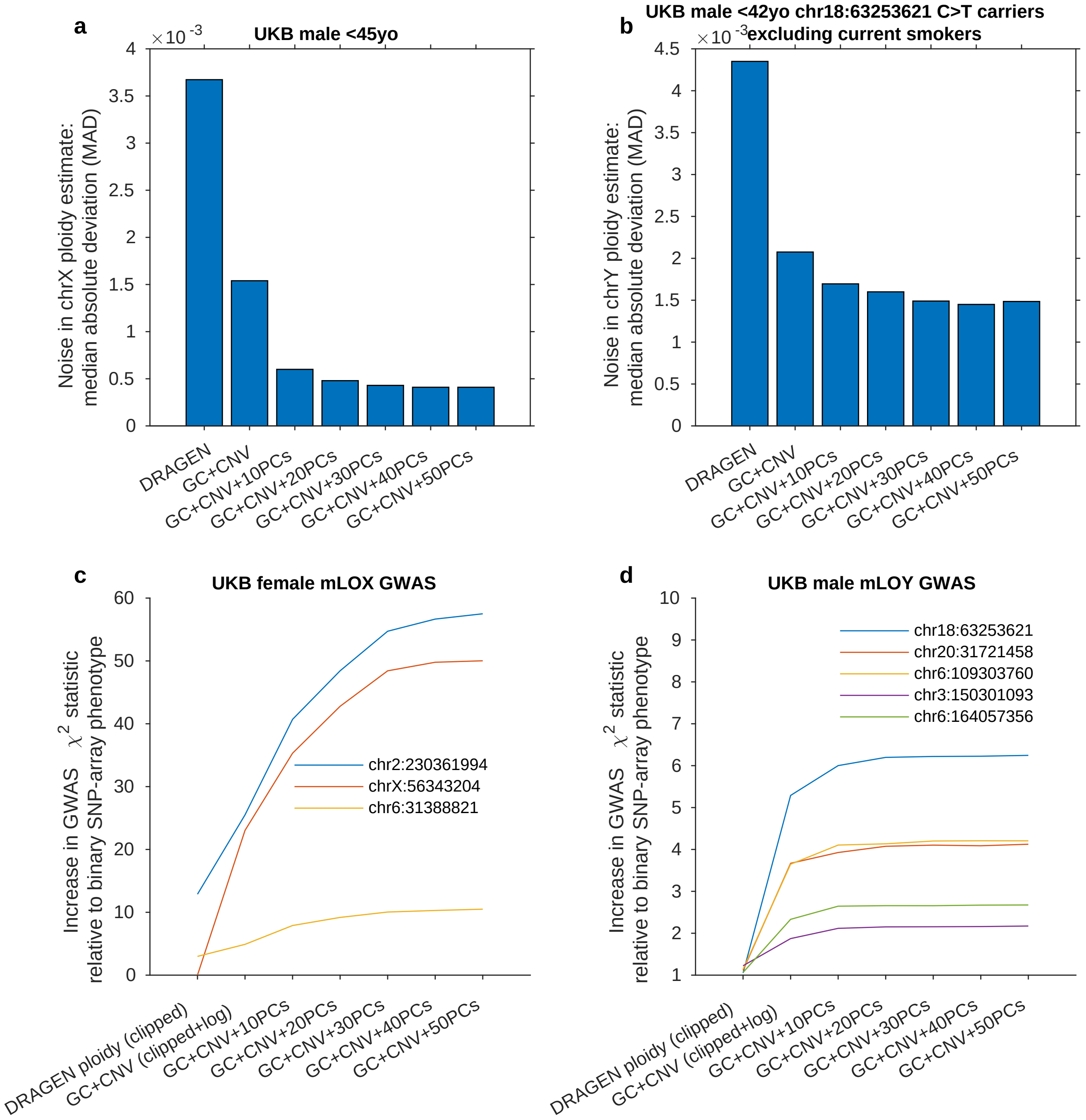

**Supplementary Figure 9. Comparison of measurement error in sex chromosome ploidy estimates from WGS read-depth using DRAGEN versus PC-denoising, and comparison of association strength at top GWAS hits.** (legend continued on next page)

**Supplementary Figure 9** ***(previous page).*** **a**, Median absolute deviation (MAD) of X chromosome ploidy in male UKB participants aged<45 years (n=23,181) estimated using DRAGEN coverage metrics or by adjusting for varying numbers of read-depth PCs (after GC-correction and CNV-masking; Methods). Males are expected to have negligible amounts of X loss[138]. **b**, MAD of Y chromosome ploidy in male UKB non-smokers aged<42 years carrying the *BCL2* SNP associated with reduced Y loss (n=312). Because Y loss is already noticeable at age 40 (the minimum age in UKB), this subset of individuals was chosen to minimize the expected amount of Y loss, facilitating assessment of measurement noise. Even in this "control" cohort, the MAD is still expected to slightly overestimate the amount of measurement noise owing to the presence of small amounts of Y loss. **c**, Association strengths at top GWAS hits for DRAGEN and PC-denoised female X loss phenotypes relative to SNP-array-based binary X loss calls (likely LOX at FDR 10%; ref.[14,15]). DRAGEN estimates of X loss cell fraction were clipped to 0–0.04 as in analyses of PC-adjusted read-depth. PC-adjusted read-depth estimates were further log-transformed for GWAS (Methods). Association analyses were performed using linear regression with age, age$^2$, smoking status, and sequencing center ("WGS provider") as covariates, restricting to female EUR-ancestry individuals (n=230,128). **d**, Analogous comparison of association strengths with male Y loss phenotypes derived from WGS read-depth (using DRAGEN or PC-denoising) versus SNP-array intensities (binary calls for likely LOY based on allelic imbalance in PAR1[11]). DRAGEN estimates of Y loss cell fraction were clipped to 0–0.2 as in analyses of PC-adjusted read-depth. Association analyses were performed on UKB EUR-ancestry males (n=199,808) using the same covariates as above.

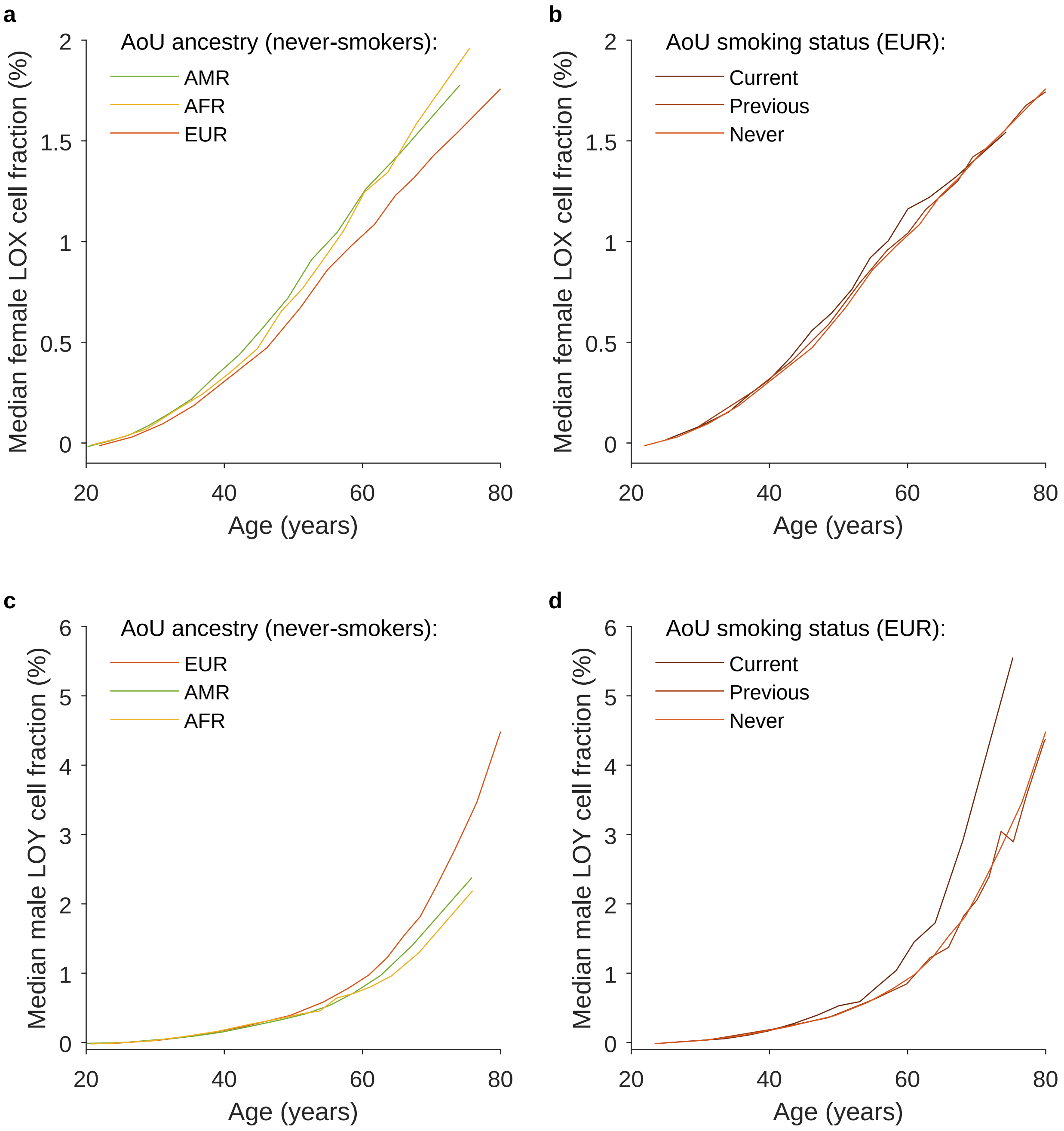

**Supplementary Figure 10. Variation of female X loss and male Y loss cell fractions with genetic ancestry and smoking status.** Panels **a** and **b** stratify AoU participants by ancestry, controlling for smoking status by restricting to never-smokers (n=130,100 and 66,600). Panels **c** and **d** stratify AoU participants by smoking status, controlling for ancestry by restricting to EUR-ancestry participants (n=113,200 and 78,800).

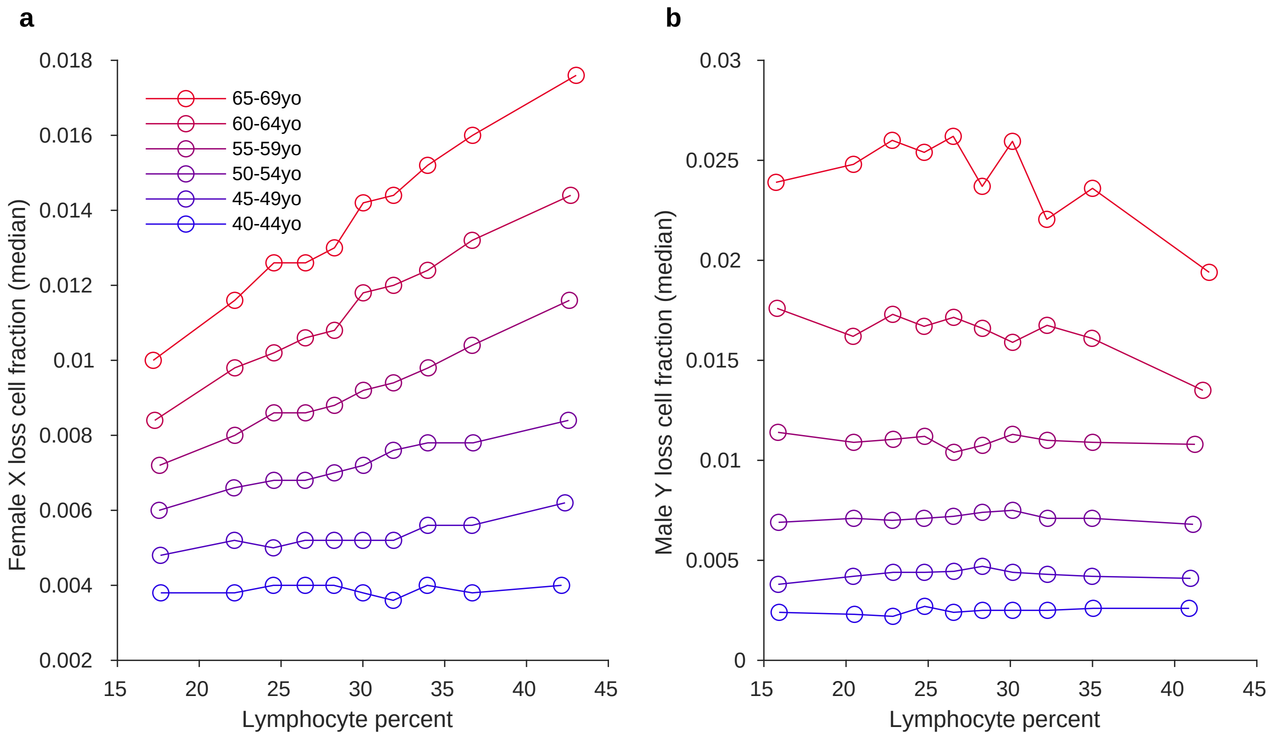

**Supplementary Figure 11. X loss cell fractions increase with lymphocyte percentages in an age-dependent manner. a**, Median X loss cell fraction in deciles of lymphocyte percentage within 5-year age strata of female UKB participants. **b**, Analogous plot for Y loss in males, showing a much weaker correlation with lymphocyte percentages.

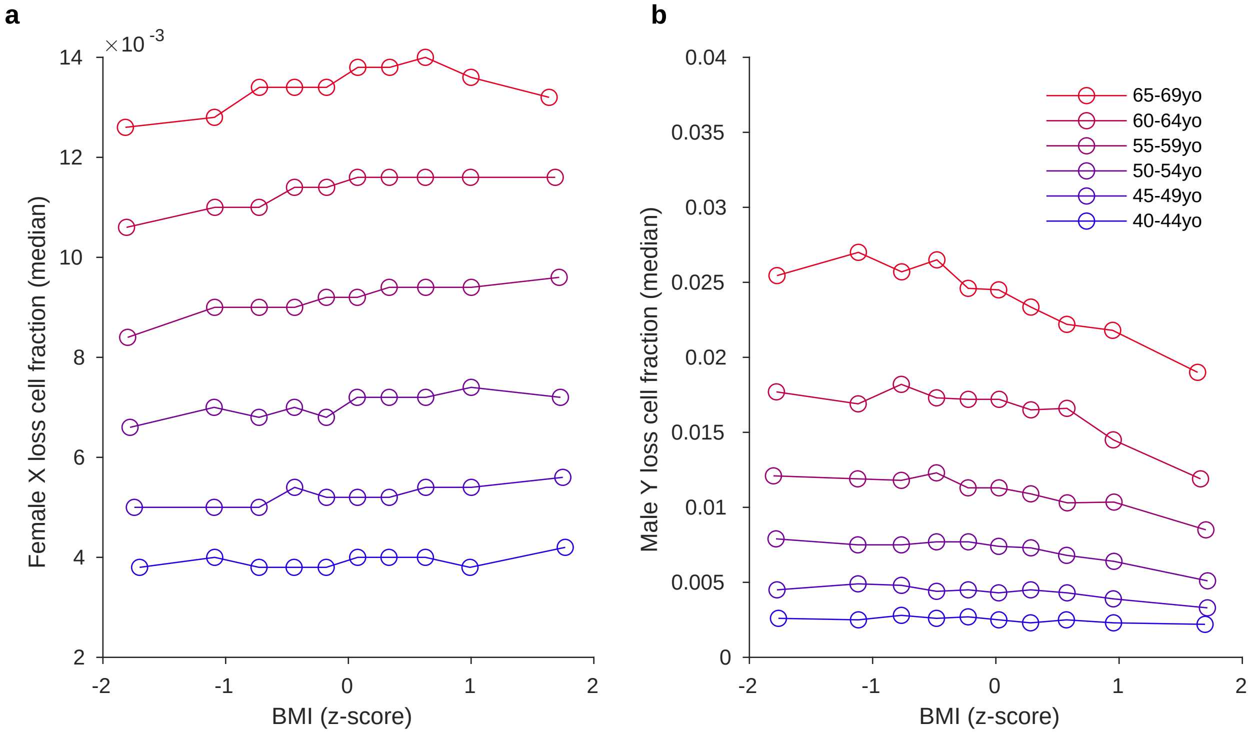

**Supplementary Figure 12. Y loss cell fractions decrease with increasing BMI in an age-dependent manner. a**, Median X loss cell fraction in deciles of BMI within 5-year age strata of female UKB participants. **b**, Analogous plot for Y loss in males, showing a strong age dependence of the BMI association, as previously observed[12].

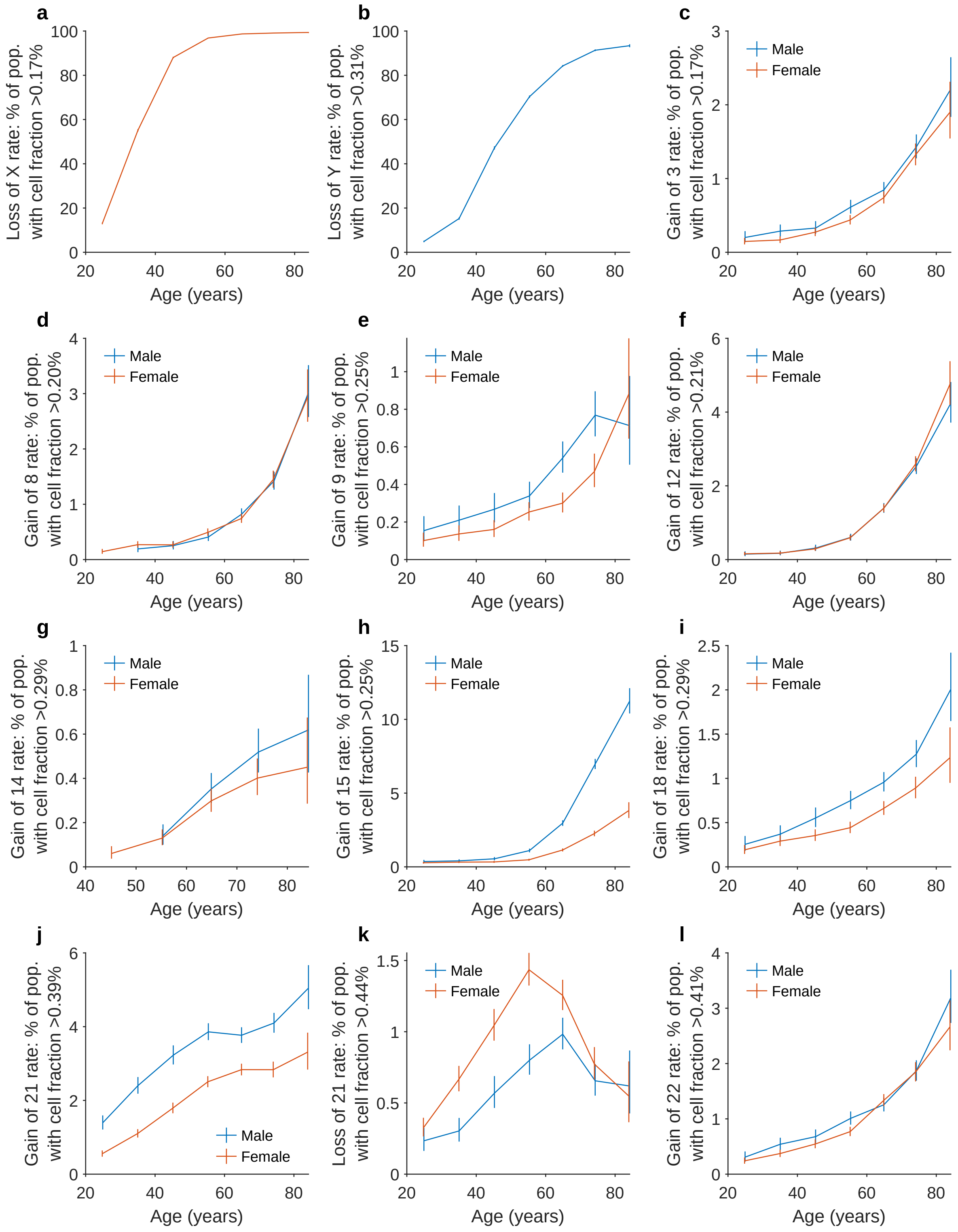

**Supplementary Figure 13. Rates of detectable aneuploidy in AoU participants (n=348,956) stratified by sex.** Positivity for each aneuploidy was defined using the threshold for minimum estimated cell fraction indicated in the y-axis to control FDR at ∼0.05 (Methods). The unusual shape of the curves for gain of 21 (panel **j**) is attributable to a larger fraction of older AoU participants being EUR-ancestry, in which chromosome 21 gain is rarer. Error bars, 95% CIs.

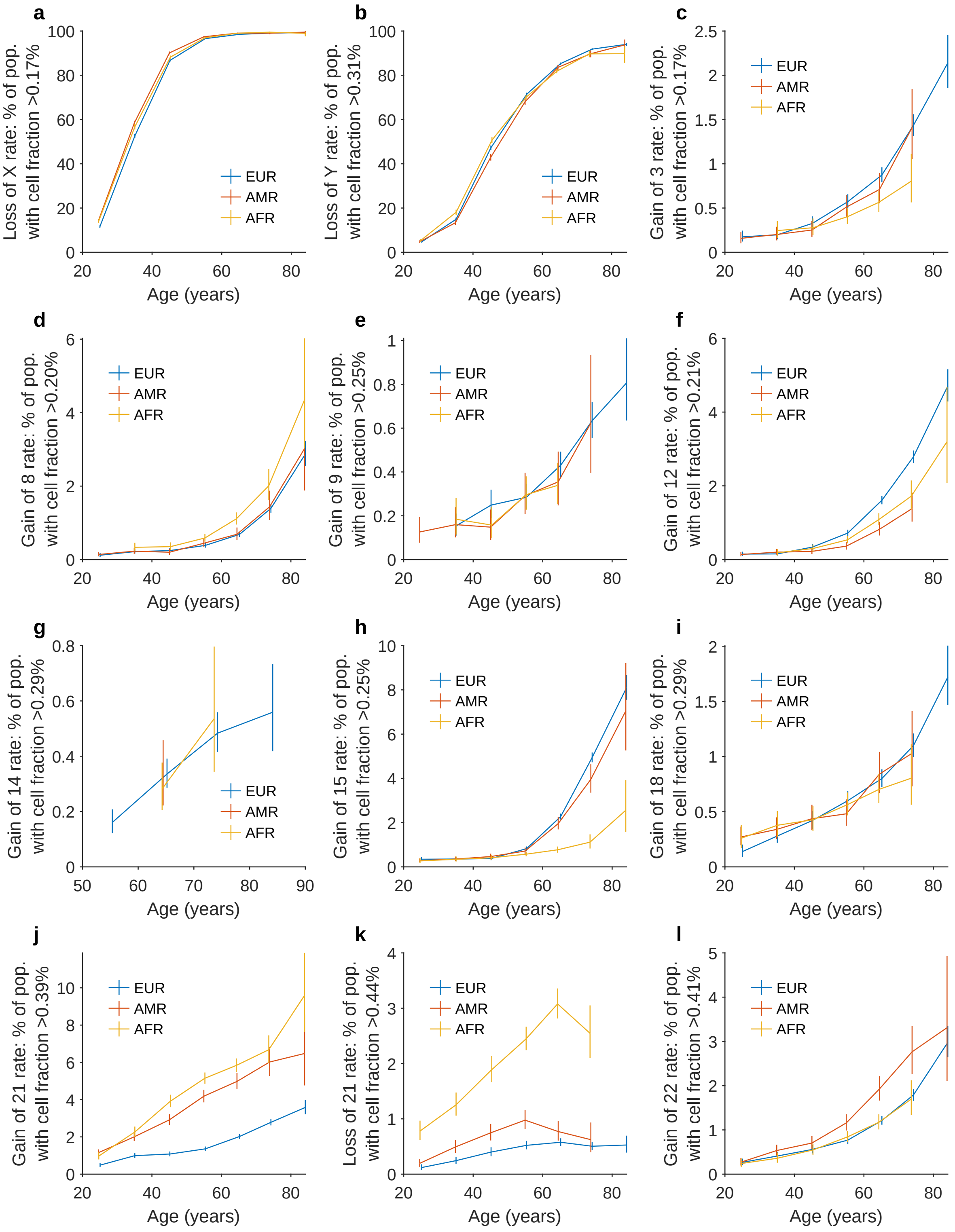

**Supplementary Figure 14. Rates of detectable aneuploidy in AoU participants (n=348,956) stratified by genetic ancestry.** Positivity for each aneuploidy was defined using the threshold for minimum estimated cell fraction indicated in the y-axis to control FDR at ∼0.05 (Methods). Error bars, 95% CIs.

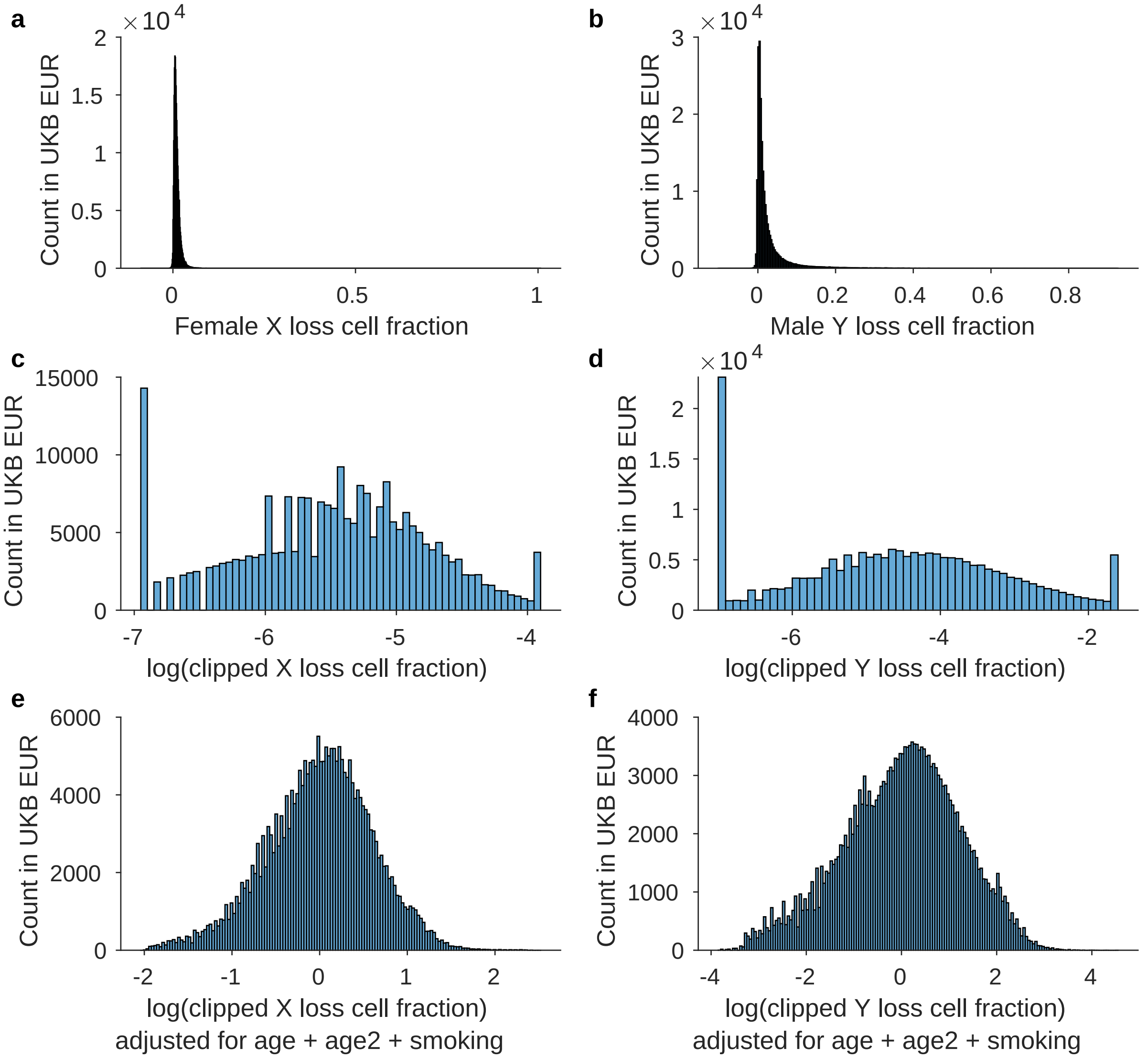

**Supplementary Figure 15. Distributions of X and Y loss GWAS phenotypes. a**, **b**, Distributions of estimated fractions of white blood cells harboring X loss (**a**, female EUR-ancestry UKB participants; n=243,385) and Y loss (**b**, male EUR-ancestry UKB participants; n=208,000). **c**, **d**, Distributions of natural log-transformed, clipped X and Y loss cell fractions (Methods). **e**, **f**, Distributions of residual X and Y loss quantitative phenotypes after adjusting for age, age$^2$, and smoking covariates. These residual phenotypes are close to Gaussian, making them suitable for regression analyses in GWAS.

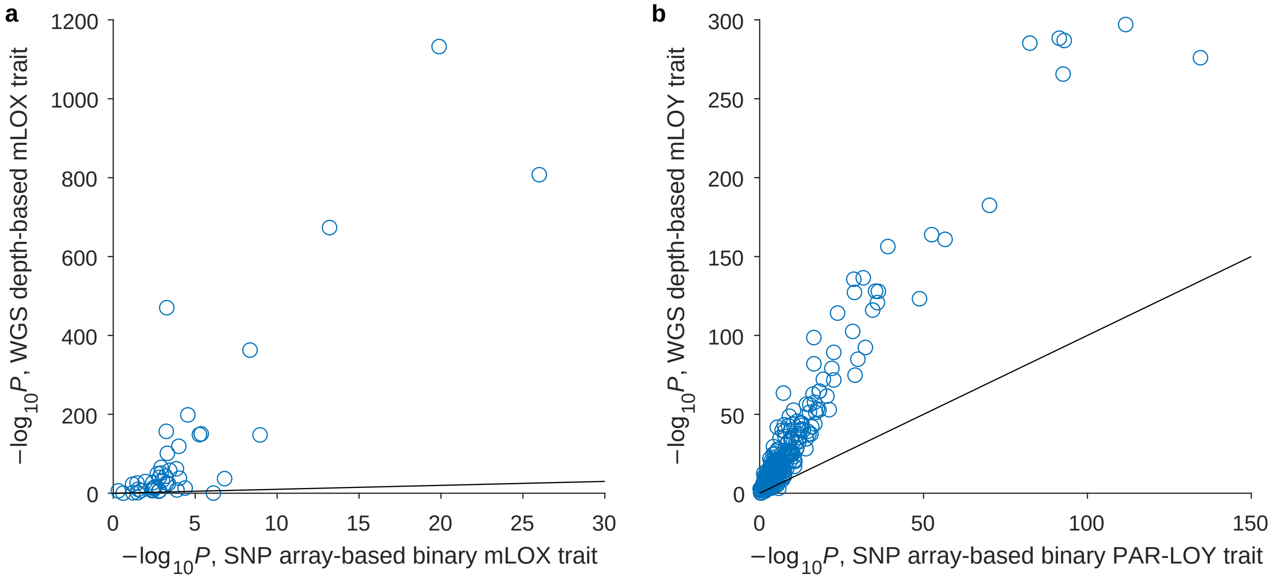

**Supplementary Figure 16. Comparison of GWAS association strengths for WGS read-depth-based X and Y loss quantitative phenotypes versus SNP-array-based binary phenotypes.** **a**, Association strengths ($-\log_{10}$ *P*-values) from GWAS of the WGS-based X loss quantitative phenotype (y-axis; Methods) compared to SNP-array-based binary X loss calls (likely LOX at FDR 10%; ref.[14,15]; x-axis). *P*-values are from linear mixed model analyses using BOLT-LMM[82] on female UKB EUR-ancestry participants. The SNP-array analyses (n=246,669) included age, age$^2$, 20 genetic principal components, genotyping array, and assessment center as covariates; the WGS analyses (n=242,531) included age, age$^2$, smoking, and 20 PCs as covariates. Data are plotted for 44 lead variants from the GWAS meta-analysis of X loss of ref.[13] that were tested in both GWAS. **b**, Association strengths ($-\log_{10}$ *P*-values) from GWAS of the WGS-based Y loss quantitative phenotype (y-axis; Methods) compared to the previous SNP-array-based binary PAR-LOY phenotype[11] (x-axis). *P*-values are from BOLT-LMM on male UKB EUR-ancestry participants (n=207,487 for SNP-array; n=207,363 for WGS) using the same covariates as above. Data are plotted for 263 lead variants from the GWAS of Y loss in MVP EUR[12] that were tested in both GWAS.

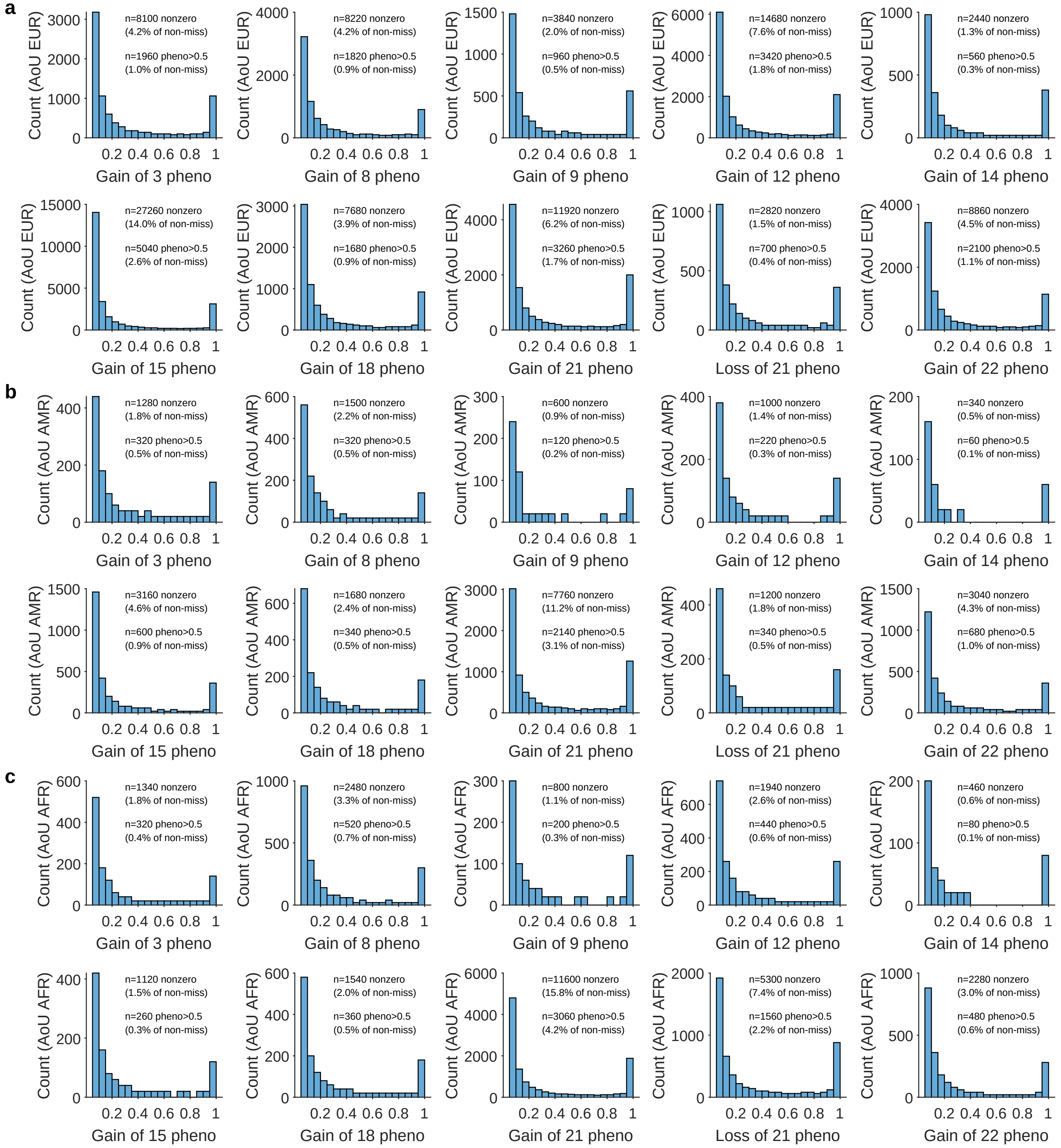

**Supplementary Figure 17. Distributions of probabilistic autosomal aneuploidy GWAS phenotypes. a**, EUR; **b**, AMR; **c**, AFR participants in AoU. Chromosomal read-depth deviations were converted into GWAS phenotypes on a 0–1 scale using priors that accounted for age and sex (Methods). For each aneuploidy, most participants did not have evidence of the aneuploidy and were assigned a phenotype value of 0; histograms show distributions of nonzero phenotype values, with the fraction of nonzeros indicated above. Participant counts were rounded to the nearest 20.

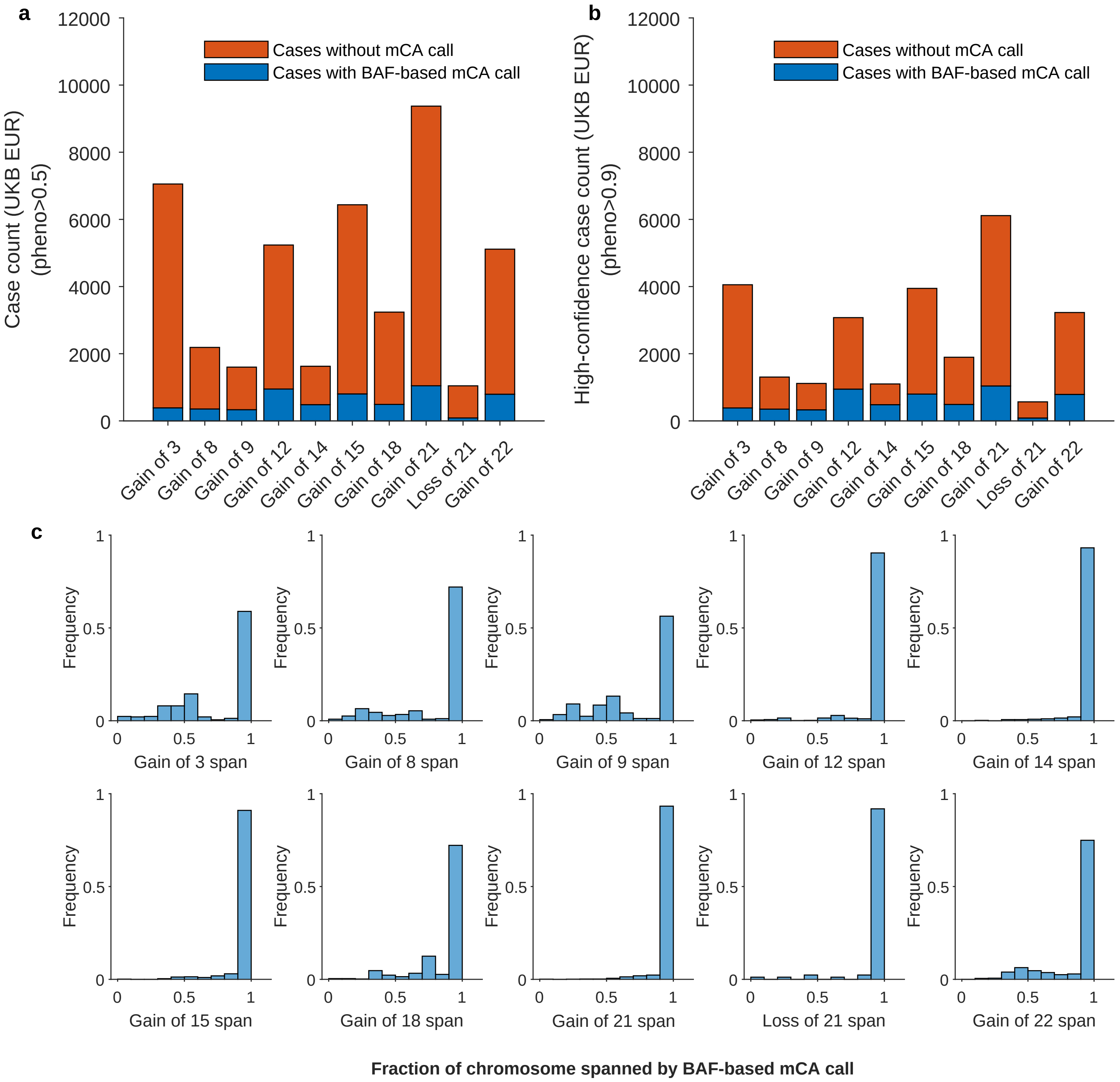

**Supplementary Figure 18. Comparison of WGS read-depth-based autosomal aneuploidy calls to allelic imbalance-based mCA calls. a–b**, Numbers of individuals with probabilistic autosomal aneuploidy calls (**a**, phenotype value >0.5; **b**, phenotype value >0.9) with and without a corresponding mCA call (i.e., same chromosome, same copy number change) from allelic imbalance (phased BAF) analysis[17]. Data are shown for UKB EUR-ancestry participants (n=451,924 after excluding the 0.5% with noisiest WGS read-depth profiles). **c**, Distributions of the proportion of each chromosome spanned by corresponding mCA calls. Each plot shows data from individuals represented in the blue bars in panel **a**. For acrocentric chromosomes, the plot shows the fraction of the q-arm spanned.

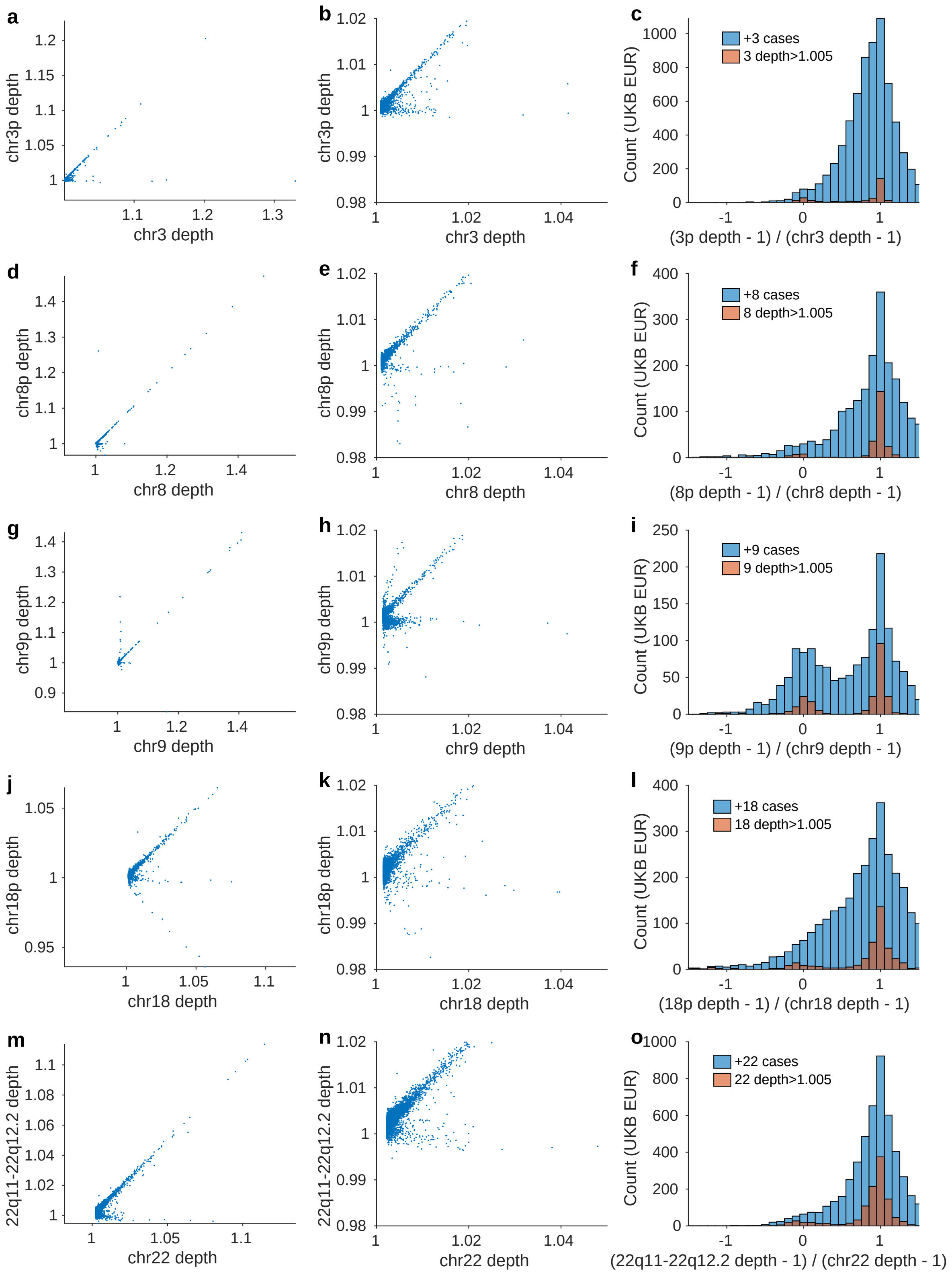

**Supplementary Figure 19. Evaluation of the extent to which increased WGS read-depth on some autosomes may reflect arm-level rather than whole-chromosome mosaic gains.** (legend continued on next page)

**Supplementary Figure 19** ***(previous page)*****.** Among nine autosomes for which mosaic whole-chromosome gains are commonly observed in leukocytes (chromosomes 3, 8, 9, 12, 14, 15, 18, 21, and 22)[15–17], five of the nine autosomes also commonly experience sub-chromosomal mosaic gains that comprise a moderate fraction of all mosaic gains (chromosomes 3, 8, 9, 18, and 22)[17]. For these five autosomes, we evaluated the extent to which increased WGS-read depth (specifically, the median across the chromosome, which we used to identify mosaic gains; Methods) tended to capture full-chromosome versus sub-chromosomal gains. We did so by comparing chromosome-wide median read-depth (x-axis of scatter plots) to median read-depth across the p-arms of chromosomes 3, 8, 9, and 18 and across the centromeric half of 22q (before chr22:32000000) (y-axis of scatter plots). The plots in the middle column are zoom-ins of the plots in the left column. Each data point corresponds to a UKB EUR-ancestry participant with a probabilistic call of a mosaic gain (phenotype value $>0.5$) based on elevated chromosome-wide median read-depth, taking into account age and sex (Methods). The right column of plots shows distributions of the ratio of the read-depth deviation in the sub-chromosomal region compared to the read-depth deviation of the chromosome-wide median. Count histograms restricted to the subset of individuals with higher read-depths ($>1.005$, for which the ratio is less noisy) are also shown in orange-brown. These data indicate that for all five autosomes, the majority of mosaic gains are full-chromosome gains, and chromosome 9 has the largest fraction of sub-chromosomal gains (roughly half of all mosaic gains), consistent with previous analyses[17].

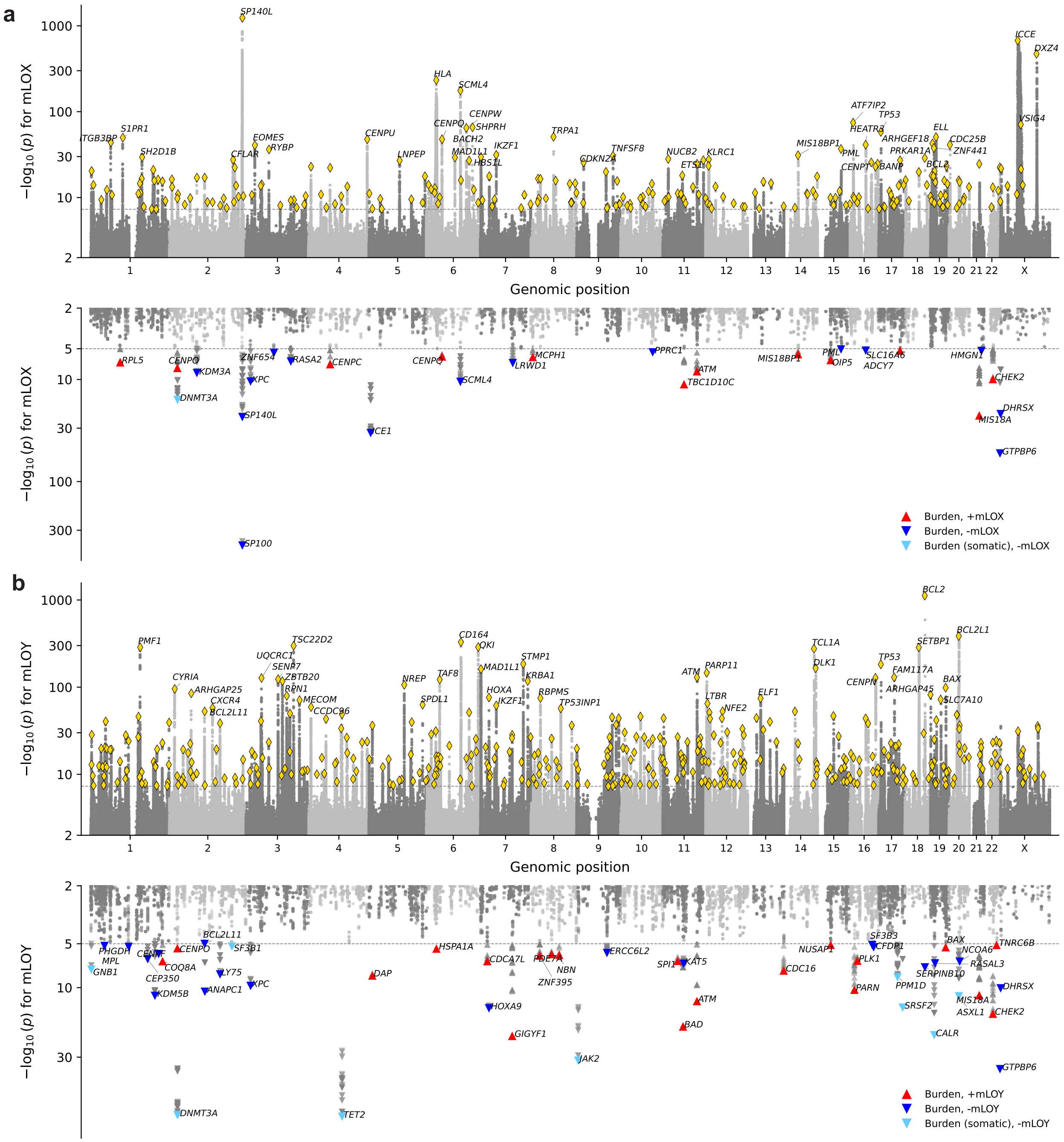

**Supplementary Figure 20. Common-variant and gene-based rare coding burden associations with X and Y loss.** GWAS and burden test results for WGS-based quantitative phenotypes for (**a**) X loss and (**b**) Y loss in UKB EUR (n=242,531 and n=207,363, respectively). Top: common variant associations. Yellow diamonds mark lead variants at independent genome-wide significant loci ($P<5\times10^{-8}$). Nearby genes are annotated for the top 40 lead variants. Bottom: gene-level burden test associations. For each gene, rare coding variants were collapsed using up to 36 burden masks (Methods); each triangle represents a single mask. Red, upward triangles indicate genes for which a burden of germline coding variants associated with increased X/Y loss ($P<1\times10^{-5}$, FDR$\sim$0.01); dark blue, downward triangles indicate associations of germline coding variation with decreased X/Y loss; light blue, downward triangles indicate associations with decreased X/Y loss driven by somatic coding variation.

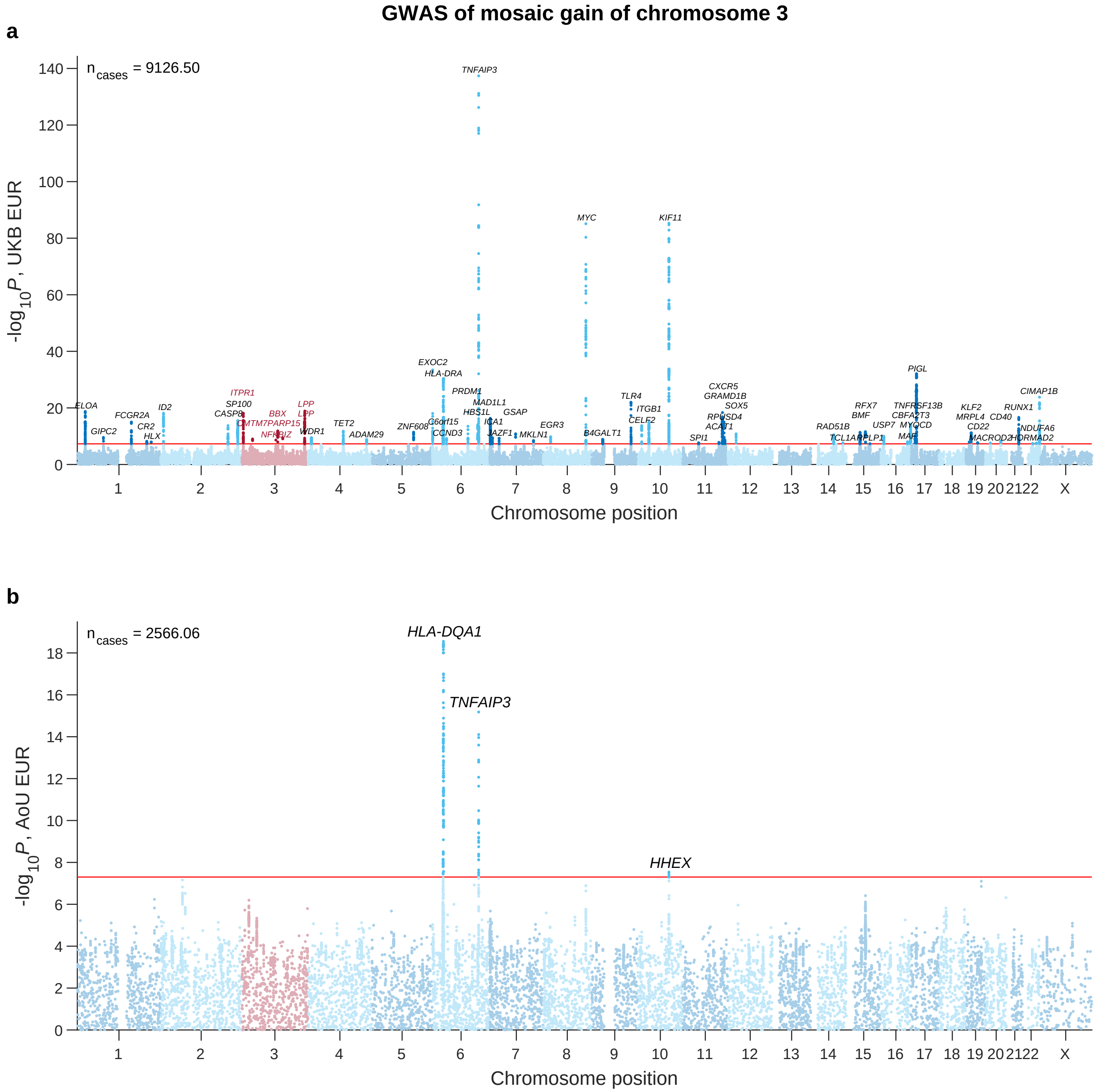

**Supplementary Figure 21. Associations of common inherited variants with mosaic gain of chromosome 3.** *P*-values are from linear regression on probabilistic calls of mosaic aneuploidies ($n_{cases}$ = cohort-wide sum) in unrelated individuals, with sex, age, age$^2$, smoking, and genetic principal components included as covariates (Methods). **a**, UKB EUR (n=406,312); **b**, AoU EUR (n=184,423). Lead variants ($P<5\times10^{-8}$) are labeled with the nearest gene.

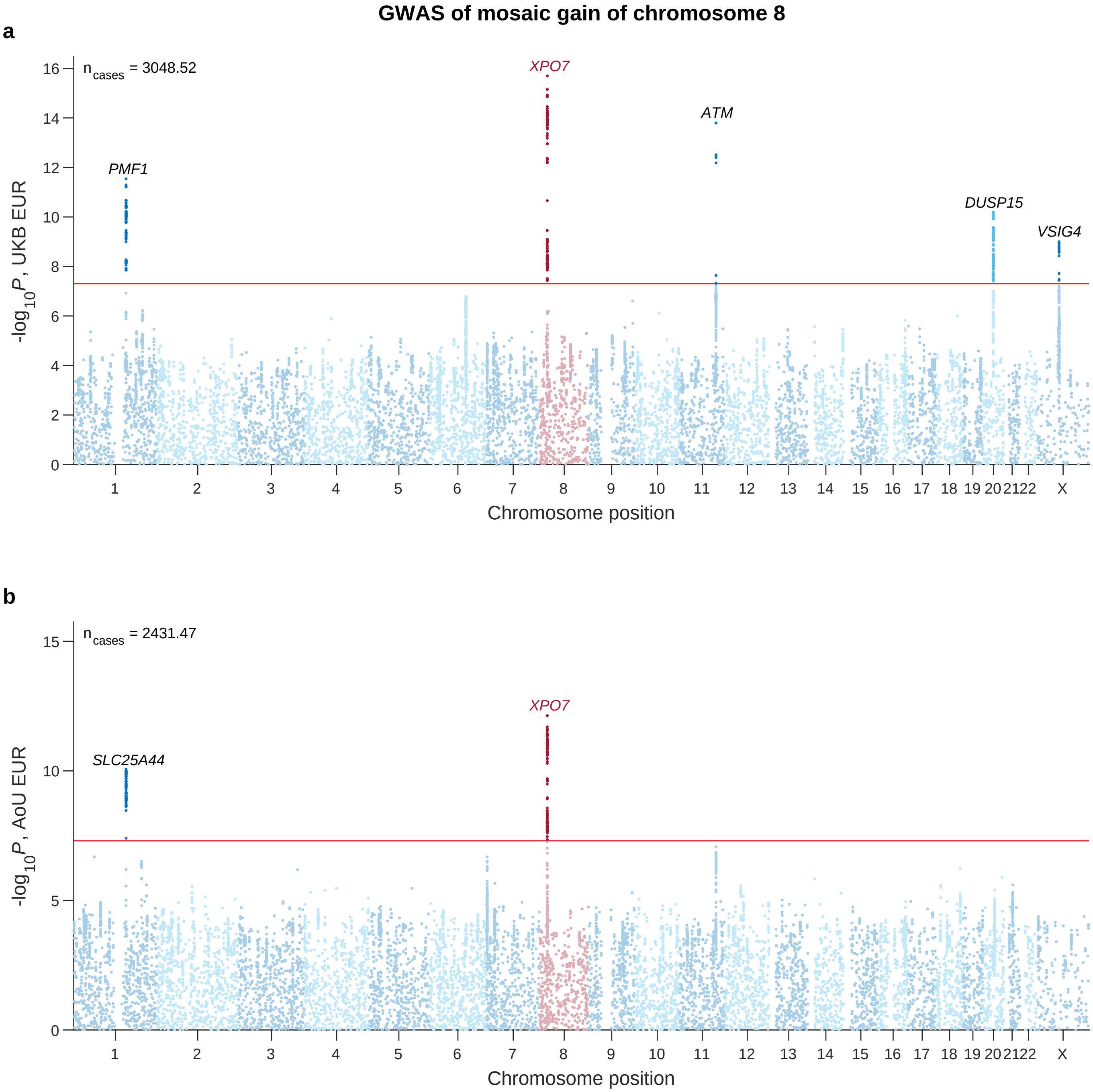

**Supplementary Figure 22. Associations of common inherited variants with mosaic gain of chromosome 8.** *P*-values are from linear regression on probabilistic calls of mosaic aneuploidies ($n_{cases}$ = cohort-wide sum) in unrelated individuals, with sex, age, age$^2$, smoking, and genetic principal components included as covariates (Methods). **a**, UKB EUR (n=406,070); **b**, AoU EUR (n=185,716). Lead variants ($P<5\times10^{-8}$) are labeled with the nearest gene.

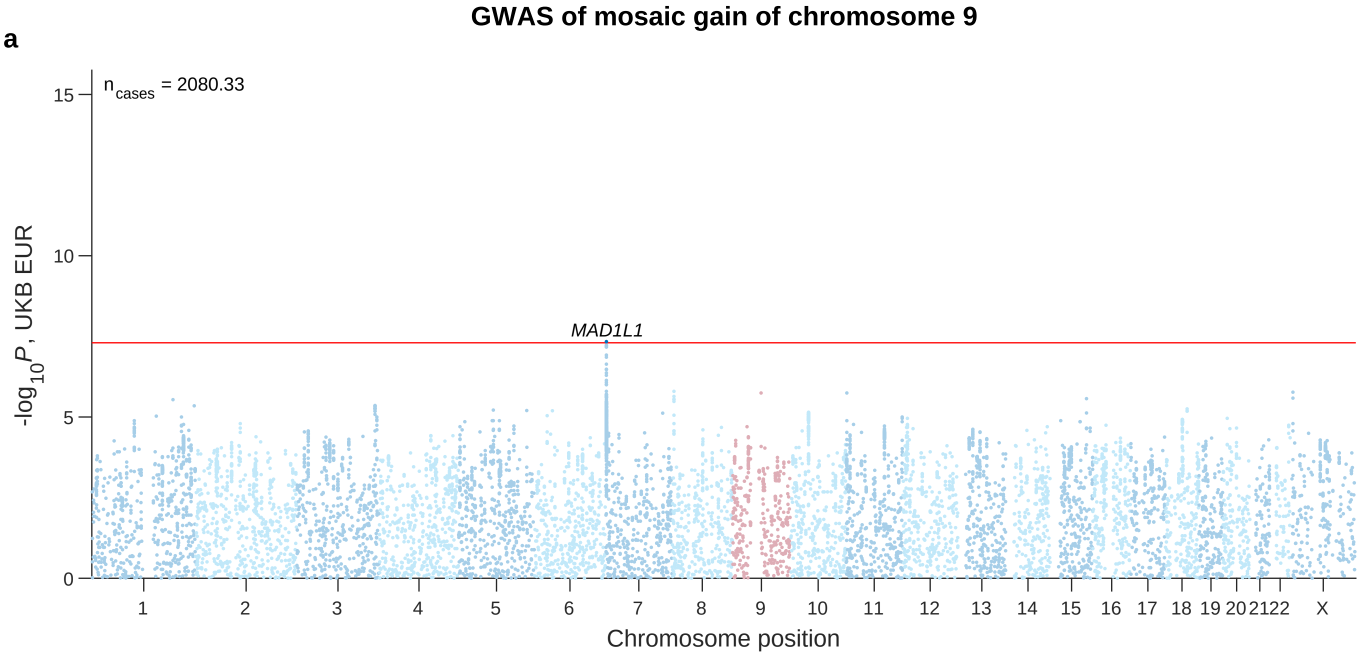

**Supplementary Figure 23. Associations of common inherited variants with mosaic gain of chromosome 9.** *P*-values are from linear regression on probabilistic calls of mosaic aneuploidies ($n_{cases}$ = cohort-wide sum) in unrelated individuals, with sex, age, $age^2$, smoking, and genetic principal components included as covariates (Methods). **a**, UKB EUR (n=405,728); **b**, AoU EUR (n=185,176). Lead variants ($P<5\times10^{-8}$) are labeled with the nearest gene.

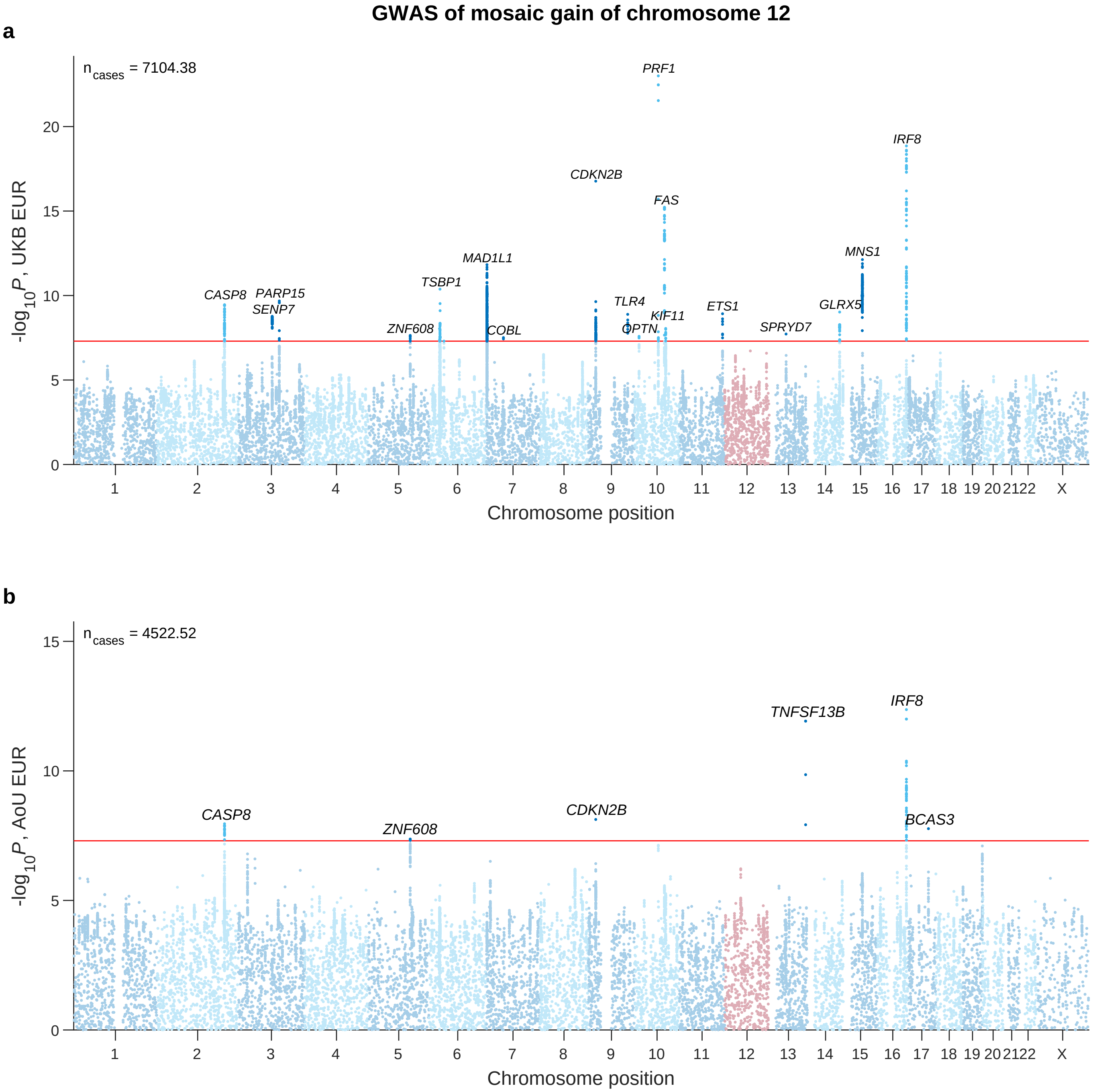

**Supplementary Figure 24. Associations of common inherited variants with mosaic gain of chromosome 12.** *P*-values are from linear regression on probabilistic calls of mosaic aneuploidies ($n_{cases}$ = cohort-wide sum) in unrelated individuals, with sex, age, age$^2$, smoking, and genetic principal components included as covariates (Methods). **a**, UKB EUR (n=407,298); **b**, AoU EUR (n=185,239). Lead variants ($P<5\times10^{-8}$) are labeled with the nearest gene.

GWAS of mosaic gain of chromosome 14

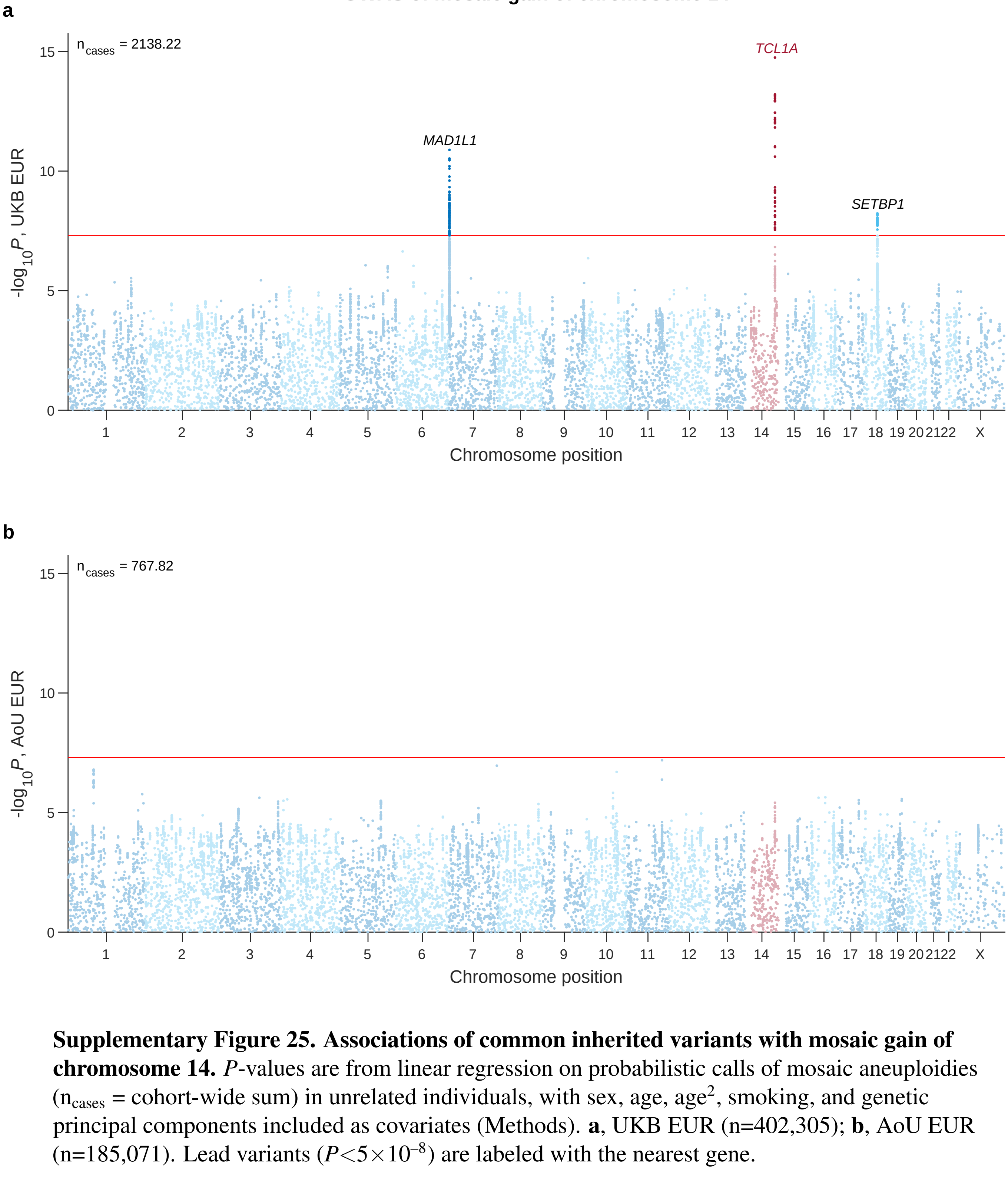

**Supplementary Figure 25. Associations of common inherited variants with mosaic gain of chromosome 14.** *P*-values are from linear regression on probabilistic calls of mosaic aneuploidies ($n_{cases}$ = cohort-wide sum) in unrelated individuals, with sex, age, age$^2$, smoking, and genetic principal components included as covariates (Methods). **a**, UKB EUR (n=402,305); **b**, AoU EUR (n=185,071). Lead variants ($P<5\times10^{-8}$) are labeled with the nearest gene.

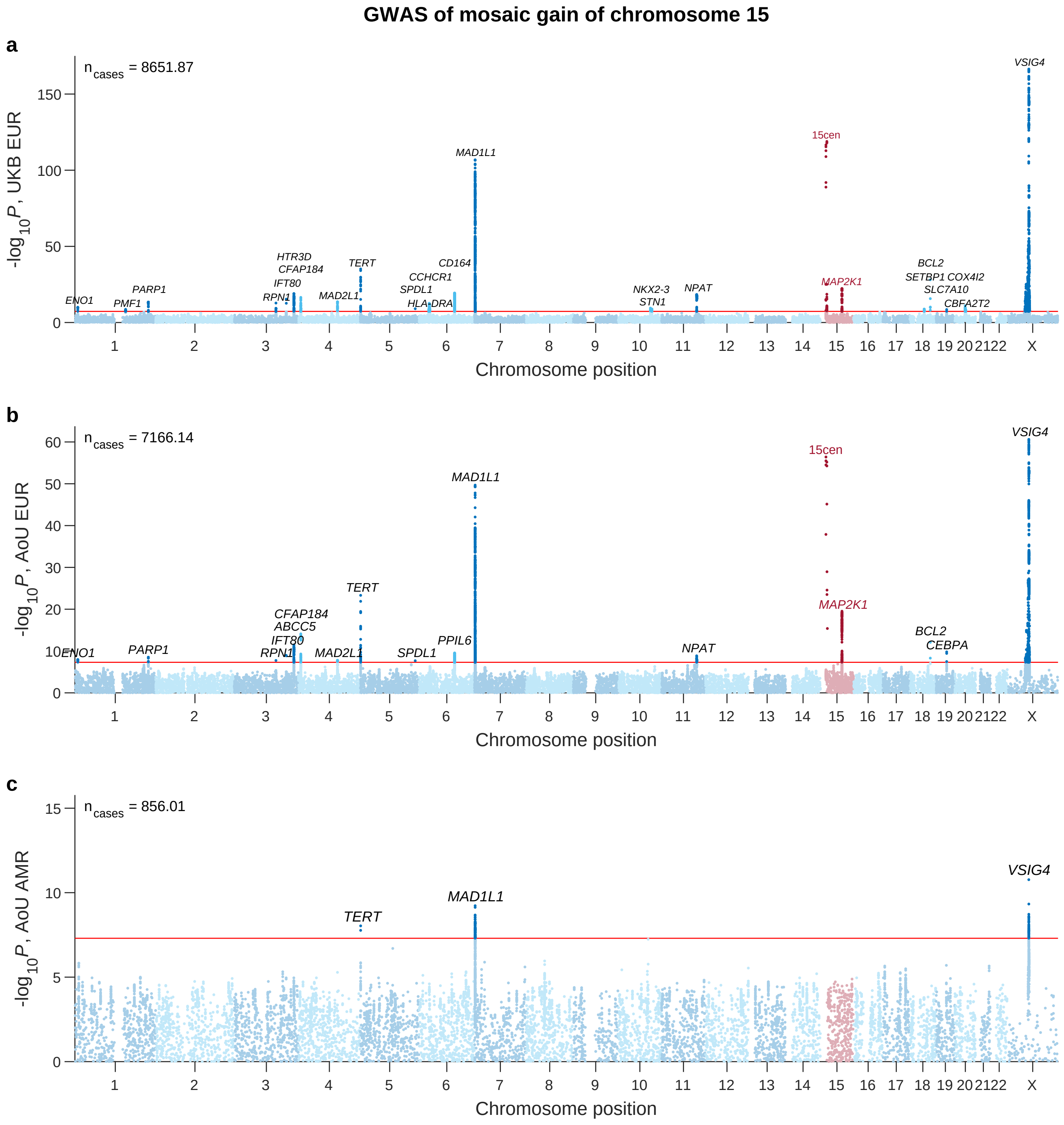

**Supplementary Figure 26. Associations of common inherited variants with mosaic gain of chromosome 15.** $P$-values are from linear regression on probabilistic calls of mosaic aneuploidies ($n_{cases}$ = cohort-wide sum) in unrelated individuals, with sex, age, age$^2$, smoking, and genetic principal components included as covariates (Methods). **a**, UKB EUR (n=405,348); **b**, AoU EUR (n=186,117); **c**, AoU AMR (n=62,163). Lead variants ($P<5\times10^{-8}$) are labeled with the nearest gene.

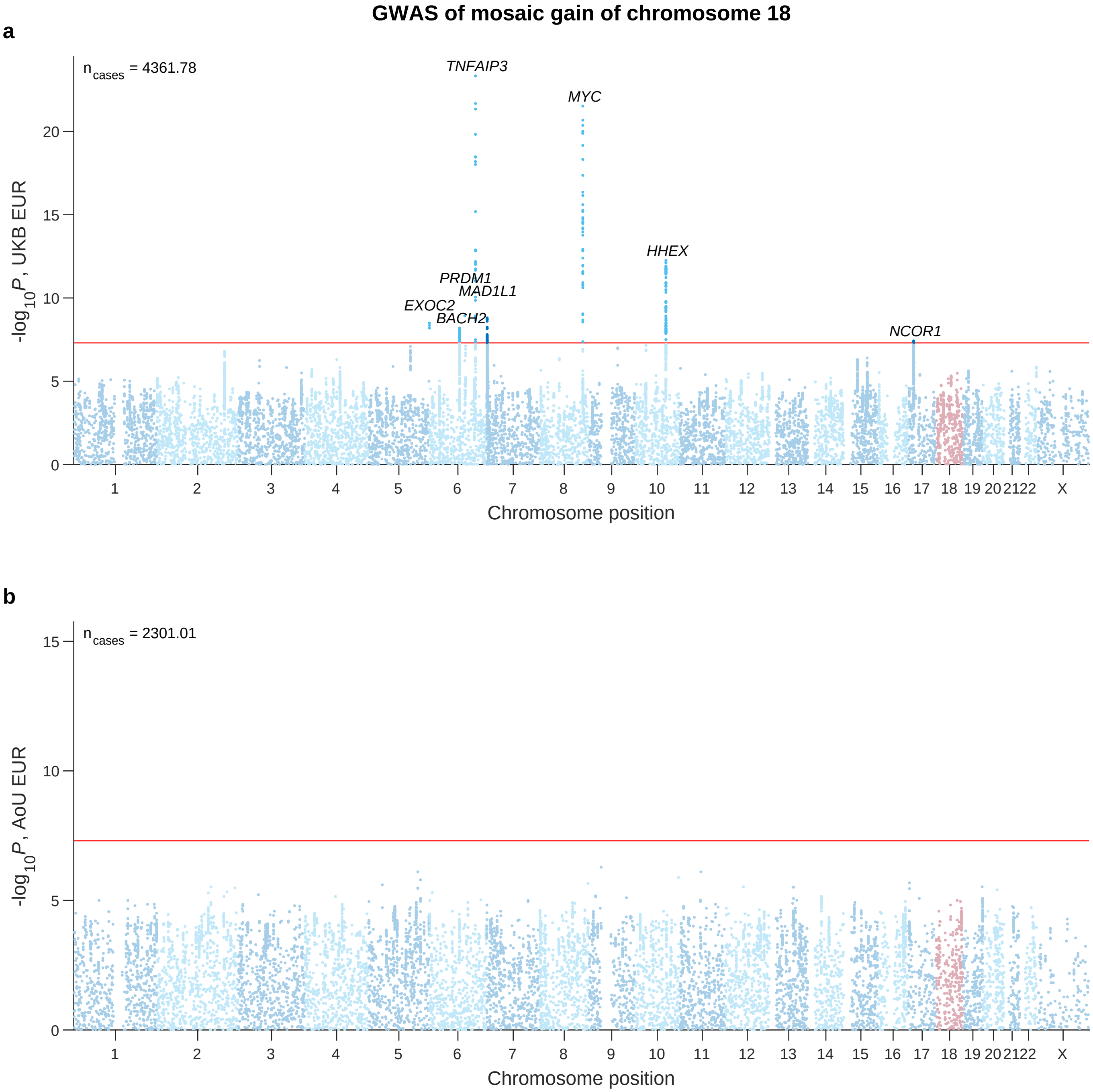

**Supplementary Figure 27. Associations of common inherited variants with mosaic gain of chromosome 18.** *P*-values are from linear regression on probabilistic calls of mosaic aneuploidies ($n_{cases}$ = cohort-wide sum) in unrelated individuals, with sex, age, age$^2$, smoking, and genetic principal components included as covariates (Methods). **a**, UKB EUR (n=404,400); **b**, AoU EUR (n=185,967). Lead variants ($P<5\times10^{-8}$) are labeled with the nearest gene.

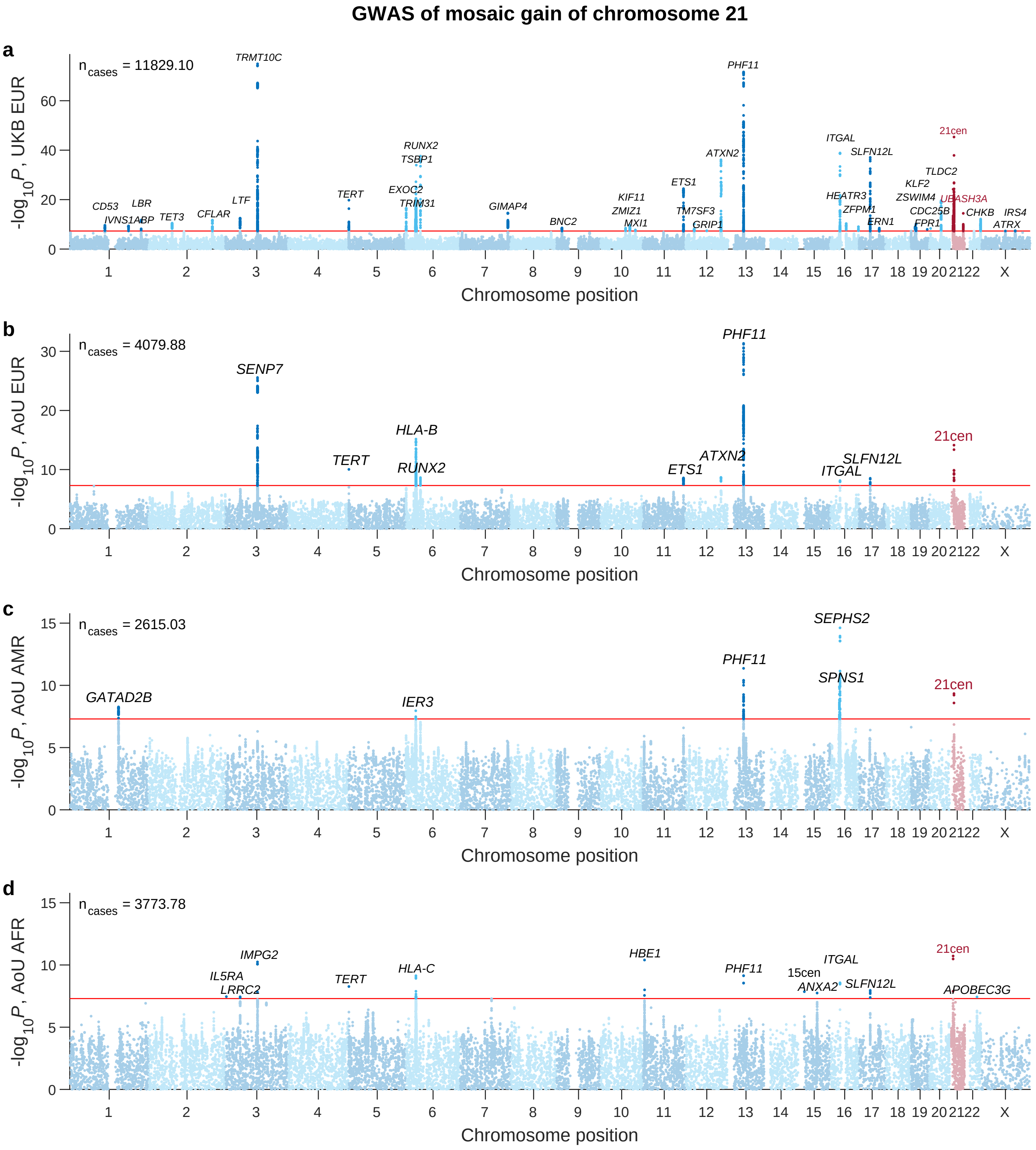

**Supplementary Figure 28. Associations of common inherited variants with mosaic gain of chromosome 21.** *P*-values are from linear regression on probabilistic calls of mosaic aneuploidies ($n_{cases}$ = cohort-wide sum) in unrelated individuals, with sex, age, age$^2$, smoking, and genetic principal components included as covariates (Methods). **a**, UKB EUR (n=404,136); **b**, AoU EUR (n=184,170); **c**, AoU AMR (n=61,896); **d**, AoU AFR (n=64,521). Lead variants ($P<5\times10^{-8}$) are labeled with the nearest gene.

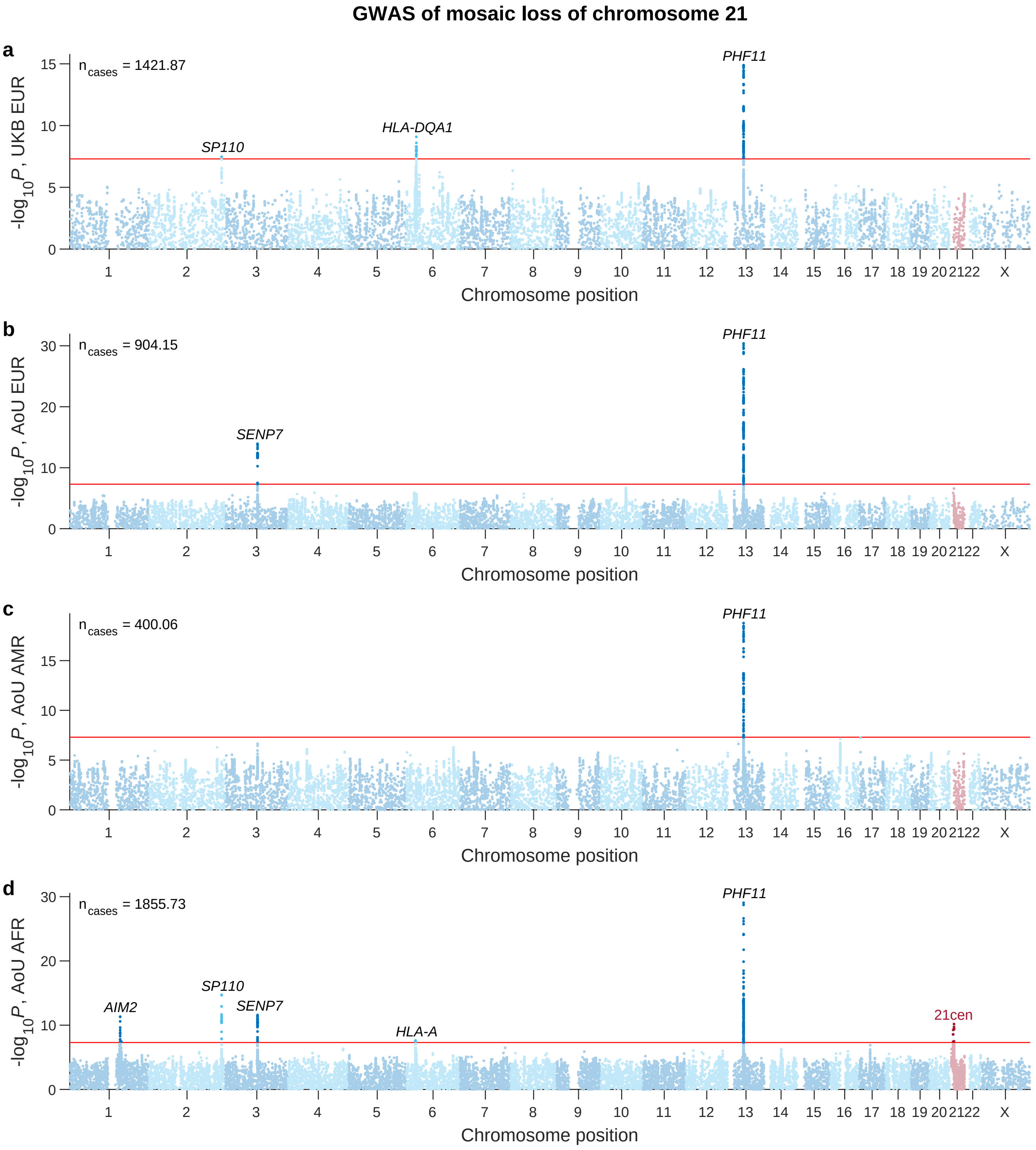

**Supplementary Figure 29. Associations of common inherited variants with mosaic loss of chromosome 21.** *P*-values are from linear regression on probabilistic calls of mosaic aneuploidies ($n_{cases}$ = cohort-wide sum) in unrelated individuals, with sex, age, age$^2$, smoking, and genetic principal components included as covariates (Methods). **a**, UKB EUR (n=396,630); **b**, AoU EUR (n=181,725); **c**, AoU AMR (n=59,897); **d**, AoU AFR (n=63,030). Lead variants ($P<5\times10^{-8}$) are labeled with the nearest gene.

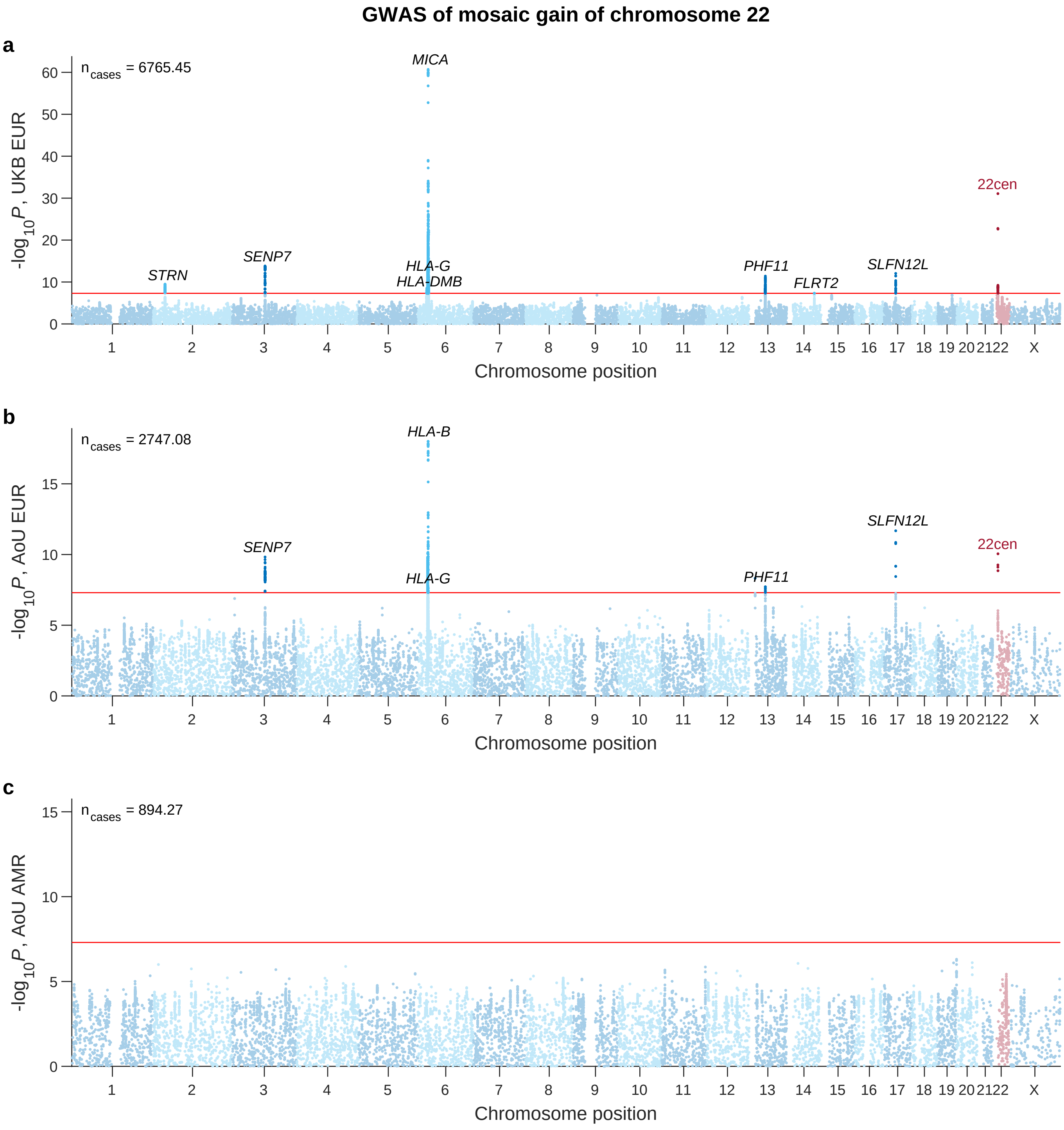

**Supplementary Figure 30. Associations of common inherited variants with mosaic gain of chromosome 22.** *P*-values are from linear regression on probabilistic calls of mosaic aneuploidies ($n_{cases}$ = cohort-wide sum) in unrelated individuals, with sex, age, age$^2$, smoking, and genetic principal components included as covariates (Methods). **a**, UKB EUR (n=407,008); **b**, AoU EUR (n=186,312); **c**, AoU AMR (n=62,857). Lead variants ($P<5\times10^{-8}$) are labeled with the nearest gene.

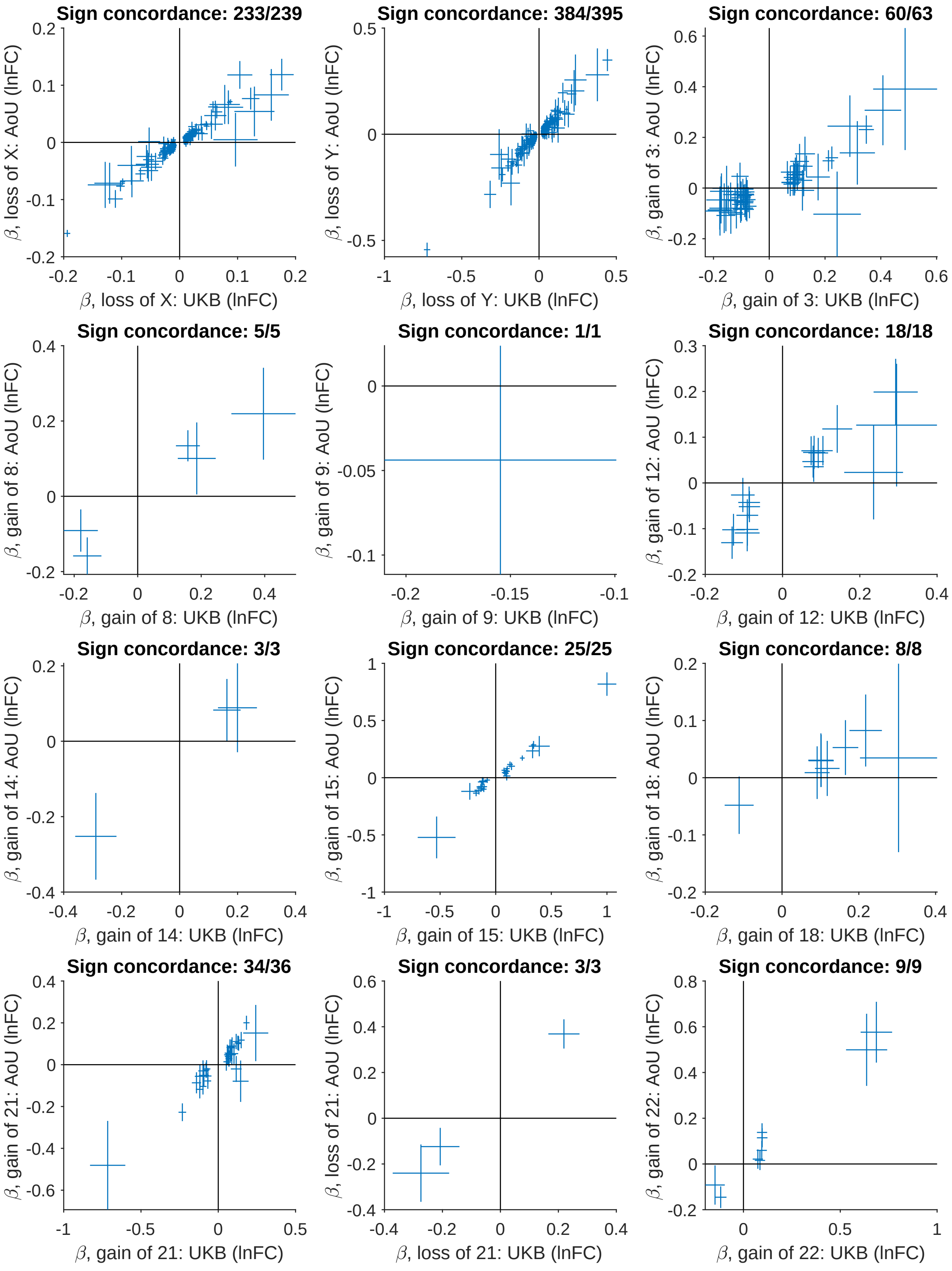

**Supplementary Figure 31. Replication in AoU of lead variant associations from UKB GWAS of mosaic aneuploidies.** (legend continued on next page)

**Supplementary Figure 31** ***(previous page)*****.** For lead variants identified from GWAS of 12 mosaic aneuploidy phenotypes in UKB, effect sizes in UKB (*x*-axis) are plotted against effect sizes in AoU (*y*-axis). For X and Y loss, the GWAS phenotypes were natural log-transformed, clipped X and Y loss cell fractions (Methods), such that effect sizes ($\beta$) are on this scale. For autosomal aneuploidies, the GWAS phenotype was a probabilistic call, so to facilitate interpretation, effect sizes ($\beta$) were transformed to the scale of natural log fold change in phenotype prevalence (Methods). A total of 828 lead variants (across the 12 aneuploidies) were initially considered for replication; of these, 14 mLOY-associated variants on chrX were excluded because of a known issue with the encoding of male chrX genotypes in the AoU v8 ACAF bgen file, and 9 additional variants were excluded because they were not genotyped in AoU, leaving 805 variants for which to attempt replication. For each aneuploidy, the number of variants for which the effect direction was consistent in UKB and AoU is indicated above the plot. Error bars, 95% CIs. GWAS sample sizes are provided in Supplementary Table 3.

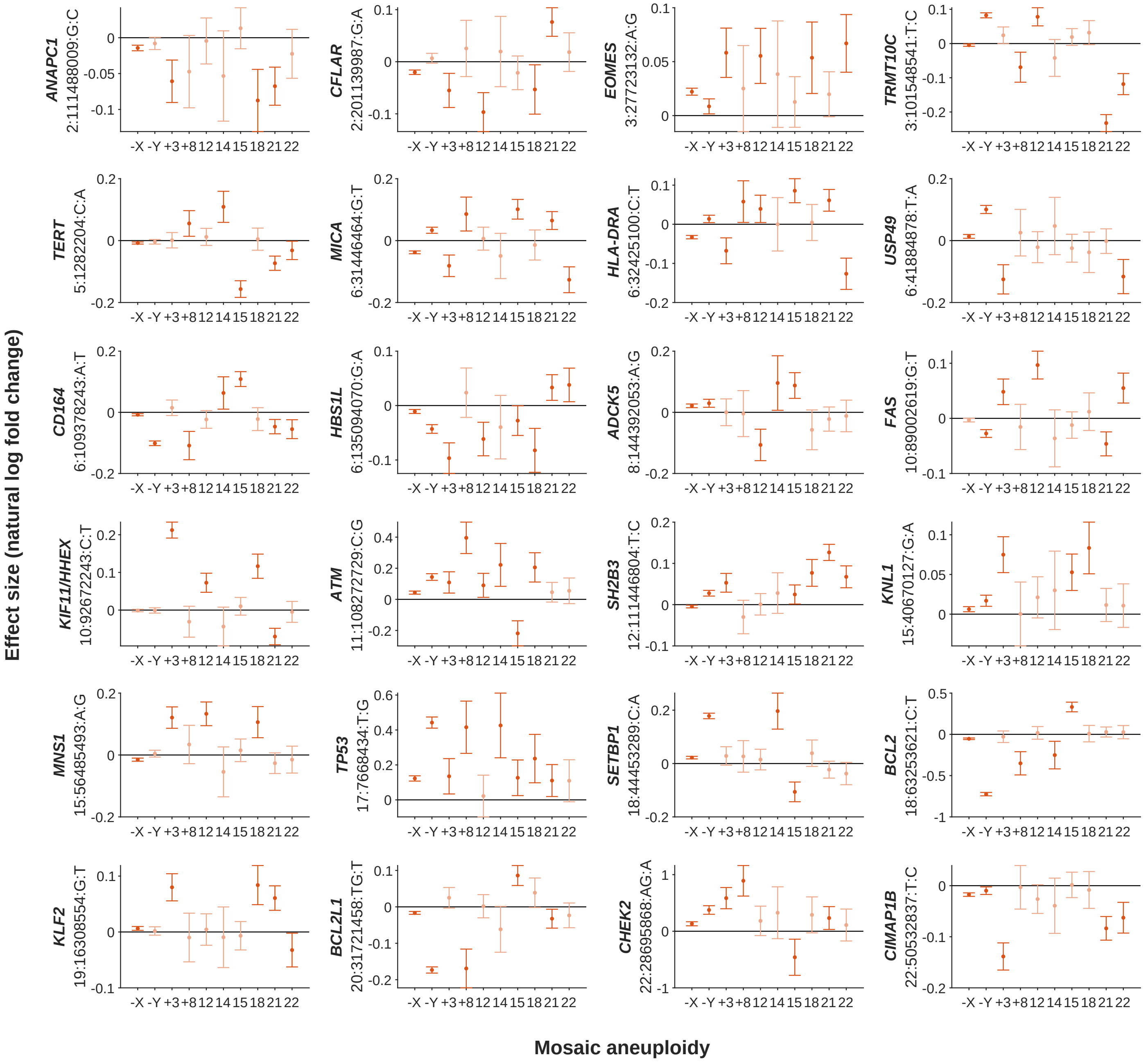

**Supplementary Figure 32. Effect sizes of inherited variants associated with several aneuploidy phenotypes.** Data are shown for variants associated with four or more aneuploidies, excluding variants already shown in Fig. 3 (which highlights genetic effects on chromosome segregation). Variant effect sizes (in the UKB EUR cohort) are in units of natural log fold change (of cell fraction for X and Y loss and of prevalence for autosomal aneuploidies; Methods). Darker colors indicate effects with 95% CIs (error bars) that do not overlap 0 ($P<0.05$). GWAS sample sizes are provided in Supplementary Table 3.

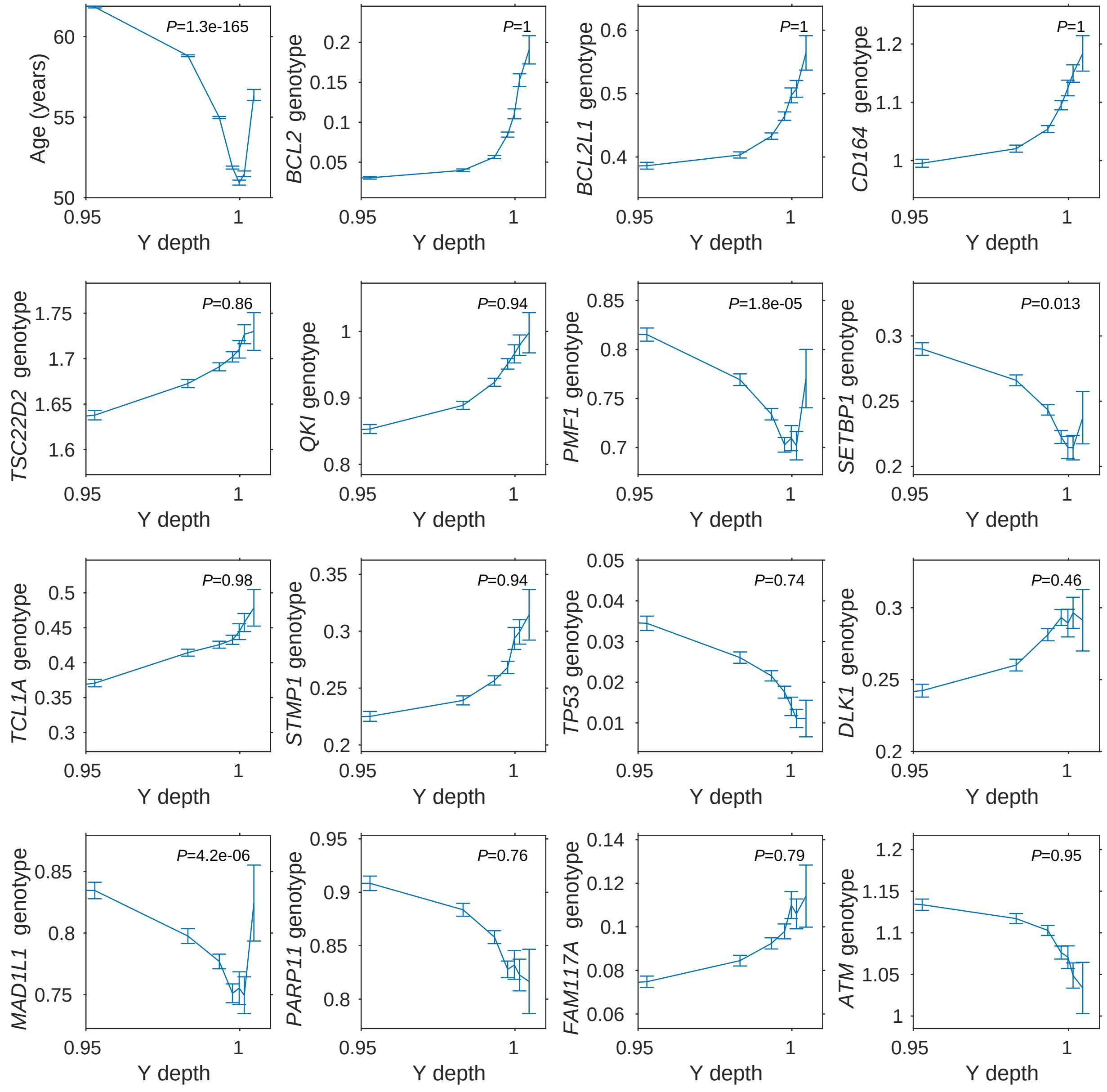

**Supplementary Figure 33. Association of *PMF1* and *MAD1L1* haplotypes with risk of both Y loss and mosaic gain of Y.** (legend continued on next page)

**Supplementary Figure 33** ***(previous page)*****.** The top-left plot shows mean age in different tranches of male EUR-ancestry UKB participants stratified by Y chromosome WGS read-depth (quantile boundaries of 0.01, 0.05, 0.25, 0.5, 0.75, 0.9, 0.95, 0.99). Individuals with decreased Y depth ($<1$) and those with increased Y depth ($>1$) both tend to be older, consistent with these deviations in Y depth being indicative of mosaic Y loss and gain of Y, respectively. The remaining fifteen plots show the mean genotype value (0/1/2) in each Y-depth tranche for each of the top fifteen lead variants from GWAS of Y loss in UKB (Supplementary Table 5). For most of the Y loss-associated variants, the mean genotype increases or decreases monotonically throughout the Y-depth range, consistent with the genetic effect being specific to Y loss (e.g., relating to clonal selection on cells with Y loss); in such a scenario, the monotonic trend is expected to extend through the range of Y-depth $>1$, as this indicates not only gain of Y but also absence of substantial Y loss. However, for the *PMF1* and *MAD1L1* lead variants, the trend inverts for Y-depth $>1$, consistent with these genetic effects influencing both Y loss and gain of Y (presumably by modulating the risk of Y-chromosome missegregation, which can result in either loss or gain of the Y chromosome). The *SETBP1* haplotype also exhibits some evidence of this behavior ($P$=0.013), but not to a level reaching Bonferroni significance. $P$-values are from a two-sample t-test comparing genotypes in the final tranche ($>99^{th}$ percentile) to genotypes in the preceding two tranches ($90^{th}$–$99^{th}$ percentiles), using a one-sided test for inversion of the trend observed in lower tranches. Error bars, 95% CIs.

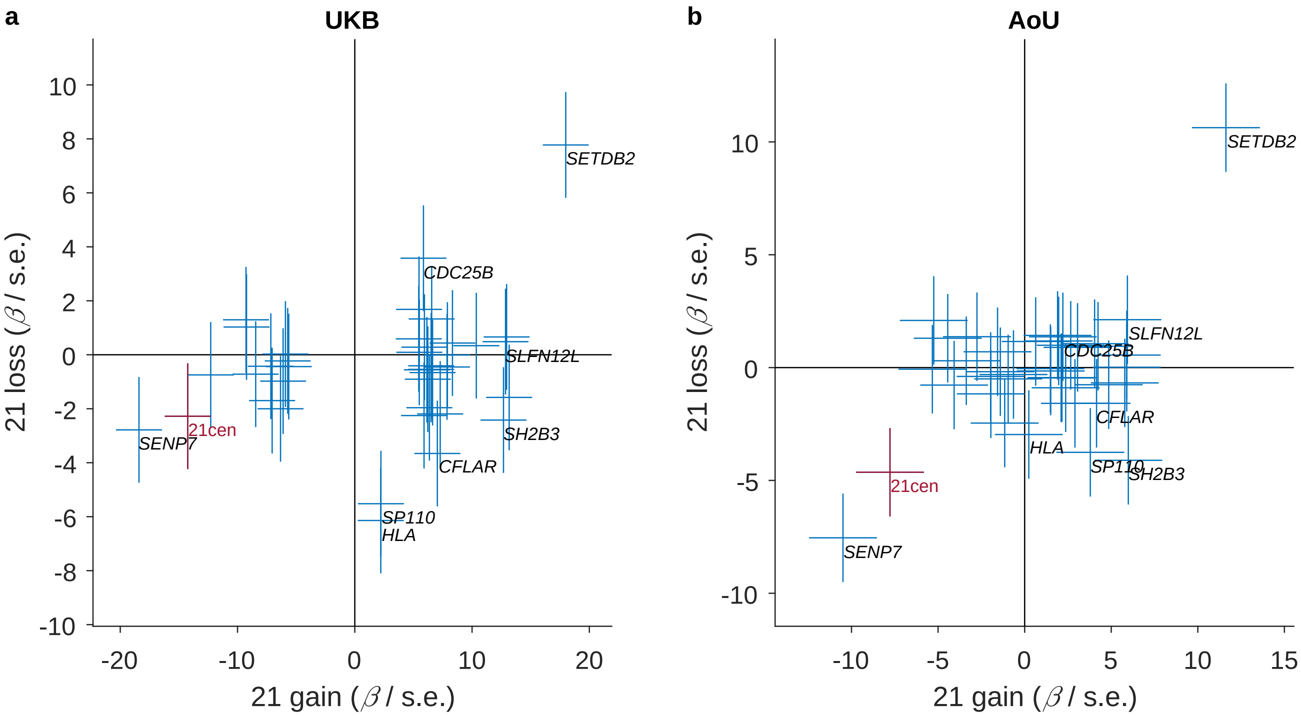

**Supplementary Figure 34. Comparison of associations of inherited variants with mosaic loss versus gain of chromosome 21.** Association $z$-statistics ($\beta$/s.e.) for loss of 21 versus gain of 21 in EUR-ancestry participants from (**a**) UKB and (**b**) AoU are plotted against each other. Error bars, 95% CIs. For both plots, data are presented for a set of variants obtained by merging lead variants from GWAS of loss of 21 and GWAS of gain of 21 in UKB. Lead variants were deduped across these two GWAS by identifying pairs of nearby variants with similar associations to both traits ($z$ values within a factor of 1.25 for both traits). The lead variant in UKB at the chromosome 21 centromere (21:13216980:A:G) was not genotyped in AoU, so in panel **b**, the corresponding lead variant in AoU (21:13133256:C:A) was substituted. Three of these genetic effects (at *SETDB2*, the chromosome 21 centromere, and *SENP7*) show strong evidence of acting consistently on both loss and gain of chromosome 21. This suggests that these genetic effects influence risk of missegregation, which can result in either loss or gain of chromosome 21. The *SLFN12L* locus also exhibits some evidence of this behavior. GWAS sample sizes are provided in Supplementary Table 3.

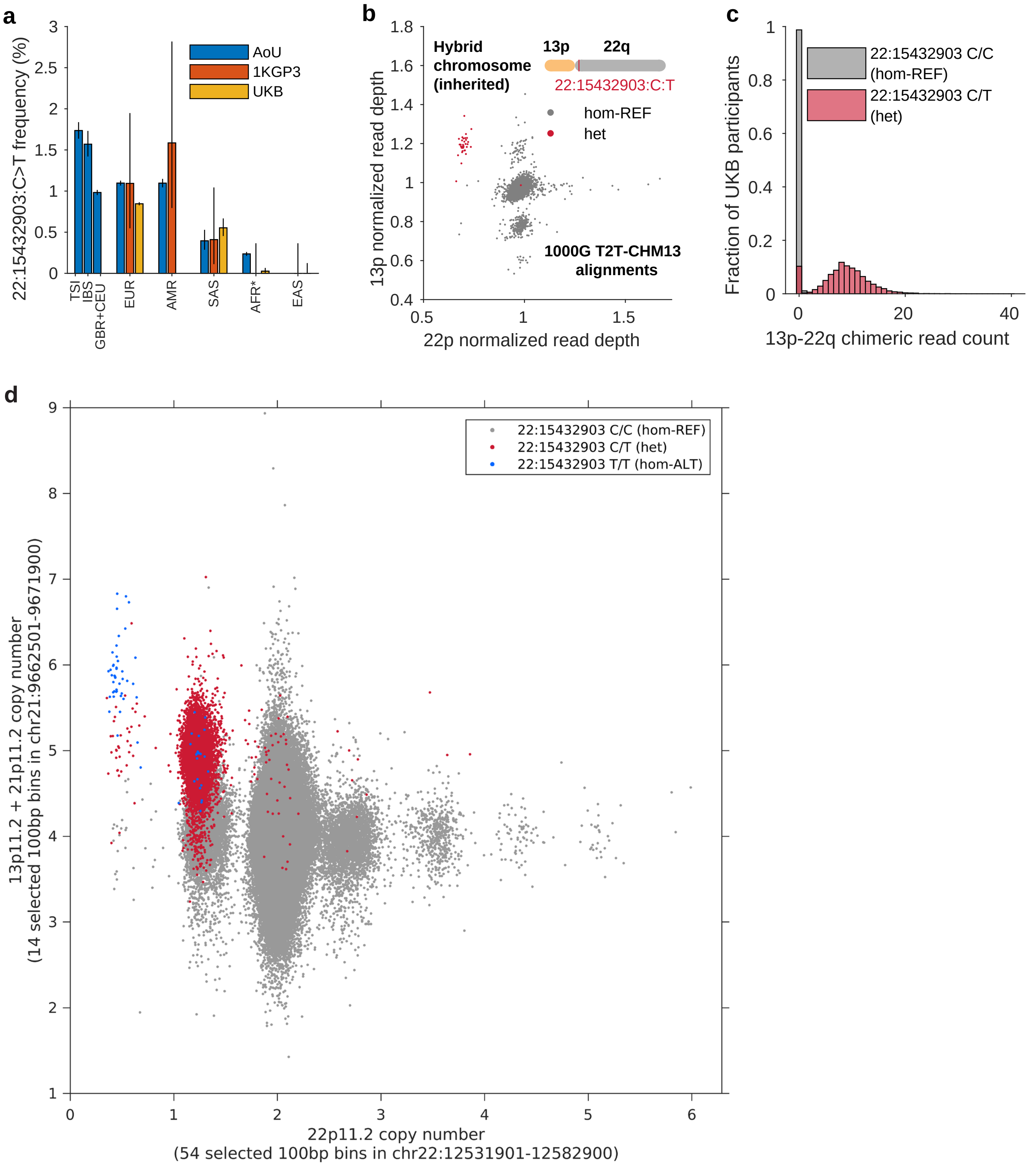

**Supplementary Figure 35. Population frequencies and tagging of the 13p–22q ectopic recombination by 22:15432903:C>T.** (legend continued on next page)

**Supplementary Figure 35** ***(previous page)*****.** **a**, Frequencies of the tag SNP 22:15432903:C>T across genetic ancestries (as previously defined for AoU[28] and UKB[81]). Within-EUR ancestry assignments were defined for AoU as described in Supplementary Note 5. *AFR ancestry for 1KGP3 excluded the admixed ACB and ASW populations (but AoU AFR included admixed individuals). Error bars, 95% CIs. **b**, Scatter plot of normalized read depth within selected genomic bins on 13p versus 22p for 1000 Genomes WGS alignments to T2T-CHM13 (n=3,202; Supplementary Note 5). Among 35 carriers of the tag SNP (red), 34 have aberrant read-depth supporting the 13p-22q recombination, and only one individual does not (in the center of the large cluster; HG01088 from the PUR population). **c**, Distributions of chimeric read counts supporting the 13p-22q recombination (Supplementary Note 5) among UKB participants heterozygous for the tag SNP (red) and among noncarriers (gray). **d**, Scatter plot of normalized read depth within genomic bins in 21p11.2 (also representing 13p11.2 in GRCh38) versus 22p11.2 for UKB WGS alignments to GRCh38 (Supplementary Note 5). Individuals are colored by genotype of the tag SNP, which segregates nearly perfectly with the cluster of points with increased 13p11.2 + 21p11.2 read-depth and reduced 22p11.2 read-depth. The multiple gray clusters with anomalous 22p11.2 read-depth but normal 13p11.2 + 21p11.2 read-depth may reflect copy-number variation at 22p11.2.

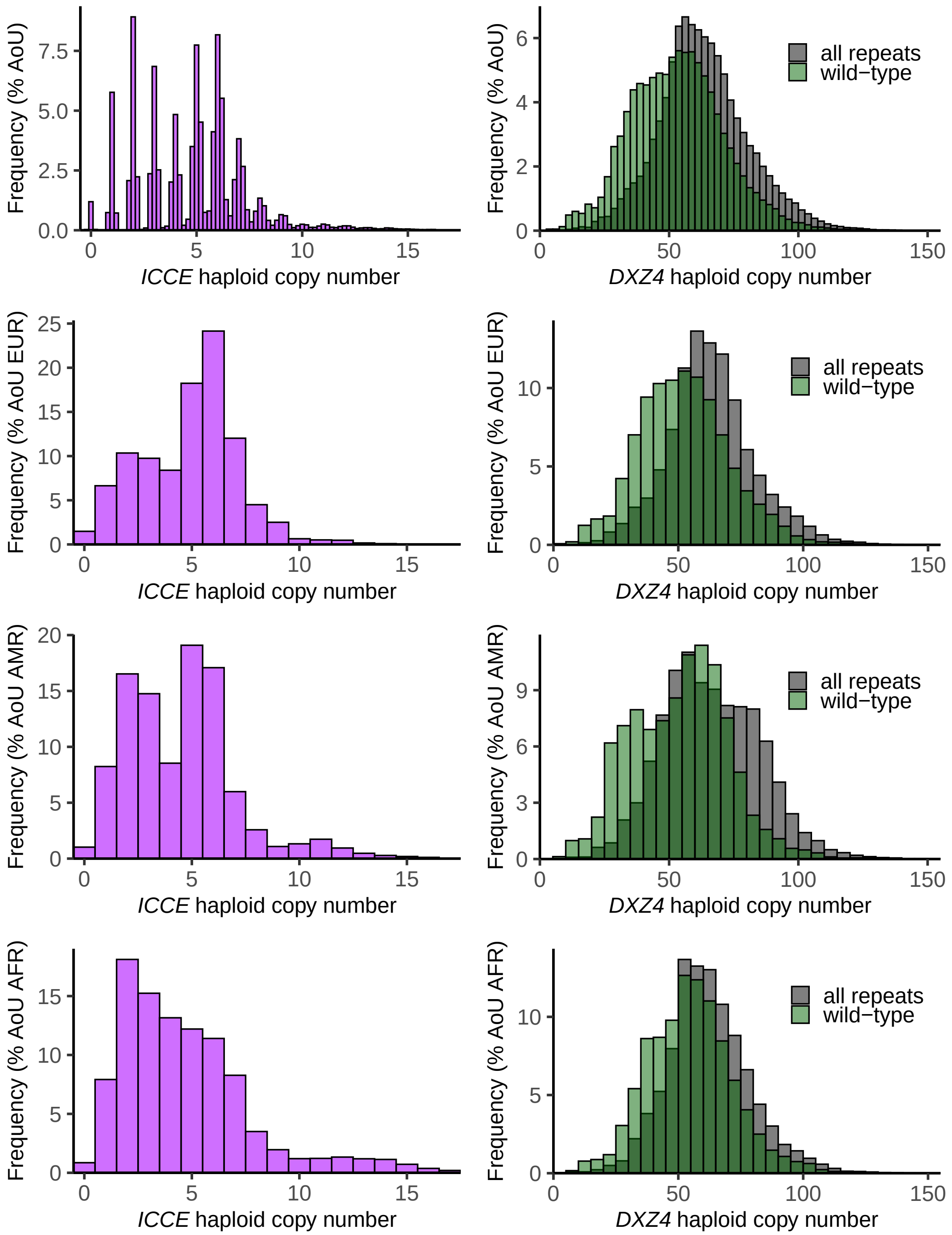

**Supplementary Figure 36. Copy-number distributions of the *ICCE* and *DXZ4* repeat arrays on chromosome X.** The top plots use finer-resolution binning (0.2 repeat units for *ICCE*, 2.5 repeats for *DXZ4*) to provide an indication of the amount of noise in the copy-number estimates. For *DXZ4*, total copy number and wild-type copy number (subtracting the numbers of copies of the PSVs impacting the CTCF and YY1 binding sites) are shown. Top, n=137,798 male AoU participants (merged across three ancestries); subsequent rows, EUR (n=80,730), AMR (n=24,507), AFR (n=32,561). Allele-specific (haploid) copy numbers estimated for females using statistical phasing exhibited consistent distributions.

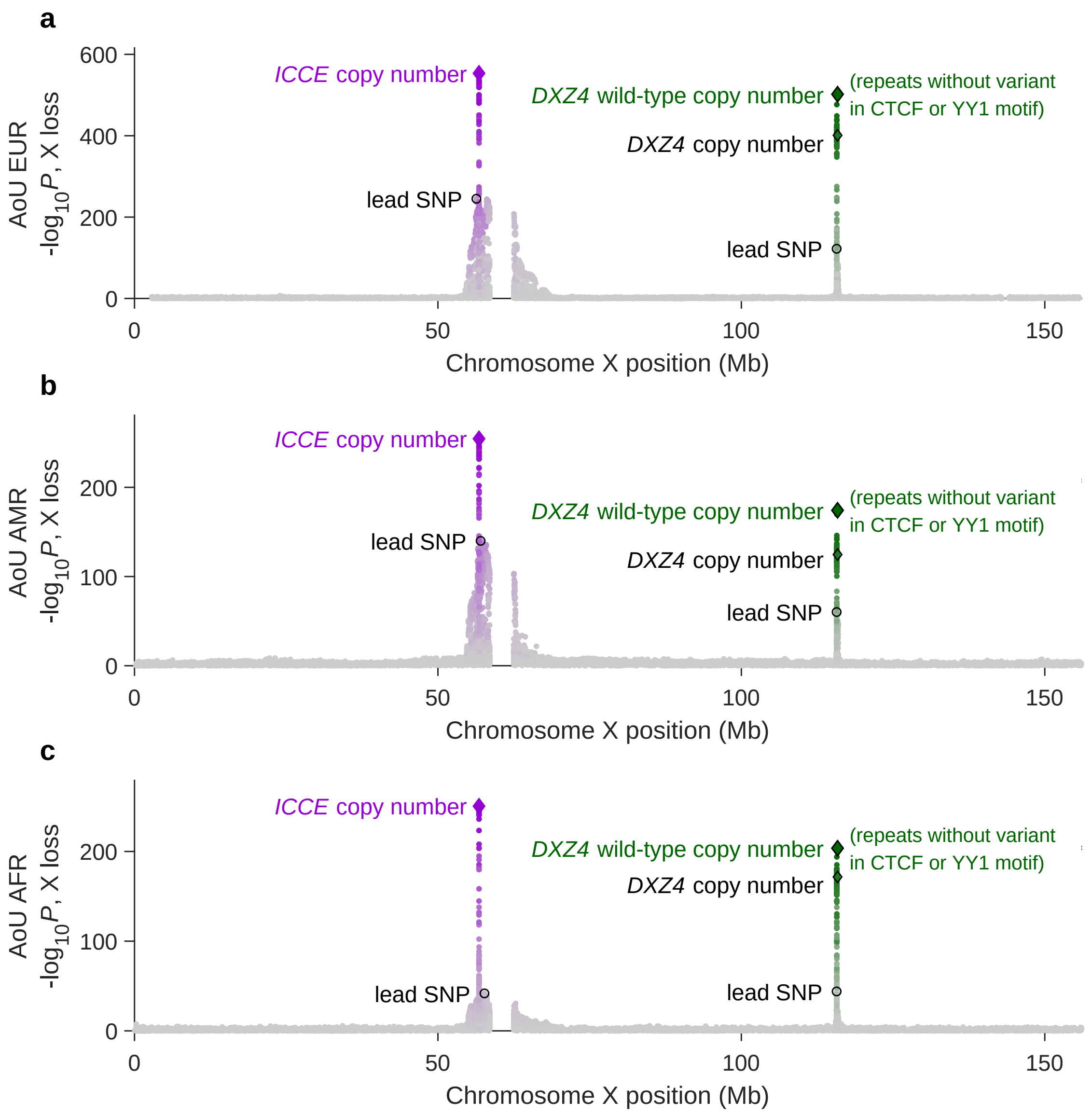

**Supplementary Figure 37. Associations between inherited X chromosome variants and X loss levels in female AoU participants.** *P*-values are from linear regression on log-transformed, clipped cell fractions (Methods) using age, age$^2$, smoking, and 16 genetic principal components as covariates. Association analyses were performed on unrelated female EUR-ancestry (n=112,092), AMR-ancestry (n=42,005), and AFR-ancestry (n=39,629) AoU participants. Variants near the *ICCE* and *DXZ4* loci are colored according to linkage disequilibrium ($R^2$) with copy number of the *ICCE* repeat (purple) and wild-type copy number of the *DXZ4* repeat (green; i.e., the number of repeat units not containing the PSVs impacting the CTCF and YY1 binding sites).

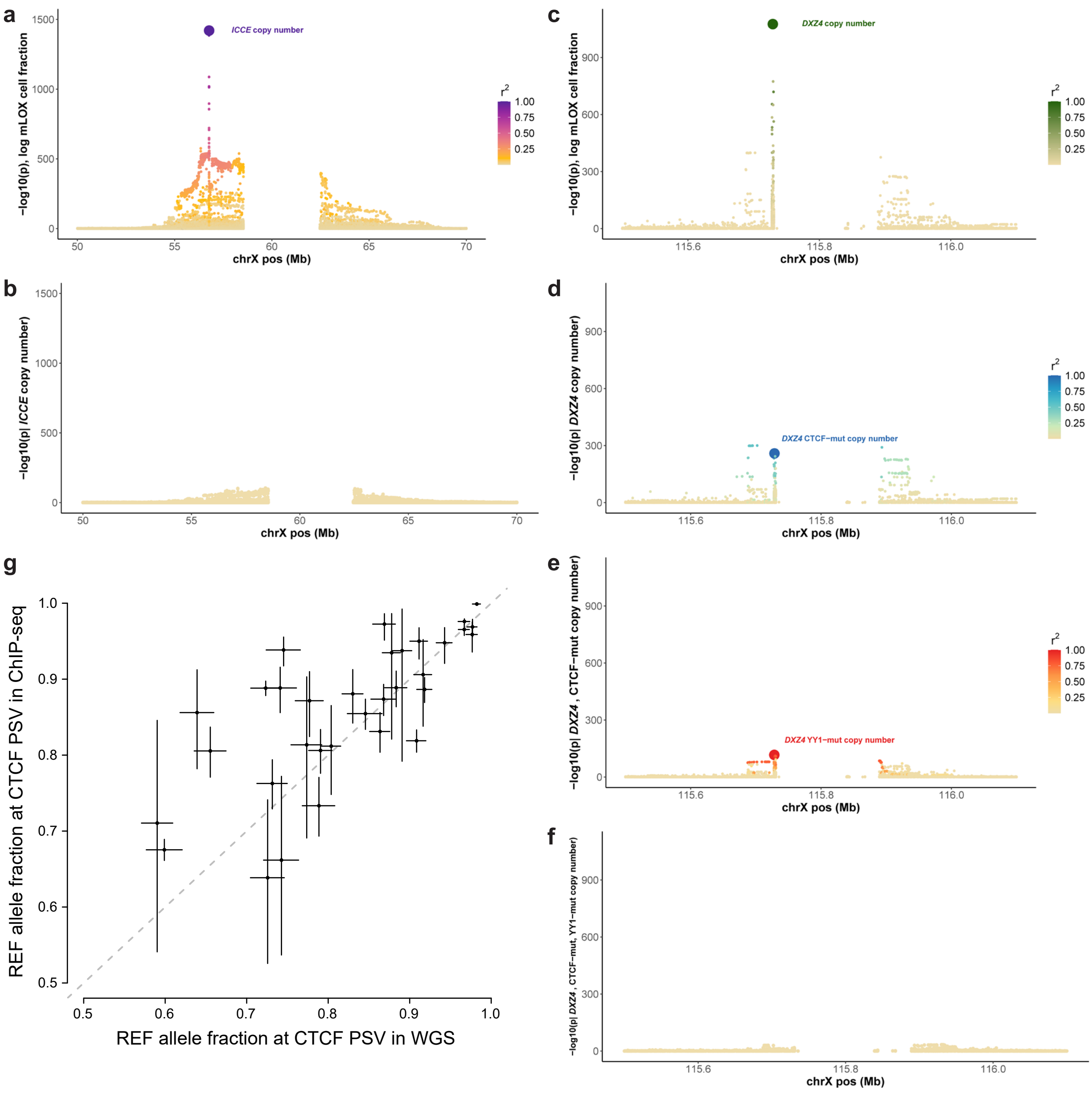

**Supplementary Figure 38. X loss stepwise conditional association analyses at *ICCE* and *DXZ4* and reduced CTCF binding to *DXZ4* CTCF motif variant. a–f**, *P*-values from stepwise conditional association analyses of X loss (linear regression with age, age$^2$, smoking, and 20 PCs as covariates in the UKB female EUR cohort; n=241,912 with available *ICCE* and *DXZ4* genotypes). *P*-values are plotted for SNPs, indels, *ICCE* and *DXZ4* repeat copy numbers, and copy numbers of minor alleles of intra-repeat SNP and indel variants. To facilitate interpretation of these stepwise conditional analyses, copy numbers of major alleles of *ICCE* and *DXZ4* PSVs are not plotted here (unlike in Fig. 4 and Supplementary Fig. 37). Markers are colored according to LD with the indicated copy-number polymorphism ($r^2$ in a random downsample; n=10,000). **g**, Allelic skew at the common *DXZ4* CTCF motif variant (represented by chrX:115728581:T:C) in CTCF ChIP-seq data from 1000 Genomes LCLs (Supplementary Note 6). The REF allele is overrepresented in ChIP-seq reads relative to WGS reads, indicating greater CTCF binding relative to the variant motif.

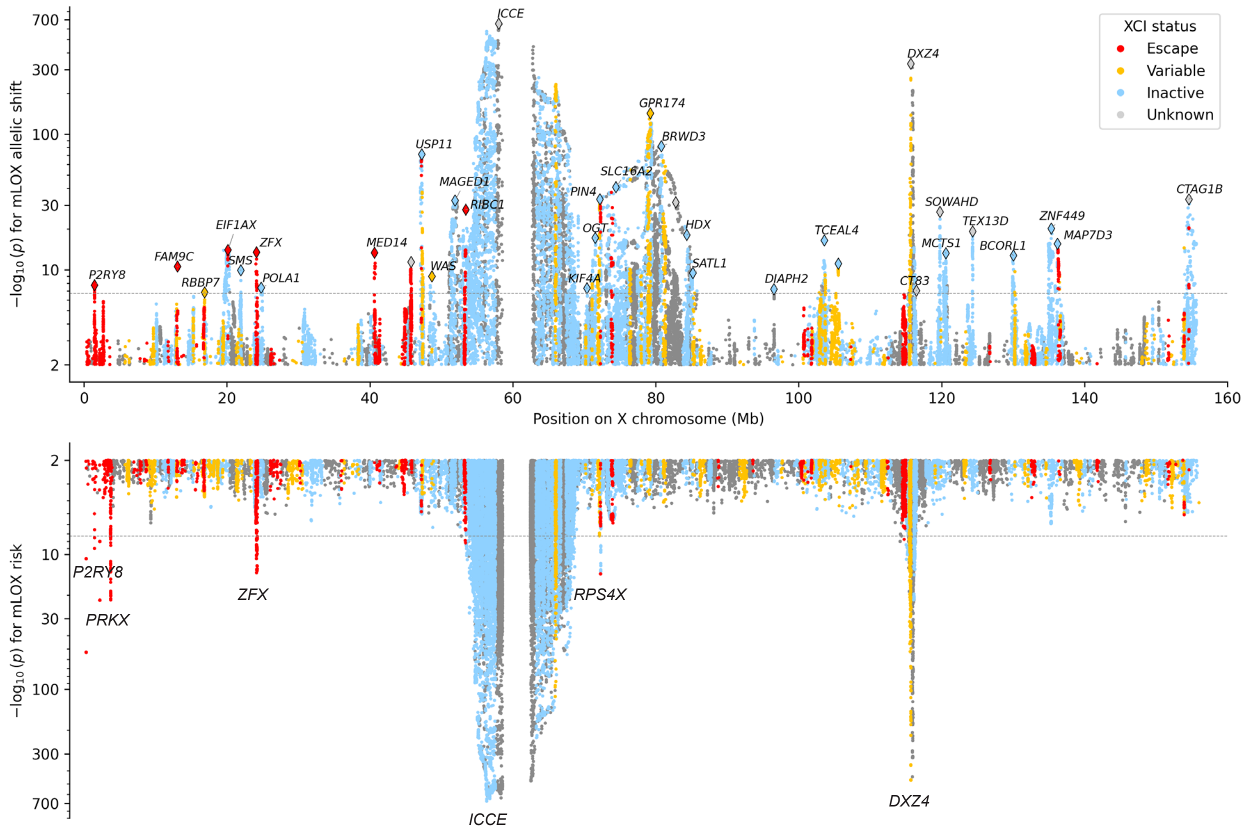

**Supplementary Figure 39. Associations between inherited X chromosome variants and X loss phenotypes (allelic bias and cell fraction) in UKB.** Top: $-\log_{10}$ $P$-values (plotted on the log scale) from tests for association between inherited SNP/indel variants on the X chromosome and allelic bias in X loss. For each variant, an allelic imbalance phenotype was generated for individuals heterozygous for the variant (by applying a sigmoid transformation to the difference between the fractions of cells having lost the homolog carrying the REF allele versus the homolog carrying the ALT allele), and this phenotype was tested for a shift toward either the REF or the ALT allele. Bottom: Association strengths from GWAS of log-transformed, clipped X loss cell fraction from BOLT-LMM on UKB EUR-ancestry female participants (n=242,531). Variants are colored according to the escape status of the nearest gene within 100 kb. The stray dots in the PAR1 region (bottom left) are from variants that failed subsequent QC for technical artifacts; this plot will be regenerated when the UKB-RAP platform comes back online.

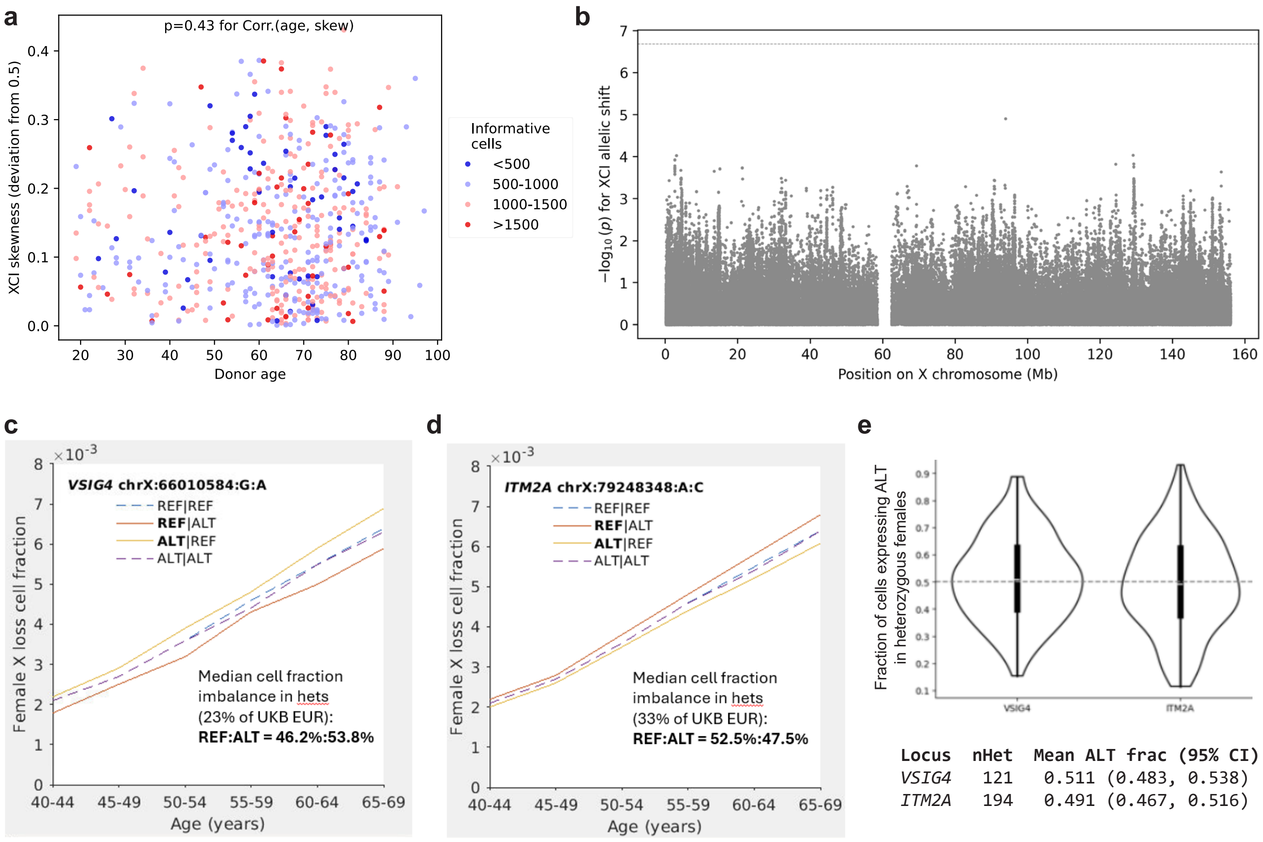

**Supplementary Figure 40. No detectable allelic biases in X inactivation skewing. a**, Estimates of X chromosome inactivation (XCI) skewing in peripheral blood mononuclear cells from 565 female donors in the OneK1K cohort. Each data point corresponds to an individual; for each individual, the estimated amount of skewing is plotted against the individual's age. Points are colored by the number of cells for which the inactivated X chromosome homolog was determined. XCI skewing was common and not age-associated ($P$=0.43). **b**, $-\log_{10}$ $P$-values from tests for association between phase orientation of common X chromosome variants and XCI skew direction in heterozygous carriers (Supplementary Note 7). The dashed horizontal line indicates the Bonferroni significance threshold; no statistically significant allelic biases in XCI skew were observed. **c**, **d**, Median haplotype-resolved X loss cell fractions in female UKB participants stratified by age and by phased genotypes of the lead variants at *MIR223HG–VSIG4* and *ITM2A* (on chromosome X) associated with X loss allelic shift. In the legend, the first allele (REF or ALT) is the allele in *cis* and the second is the allele in *trans*. Heterozygous carriers exhibit higher fractions of cells with loss of the ALT allele at *VSIG4* and the REF allele at *ITM2A*. **e**, Distribution of the fraction of cells expressing the ALT allele across OneK1K donors heterozygous for the *VSIG4* and *ITM2A* variants associated with X loss allelic shift. Although the error bars are wide, the data are more consistent with equal inactivation of the REF and ALT alleles than with the imbalance observed in X loss, suggesting that the allelic shift in X loss at these loci does not derive from biased XCI.

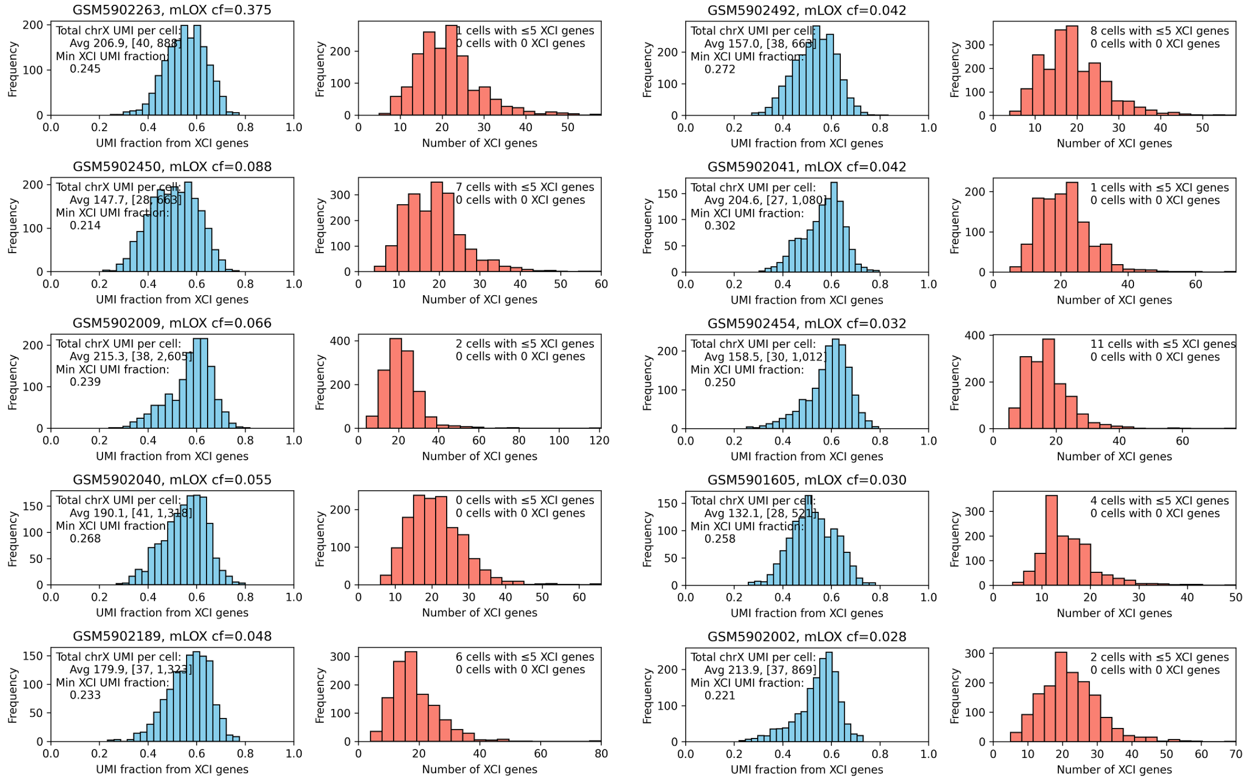

**Supplementary Figure 41. Evidence from single-cell RNA sequencing that X loss always involves loss of the inactive X chromosome.** Each pair of plots corresponds to a female OneK1K participant with substantial X loss (cell fraction 2.8%–37.5% estimated by MoChA[14,15]). The histograms provide distributions (across each donor's RNA-sequenced single cells) of the following quantities. Blue histograms: Combined expression level of 386 genes that do not escape X inactivation ("XCI genes") as a fraction of the combined expression of all genes on the X chromosome. These distributions do not show substantial skewing; no donor has any cells exhibiting unusually low expression of non-escape genes. Red histograms: numbers of non-escape genes with nonzero expression. Again, every cell in every donor exhibits expression of at least one gene that does not escape XCI. These data indicate that even in individuals who had lost an X chromosome in a substantial fraction of leukocytes, every sampled leukocyte had an active X chromosome, suggesting that the inactive X chromosome is always the one that is lost.

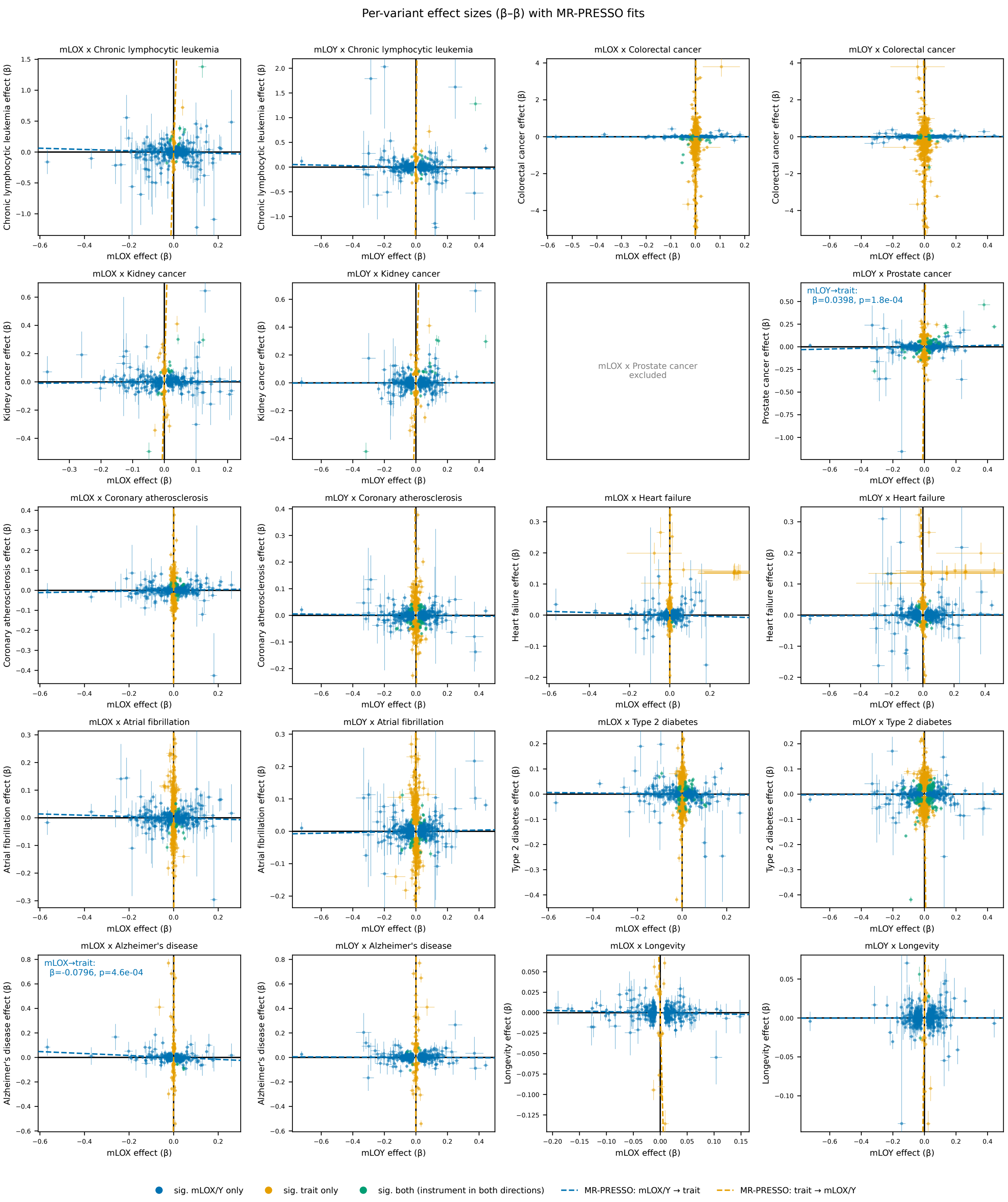

**Supplementary Figure 42. Beta-beta plots (i.e., effect-size scatter plots) for Mendelian randomization analyses of LOX and LOY with diseases and longevity.**

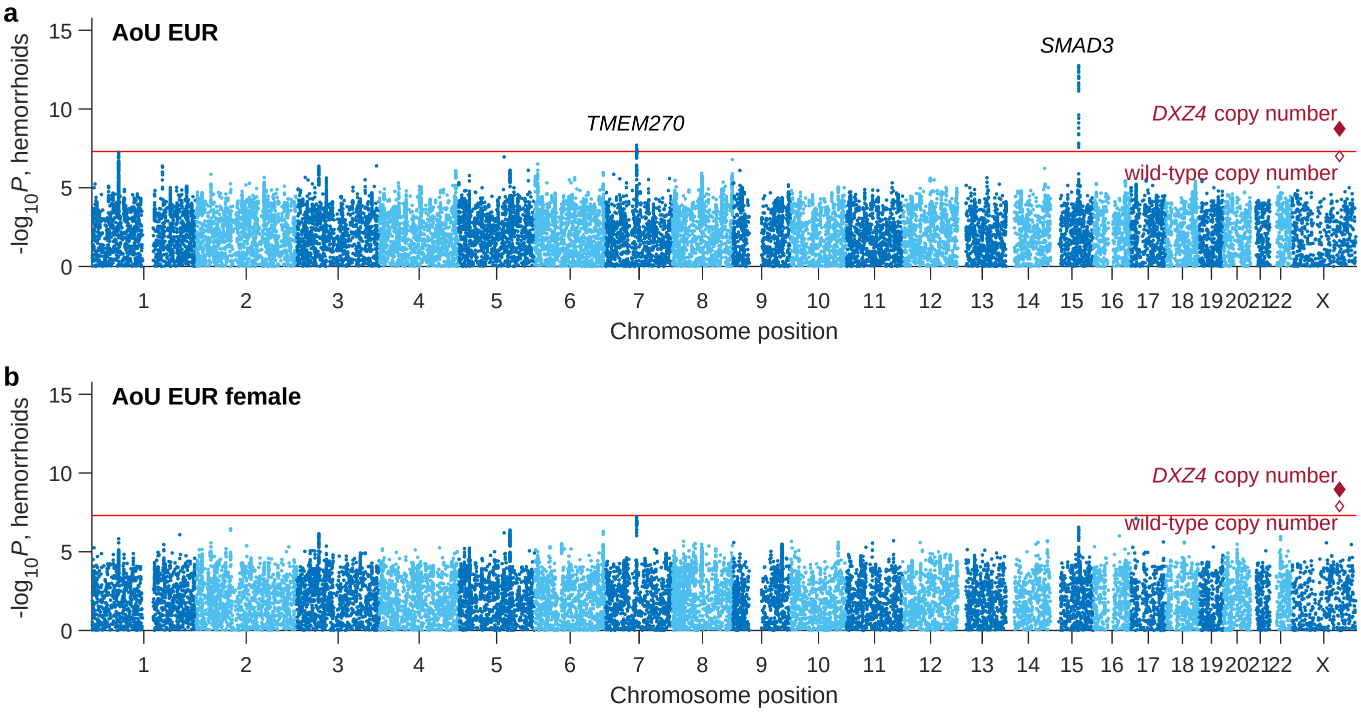

**Supplementary Figure 43. Associations of common inherited variants with self-reported hemorrhoids in AoU.** *P*-values are from linear regression on case/control status for hemorrhoids using age, age$^2$, smoking, and 16 genetic principal components as covariates. Association analyses were performed on unrelated EUR-ancestry AoU participants of (**a**) both sexes (n=125,764; 31,832 cases and 93,932 controls) or (**b**) female sex only (n=79,220; 20,684 cases and 58,536 controls). Diamonds indicate associations with copy number of the *DXZ4* repeat; the unfilled diamond indicates wild-type copy number (i.e., the number of repeat units not containing the PSVs impacting the CTCF and YY1 binding sites).

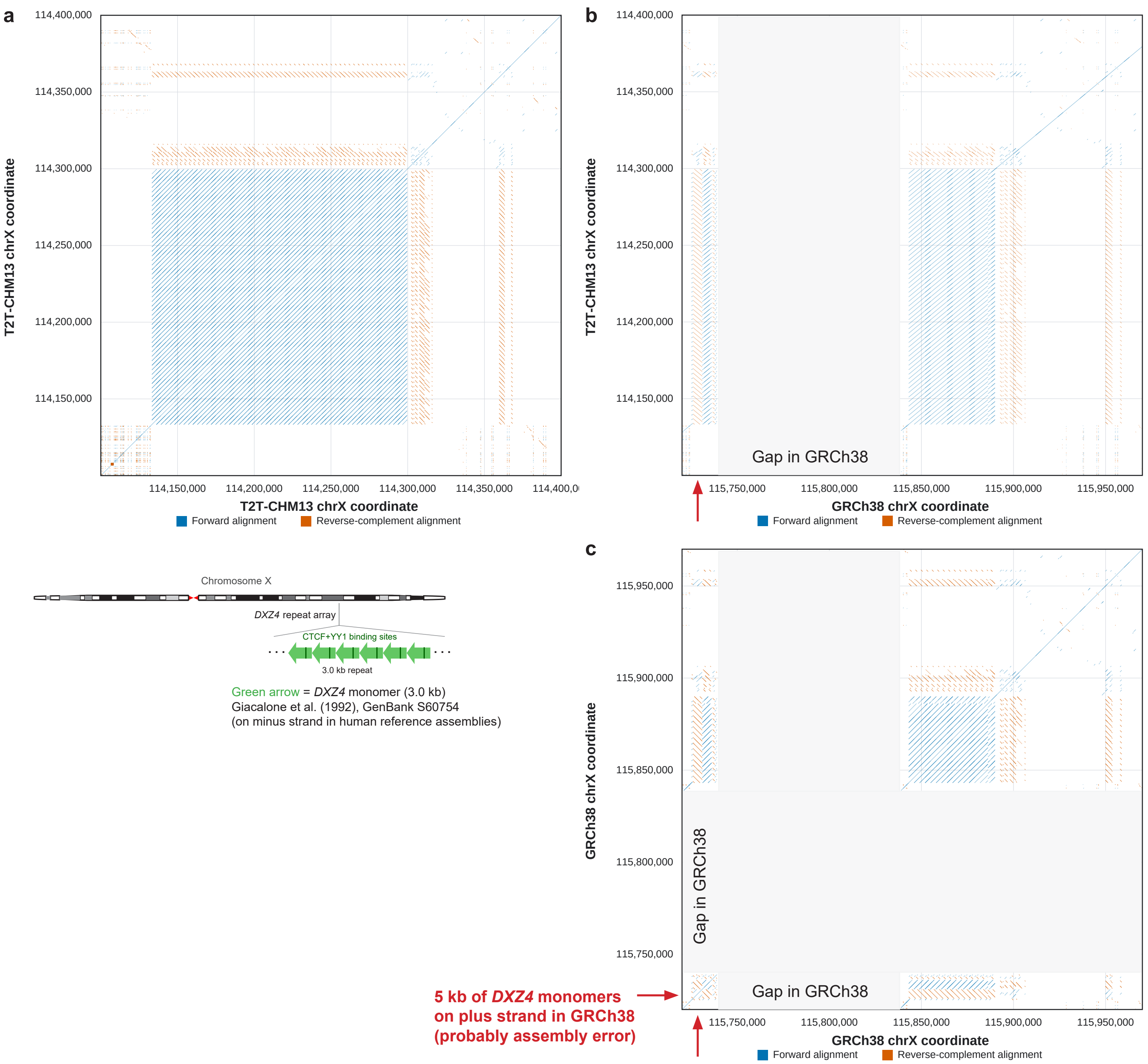

**Supplementary Figure 44. *DXZ4* repeat arrays in T2T-CHM13 and GRCh38.** Dot plots show regions of local sequence homology within and between the *DXZ4* loci in T2T-CHM13 (chrX:114.1–114.4 Mb) and GRCh38 (chrX:115.72–115.97 Mb). Blue/red dots indicate alignments on the same/opposite strand, computed using NCBI BLASTN[142] v2.17.0. The GRCh38 assembly contains a large gap within *DXZ4*, with most of the *DXZ4* repeat array located on the right side of the gap. A few repeat units are also situated immediately to the left of the gap, including a 5-kb sequence (farthest left) inverted relative to the rest of the array. This inversion probably reflects an assembly error, but because it has the same orientation as the original *DXZ4* monomer (GenBank S60754)[62], we used a repeat unit within it as a reference (GRCh38 chrX:115727764–115730717).

**Supplementary Table 1.** Standard errors of LOX and LOY cell fractions estimated using different approaches.

**Supplementary Table 2.** Accuracy of denoised WGS read depth and rates of binary aneuploidy calls in AoU using fixed or chromosome-specific cell fraction thresholds.

**Supplementary Table 3.** Cohort statistics for GWAS of aneuploidy phenotypes in UKB and AoU.

**Supplementary Table 4.** Lead associations from GWAS of X loss in UKB EUR and replication in AoU EUR.

**Supplementary Table 5.** Lead associations from GWAS of Y loss in UKB EUR and replication in AoU EUR.

**Supplementary Table 6.** Lead associations from GWAS of autosomal aneuploidies in UKB EUR and replication in AoU EUR.

**Supplementary Table 7.** Lead associations from GWAS of X loss in the AoU EUR, AMR, and AFR cohorts.

**Supplementary Table 8.** Lead associations from GWAS of Y loss in the AoU EUR, AMR, and AFR cohorts.

**Supplementary Table 9.** Lead associations from GWAS of autosomal aneuploidies in the AoU EUR, AMR, and AFR cohorts.

**Supplementary Table 10.** Association tests for allelic imbalance (shift) of lead variants associated with mosaic gain of the same autosome.

**Supplementary Table 11.** Associations from gene-based burden tests for X loss in UKB EUR.

**Supplementary Table 12.** Associations from gene-based burden tests for Y loss in UKB EUR.

**Supplementary Table 13.** Associations from gene-based burden tests for autosomal aneuploidies in UKB EUR.

**Supplementary Table 14.** Centromere haplotypes independently associated with mosaic gain of the same chromosome.

**Supplementary Table 15.** Normalized WGS read-depth on acrocentric chromosomes for 1000 Genomes participants.

**Supplementary Table 16.** Lead variants on the X chromosome associated with loss of X in cis or trans in the UKB EUR cohort.

**Supplementary Table 17.** GWAS summary statistics used for Mendelian randomization analyses.

**Supplementary Table 18.** Mendelian randomization analyses of disease phenotypes and longevity using X and Y loss as either the exposure or the outcome.
